# Population dynamics of HIV drug resistance during treatment scale-up in Uganda: a population-based longitudinal study

**DOI:** 10.1101/2023.10.14.23297021

**Authors:** Michael A. Martin, Steven James Reynolds, Brian T. Foley, Fred Nalugoda, Thomas C. Quinn, Steven A. Kemp, Margaret Nakalanzi, Edward Nelson Kankaka, Godfrey Kigozi, Robert Ssekubugu, Ravindra K. Gupta, Lucie Abeler-Dörner, Joseph Kagaayi, Oliver Ratmann, Christophe Fraser, Ronald Moses Galiwango, David Bonsall, M. Kate Grabowski, the PANGEA-HIV Consortium, the Rakai Health Sciences Program

## Abstract

**Background:** Clinical studies have reported rising pre-treatment HIV drug resistance during antiretroviral treatment (ART) scale-up in Africa, but representative data are limited. We estimated population-level drug resistance trends during ART expansion in Uganda

**Methods:** We analyzed data from the population-based open Rakai Community Cohort Study conducted at agrarian, trading, and fishing communities in southern Uganda between 2012 and 2019. Consenting participants aged 15-49 were HIV tested and completed questionnaires. Persons living with HIV (PLHIV) provided samples for viral load quantification and virus deep-sequencing. Sequence data were used to predict resistance. Population prevalence of class-specific resistance and resistance-conferring substitutions were estimated using robust log-Poisson regression.

**Findings:** Data from 93,622 participant-visits, including 4,702 deep-sequencing measurements, showed that the prevalence of NNRTI resistance among pre-treatment viremic PLHIV doubled between 2012 and 2017 (PR:1.98, 95%CI:1.34–2.91), rising to 9.61% (7.27-12.7%). The overall population prevalence of pre-treatment viremic NNRTI and NRTI resistance among all participants decreased during the same period, reaching 0.25% (0.18% - 0.33%) and 0.05% (0.02% - 0.10%), respectively (*p-*values for trend = 0.00015, 0.002), coincident with increasing treatment coverage and viral suppression. By the final survey, population prevalence of resistance contributed by treatment-experienced PLHIV exceeded that from pre-treatment PLHIV, with NNRTI resistance at 0.54% (0.44%-0.66%) and NRTI resistance at 0.42% (0.33% - 0.53%). Overall, NNRTI and NRTI resistance was predominantly attributable to rtK103N and rtM184V. While 10.52% (7.97%-13.87%) and 9.95% (6.41%-15.43%) of viremic pre-treatment and treatment-experienced PLHIV harbored the inT97A mutation, no major dolutegravir resistance mutations were observed.

**Interpretation:** Despite rising NNRTI resistance among pre-treatment PLHIV, overall population prevalence of pre-treatment resistance decreased due to treatment uptake. Most NNRTI and NRTI resistance is now contributed by treatment-experienced PLHIV. The high prevalence of mutations conferring resistance to components of current first-line ART regimens among PLHIV with viremia is potentially concerning.

**Funding:** National Institutes of Health and the Gates Foundation

**Research in context:** *Evidence before the study:* We searched PubMed for studies matching the keywords “hiv” “resistance” “longitudinal” “cohort” “population” published since 2004 (the beginning of antiretroviral therapy (ART) availability in sub-Saharan Africa) and identified 50 studies. We excluded 34 studies not based in sub-Saharan Africa, five studies primarily concerned with infection with other pathogens (e.g. HBV, *M. tuberculosis*), two studies concerned with insulin resistance, one sequencing-methods paper, and one paper concerned with host susceptibility to HIV infection. The remaining seven studies were not population-based meaning that the study population was not all persons but e.g. people living with HIV enrolled in care at a given clinic. Population-based cohort are essential for monitoring HIV drug resistance in both treated and untreated individuals, including those people who may go undetected in clinical settings, capturing evolutionary dynamics of resistance in real-world conditions.

*Added value of this study:* We estimated the prevalence of drug resistance over five consecutive survey rounds of a population-based open-cohort study in southern Uganda between 2012 and 2019 during a period of intense treatment scale-up. We show that among the entire population regardless of HIV status, 0.8% and 0.5% of individuals harbor viremic resistance to non-nucleoside reverse transcriptase inhibitors (NNRTIs) and nucleoside-reverse transcriptase inhibitors (NRTIs), respectively, of which the majority is dual-class NNRTI/NRTI resistance. Despite a two-fold increase in the prevalence of NNRTI resistance among pre-treatment viremic PLHIV, the overall prevalence of pre-treatment viremic resistance in the entire population decreased by more than 50% due to increased treatment initiation and population viral load suppression. The majority of resistance in recent survey rounds was contributed by treatment-experienced PLHIV. Among treatment-experienced viremic PLHIV, we observe a substantial burden of mutations that confer resistance to the NNRTI and NRTI components of dolutegravir and cabotegravir based regimens e.g. rtM184V (34%) rtY181C (15%), rtG190A (12%), rtK65R (12%), and rtK101E (9.5%). The integrase strand transfer inhibitor (INSTI) resistance mutation inT97A was observed in about a tenth of viremic PLHIV. These results provide the first longitudinal population-based estimates of temporal trends in the prevalence of drug resistance during ART program expansion in a high-burden setting. Further, they provide critical insight into the landscape of prevalent drug resistance substitutions circulating in this population.

*Implications of all the available evidence:* Scale-up of HIV treatment has increased the prevalence of drug resistance mutations among viremic people living with HIV in sub-Saharan Africa. The relatively high prevalence of NNRTI resistance has prompted a recent shift to first-line regimens including dolutegravir (an INSTI) in combination with NRTIs. The high prevalence of mutations conferring resistance to components of current first-line regimens in our population warrants continued monitoring of treatment failures and the prevalence of drug resistance in high burden settings.

## Introduction

Antiretroviral therapy (ART) suppresses human immunodeficiency virus (HIV) replication in persons living with HIV (PLHIV),^1^ which slows disease progression^2^ and prevents viral transmission.^3^ With increased uptake of ART as well as other interventions such as voluntary medical male circumcision, HIV incidence has fallen by nearly 40% globally since 2010.^4^

Viral resistance to ART threatens the clinical and public health impact of treatment scale-up^5,6^. Drug resistance can be acquired when an individual infected with a susceptible virus develops resistance following treatment. This is more common when treatment adherence is intermittent,^7^ but can occur despite high adherence.^8^ Throughout sub-Saharan Africa, the epicenter of the global HIV epidemic^4^, the majority of patients who remain viremic despite being on treatment with first-line regimens harbor resistance to at least one component of that regimen^1^. Viral genomes with resistance-conferring mutations can be transmitted to HIV seronegative individuals, increasing the risk of first-line treatment failure approximately five-fold.^9^

Through 2018, preferred first-line HIV ART regimens relied on a combination of nucleoside reverse transcriptase inhibitors (NRTIs) and non-nucleoside reverse transcriptase inhibitor (NNRTI)^10^. During the scale-up of ART, an increase in the prevalence of NNRTI resistance among pre-treatment PLHIV has been observed globally^11^. A systematic meta-regression estimated a 1.3% annual increase in NNRTI resistance in eastern Africa, reaching 10% by 2016. These findings have been corroborated by recent cross-sectional studies and World Health Organization (WHO) surveys^1,12,13^ and prompted a shift in recommendations to dolutegravir (DTG, an integrase strand transfer inhibitor [INSTI]) given in combination with NRTIs (e.g. Tenofovir disoproxil fumarate [TDF] and Lamivudine [3TC]) for first-line ART. Further, long-lasting injectable INSTIs (e.g. cabotegravir [CAB]) in combination with an NNRTI (rilpivirine [RPV]) are currently being rolled out throughout sub-Saharan Africa^14^.

The vast majority of data on the prevalence of HIV ART resistance in sub-Saharan Africa is derived from the population of PLHIV who report to healthcare clinics or hospitals^1,11–13^. Clinic-based studies are subject to biases in the population of PLHIV who are engaged and retained in care, which is not universal^15^. Specifically in eastern and southern Africa, only 95% of women and 91% of men are aware of their HIV status and of those only 92% and 86% are on ART, respectively ^4^. Further, clinic-based studies are able to estimate only the prevalence of ART resistance among PLHIV and not among the general population. The latter is the epidemiologically relevant parameter to inform the risk of exposure to HIV with ART resistance among seronegative individuals^16^.

General population-based studies, in which all individuals, regardless of HIV serostatus, are recruited to participate, can address these shortcomings. This design also allows for the accurate estimation of the overall prevalence of resistance and the relative contributions from different groups, such as pre-treatment and treatment-experienced PLHIV. A recent cross-sectional population-based study of drug resistance in KwaZulu-Natal, South Africa found very low (<1%) levels of resistance to INSTIs prior to DTG roll-out, but observed rtM184V (3TC resistance), rtK65R (TDF resistance), and rtK70E (TDF resistance) in 32.6%, 12.0%, and 6.2% of treatment-experienced PLHIV. Further, the rtE138A mutation, which confers resistance to RPV, was observed in in 6.5% and 7.9% of ART-experienced and -naïve PLHIV, respectively, in this setting^17^. Cross-sectional studies, however, are unable to assess temporal trends in the prevalence of resistance among PLHIV and the general population and do not capture the overall decrease in the prevalence of viremic HIV during ART scale-up. Conversely, longitudinal population-based cohort designs enable more precise monitoring of resistance evolution and a dynamic evaluation of the risks posed to current and future ART regimens. This study design is particularly useful in the context of rapidly changing population sizes of pre-treatment and treatment-experienced viremic PLHIV as has been observed globally in recent decades during expansion of treatment and prevention programs^15^.

Here, we analyzed HIV deep-sequence data collected from 3,407 PLHIV as part of a general population-based open cohort study in southern Uganda spanning a nine-year period of intense ART scale-up and declines in HIV incidence^18–20^. Our validated deep-sequencing protocol^21^ allows for the identification and quantification of drug resistance mutations present in a minority of the viral population within a given PLHIV, which can be selected for upon treatment initiation but are missed by consensus sequencing methods^22–24^. We estimated the prevalence of NNRTI, NRTI, and protease inhibitor (PI) resistance among the entire study population and among PLHIV as well as the temporal dynamics of viral resistance-conferring mutations among pre-treatment and treatment-experienced PLHIV.

## Methods

### Study design and participant selection

The Rakai Community Cohort Study (RCCS) is an open population-based census and cohort study conducted at approximately 18-24 month intervals (appendix pp 2-3) in agrarian (HIV prevalence 9-26%^25^), semi-urban trading (11-21%^25^), and Lake Victoria fishing (38-43%^25^) communities in southern Uganda.^20^ At each survey round, households in participating communities are censused and residents aged 15-49 capable of providing informed written consent (or assent if under 18) are invited to participate. Consenting participants are administered a structured questionnaire that obtains sociodemographic, behavioral, and health information, including self-reported past and current ART use. Voluntary HIV testing of participants is conducted using a rapid test algorithm^26^ and venous blood samples are taken for viral quantification and sequencing.

The RCCS is administered by the Rakai Health Sciences Program (RHSP) and has received ethical approval from the Uganda Virus Research Institute’s Research and Ethics Committee (HS540), the Uganda National Council for Science and Technology (GC/127/08/12/137), and the Johns Hopkins School of Medicine (IRB00217467). Participants provided written informed consent at each survey round.

We used survey data from 19 RCCS surveys conducted between November 5, 1994 and November 4, 2020, and HIV viral load and sequence data from five rounds conducted between August 10, 2011 and November 4, 2020. Participants with serologically confirmed HIV infection were considered pre-treatment during a given round if they reported never having taken ARTs at that round and all prior rounds in which they participated. PLHIV were considered treatment-experienced during a given round if they reported using ART at that round or any earlier rounds. Recommended first-line ART regimens in RCCS communities are presented in appendix p 4. Herein, study rounds were referred to by the year of the median interview date (appendix p 2). Reporting of this study adheres to the STROBE guidance^27^.

### HIV viral load quantification

HIV viral load was measured on serum/plasma samples using the Abbott real-time m2000 assay (Abbott Laboratories) at the Rakai health Sciences Program (Kalisizo, Uganda). Viral load measurements were conducted primarily among PLHIV in fishing communities in the 2012 survey round and for all PLHIV in later survey rounds. Viral loads ≥ 1000 copies/mL were considered viremic. Pre-treatment PLHIV in the 2012 round with missing viral load were imputed (appendix pp 4-5). In the rare instance where viral load measurements were missing for PLHIV in subsequent survey rounds, these observations were dropped from the analysis.

### HIV deep sequencing

Full-length HIV deep sequencing was conducted through the Phylogenetics and Networks for Generalized HIV Epidemics in Africa consortium (PANGEA-HIV).^28,29^ As described elsewhere,^19^ for the 2012 and 2014 surveys sequencing on Illumina MiSeq and HiSeq platforms using an amplicon-based approach^30^ was attempted for participants who self-reported never having been on ART and either had a missing viral load or were known to be viremic (appendix p 21). All viremic participant-visits, regardless of treatment status, in the 2015 through 2019 survey rounds, as well as select 2012 and 2014 participant-visits, were sequenced using the veSEQ-HIV protocol, which involves oligo-nucleotide bait enrichment of HIV from pooled metagenomic libraries prepared without virus-specific PCR.^31^ Our high-throughput implementation of the veSEQ-HIV protocol incorporates quantitative positive controls consisting of a serial dilution HXB2 cultured virus diluted in pooled human plasma from donors testing negative for HIV, as well and negative plasma controls included with every batch of samples processed. Sequencing quality was monitored from total counts of HIV reads detected in the quantitative controls, their PCR duplication rates and median insert sizes. Where contaminations were detected either by the presence of HIV reads in the negative control, or HXB2 reads present in the samples, sequencing runs were repeated. Sequencing for the 2019 round was only conducted for samples collected through May 17, 2019 (N = 171/453, 37.7% of viremic participant visits). Consensus sequences were generated using shiver^32^ and subtyped by identifying the most similar reference sequence and using the Recombination Identification Program.^33^

### Identification of drug resistance mutations

A validated bioinformatic pipeline, drmSEQ, was used to identify amino acid substitutions associated with reduced susceptibility to ART and predict individual drug and drug class susceptibilities at the University of Oxford Nuffield Department of Medicine.^34^ Paired-end reads were trimmed of adapters, primers, and low-quality bases with trimmomatic^35^ and then filtered to remove *pol* hypermutated sequences and non-HIV *pol* sequences. Duplicate reads introduced by PCR were removed using Picard MarkDuplicates^36^ and unique reads were locally aligned to 142 HIV subtype references using blastx.^37^ A manually-curated codon-restricted multiple-alignment of the references was used to lookup coordinates and mutations relative to HXB2 (GenBank: K03455.1). Only mutations supported by a minimum of 10 PCR-deduplicated reads and by ≥5% of reads spanning the corresponding site were considered.^38^ These thresholds are based on HIV read counts after removal of non-unique PCR duplicate reads. Importantly, the same thresholds were used in a previous validation and demonstrated comparable sensitivity to a gold standard clinical assay^34^. Amino acid substitutions were scored according to the Stanford University HIV Drug Resistance Database. Scores were summed to predict susceptibility to 25 HIV drugs (appendix p 13).^39–41^ A score ≥30 (intermediate/high-level) for a given drug was categorized as resistant. Resistance was not predicted if less than half of the relevant positions for a given drug had fewer than 10 reads. Samples in which there was insufficient sequencing coverage for one or more drug within a class were not assigned a resistance categorization for that class. Samples with resistance to at least one drug within each class were categorized as resistant to that class.

### Outcome measures

The primary outcomes of this study were the prevalence of viremic PLHIV with INSTI, NNRTI, NRTI, and or PI resistance among all participants, regardless of HIV serostatus, in each survey round. We also estimated the population prevalence of NNRTI, NRTI, and PI resistance contributed by viremic pre-treatment and treatment-experienced PLHIV and the population prevalence of multi-class resistance. We further estimated the prevalence of NNRTI, NRTI, and PI resistance and individual resistance-conferring viral mutations specifically among viremic pre-treatment and treatment-experienced PLHIV in each survey round. Given the greater prevalence of viremic HIV in fishing communities, among men, and among younger age groups in the RCCS^19,20,25^ we evaluated the association between these variables and resistance in bivariate analyses. Stratified estimates were generated for covariates identified as significant in bivariate analyses. Given the availability of sequence data, we restricted prevalence estimates that include treatment-experienced PLHIV to the 2015 and 2017 survey rounds and use the 2017 survey as an end-point for pre-treatment PLHIV. For context, we estimated the prevalence of PLHIV, viremic PLHIV (2014 and later due to missing viral load data), viremic PLHIV pre-treatment, and viremic treatment-experienced PLHIV among participants in each round (2014 and later).

### Statistical methods

Statistical analyses were conducted in R v.4.4.1.^42^ Prevalence was estimated using Poisson regression with a log-link and robust (sandwich) standard errors^43^ which were fit with general estimating equations using geepack v.1.3.11 to account for repeated measures^44^. Correlation structures were chosen by minimizing the Quasi Information Criterion (QIC). We used inverse probability weighting to account for missing sequence data among viremic study participants. Sampling weights were calculated based on availability of a viral load measurement (True/False), log_10_ copies/mL where available, community type (agrarian/fishing/trading), age category ((14,24]/(24,34]/(34,49]), and sex (M/F) stratified by survey round. Emmeans v. 1.10.4 was used to calculate prevalence within strata^45^. 95% confidence intervals and *p-*values (a = 0.05) were calculated using the Wald method. *x^2^ p-*values were calculated using the stats package in R. Data analysis and visualization was done using tidyverse v.2.0.0,^46^ ggplot2 v.3.5.1,^47^ cowplot v.1.1.3,^48^ patchwork v. 1.2.0^49^, and ggpattern v.1.1.1^50^. Readxl v.1.4.3^51^ and haven v.2.5.4.9^52^ were used to parse data files. See appendix pp 5-17 for detailed methods.

## Results

### Study population

Between August 10, 2011 and November 4, 2020, a total of 43,361 people participated in the RCCS, of whom 7,923 (18.27%) were PLHIV. Of 93,622 participant-visits about a fifth were from PLHIV (table 1). Over the analysis period, the median age of study participants remained stable whereas the age of PLHIV increased slightly (appendix pp 18-19). Viral load measurements were available for 1,959/3,498 (56.00%) of PLHIV in the 2012 survey and 13,962/14,008 (99.67%) PLHIV in later survey rounds. A total of 46 participant-visits from the 2014-2019 surveys with missing viral loads were dropped from subsequent analyses. Among participant-visits from PLHIV in 2014-2019, 26.29% were contributed by viremic PLHIV and of those 79.81% were pre-treatment viremic. After imputation of missing viral loads (*Methods*), 56.75% of PLHIV in the 2012 survey were identified as pre-treatment viremic.

HIV seroprevalence among participants decreased from 20.38% (95% CI 19.78% - 20.99%) in the 2012 survey to 17.20% (95% CI 16.68% - 17.74%) in the 2019 survey (figure 1A, appendix p 20). Concurrent with an increase in the proportion of PLHIV reporting ever having been on treatment from 25.04% (2012) to 85.25% (2019, appendix p 21), HIV viremia among all participants decreased significantly from 8.14% (2014, 95% CI 7.75% - 8.55%) to 2.34% (2019, 95% CI 2.14% - 2.57%). These declines were driven by a nearly nine-fold (prevalence ratio (PR) 0.13, 95% CI 0.11 – 0.15) decrease in the prevalence of pre-treatment viremia among participants over the study (figure 1A, appendix p 20). The prevalence of treatment-experienced viremia remained stable at around 1% of participants.

**Figure 1:**
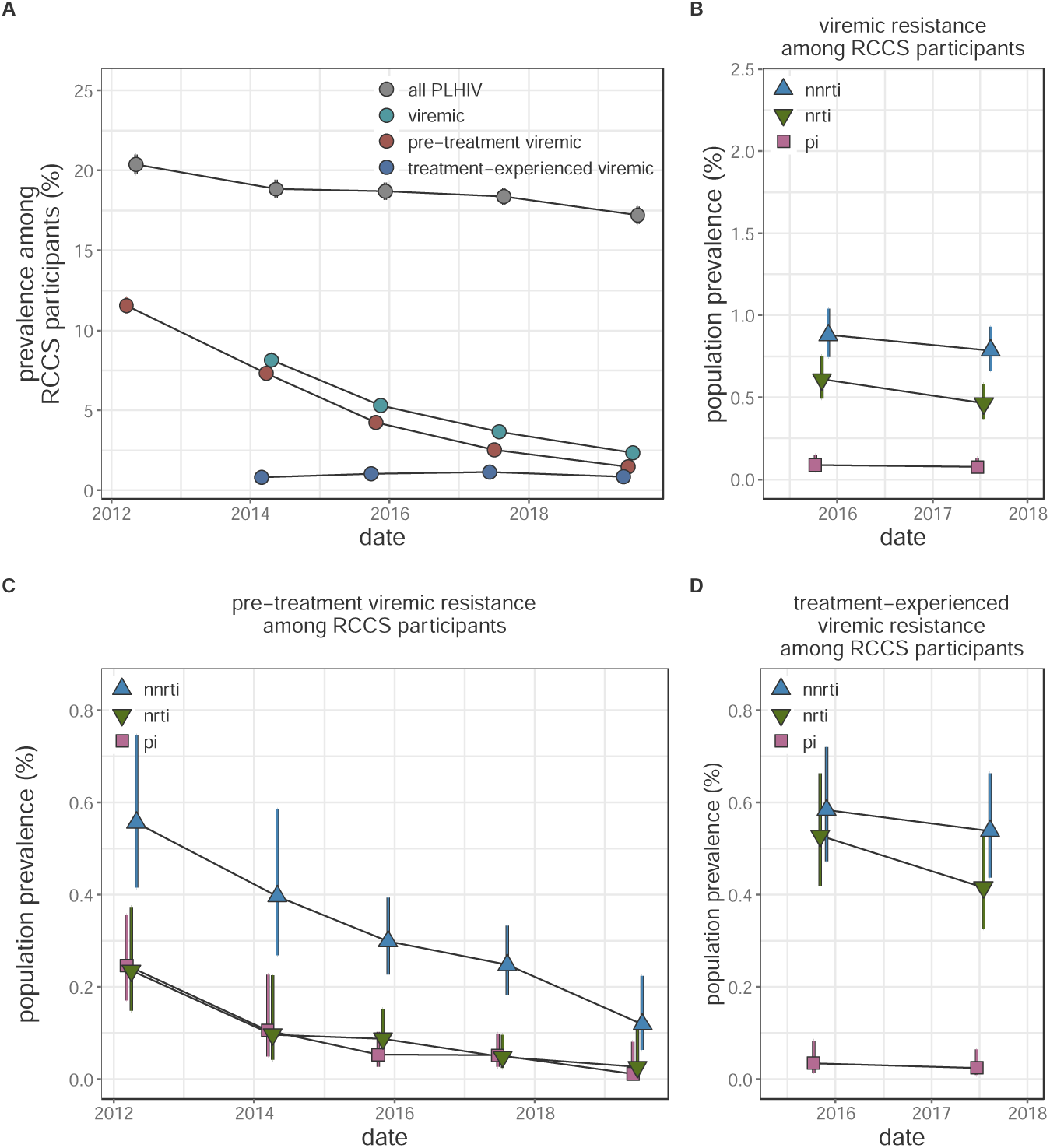
Longitudinal trends in HIV seroprevalence and population prevalence of viremic HIV drug resistance among Rakai Community Cohort Study participants, 2012-2019. (A) Estimated prevalence of all HIV, viremic HIV, viremic pre-treatment HIV, and viremic treatment-experienced HIV in each round. Due to missing viral load data, prevalence of viremic HIV and viremic treatment-experienced HIV were not estimated in the 2012 survey. For some estimates confidence bands do not extend beyond point. (B-D) Estimated population prevalence of all viremic (B), pre-treatment viremic (C), and treatment-experienced viremic (D) NNRTI, NRTI, and PI resistance among all study participants. Estimates were generated using Poisson regression with robust standard errors with survey round as a predictor variable. Generalized estimating equations with correlation structure selection by Quasi Information Criterion value (A: independent, B: independent, C: independent, D: exchangeable (NNRTI and PI), independent (NRTI)) were used to account for repeat participants across study rounds. Error bars indicate the Wald 95% confidence interval for the mean value. For clarity, points are jittered along the x-axis. PLHIV = people living with HIV. NNRTI = non-nucleoside reverse transcriptase inhibitors (blue upwards facing triangles). NRTI = nucleoside reverse transcriptase inhibitors (green downwards facing triangles). PI = integrase inhibitors (pink squares).

### Identification of resistance genotypes in deep-sequence data

Deep-sequence based identification of drug resistance mutations (DRMs) was attempted on 4,525/5,724 (79.51%) of viremic participant visits (appendix p 22). Attempted genotyping did not vary by participant age, sex, or community type of residence. The veSeq-HIV sequencing protocol was used for 44.99% of all sequenced viremic participants and the vast majority (99.37%) of those in the 2015 through 2019 survey rounds (appendix p 23). Among samples from viremic participant-visits on which deep-sequence based genotyping was attempted, sufficient data were available to reliably genotype 4,072/4,525 (90.01%, appendix pp 24-25) viruses from 3,407 PLHIV for at least one drug. Sequencing success did not depend on age, community, type or sex, (*p*-values ≥ 0.37) but was more likely among samples with higher viral load and those sequenced with veSeq-HIV (*p-*values = 0.0005). Among sequenced participant-visits successfully genotyped for all INSTIs (*n*=2,578, appendix pp 26-32), NNRTIs (*n*=3,050), NRTIs (*n*=3,009) or PIs (*n*=3,520) <1%, 12.46%, 6.75%, and 1.88% had predicted resistance, respectively (appendix pp 33-34). Given the minimal INSTI resistance we did not estimate the prevalence of INSTI resistance.

### Population prevalence of viremic resistance

In 2017, the population prevalence of viremic NNRTI, NRTI, and PI resistance among all participants, regardless of HIV serostatus, was 0.79% (95% CI 0.66% - 0.93%), 0.46% (95% CI 0.37% - 0.58%), and 0.08% (95% CI 0.04% - 0.13%), respectively. These levels were stable compared to 2015 (figure 1B, appendix p 35). In stratified analyses, NNRTI and NRTI resistance was more than three-times as common in fishing as compared to agrarian or trading communities and most prevalent among people aged 25-34 years old (*p*-values ≤ 0.0001, appendix pp 36-38).

The prevalence of NNRTI and NRTI resistance contributed by pre-treatment viremic PLHIV decreased 2.3-fold (PR 0.44, 95% CI 0.29 – 0.68) and 5-fold (PR 0.21, 95% CI 0.09 – 0.47) between the 2012 and 2017 surveys (*p-*values < 0.0001, figure 1C, appendix p 38), concurrent with the observed decline in population prevalence of pre-treatment viremia. Consequently, in the 2017 survey round, treatment-experienced viremic PLHIV contributed 68.46% (95% CI 59.49% - 75.44%) and 89.61% (79.92% - 94.62%) of all NNRTI and NRTI resistance, respectively. Specifically, the population prevalence of resistance to NNRTIs and NRTIs contributed by treatment-experienced viremic PLHIV in the 2017 survey was 0.54%, 95% CI 0.44%-0.66% and 0.42%, 0.33%-0.53% as compared to 0.25%, 0.18%-0.32% and 0.05%, 0.02%-0.1% (appendix pp 39 – 40) contributed by pre-treatment viremic PLHIV.

Resistance profiles varied considerably by treatment status (figure 2 and appendix p 41). Among pre-treatment viremic PLHIV with available genotype for NNRTIs, NRTI, and PIs the majority with any resistance were NNRTI mono-resistant (2017 survey *n*=35, 70%). In contrast, among treatment-experienced viremic PLHIV with any resistance, NNRTI/NRTI dual-class resistance was the most common profile (2017 survey *n*=64, 73.56%). Among all participants, the most common forms of viremic resistance were NNRTI/NRTI dual-class resistance (2017: 0.44%, 95% CI 0.35% - 0.56%) and NNRTI mono-resistance (2017: 0.32%, 95% CI 0.25% - 0.42%), consistent with a dominant contribution from treatment-experienced viremic PLHIV. Other resistance profiles were extremely rare (<0.1%).

**Figure 2:**
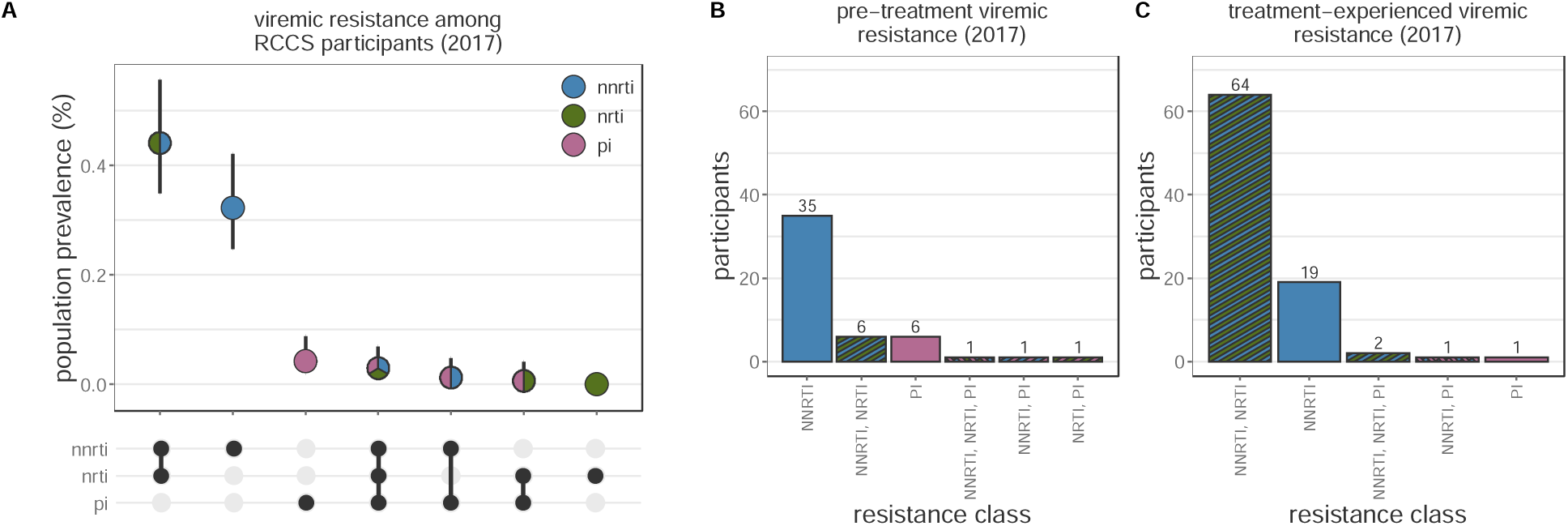
Patterns of multi-class resistance in Rakai Community Cohort Study, 2017. (A) Estimating population prevalence of NNRTI, NRTI, and PI mono-resistance and NNRTI/NRTI, NNRTI/PI, NRTI/PI, and NNRTI/NRTI/PI multi-class resistance among all RCCS study participants. Estimates were generated using Poisson regression with robust standard errors with survey round as a predictor variable. General estimating equations with the best fit correlation structure by QIC value (NNRTI, NRTI, PI mono-resistance and NNRTI/NRTI and NNRTI/PI multi-class resistance: independent, NRTI/PI and NNRTI/NRTI/PI: exchangeable) were used to account for repeated measures from the same participant Error bars indicate the Wald 95% confidence interval for the mean value. (B) Multi-class resistance profiles among 50 pre-treatment viremic 2017 participant-visits with genotype data for all NNRTIs, NRTIs, PIs, and resistance to at least one of these drug classes. (C) Multi-class resistance profiles among 87 treatment-experienced viremic 2017 participant-visits with genotype data for all NNRTIs, NRTIs, PIs, and resistance to at least one of these drug classes. NNRTI = non-nucleoside reverse transcriptase inhibitors. NRTI = nucleoside reverse transcriptase inhibitors. PI = integrase inhibitors.

### Prevalence of resistance among pre-treatment viremic PLHIV

Between the 2012 and 2017, NNRTI resistance among pre-treatment viremic PLHIV increased by a factor of 1.98 (95% CI: 1.34-2.91), reaching 9.61% (95% CI: 7.27% - 12.7%) (figure 1A and appendix pp 42). This increasing trend did not vary by sex, age, type of community of residence, or sequencing approach (appendix pp 43-44).The prevalence of NRTI and PI resistance remained stable and below 2.1% over the same time period.

Among pre-treatment viremic PLHIV, the most prevalent resistance-associated mutation was inT97A (figure 3B and appendix pp 46-48), an INSTI-resistance (particularly Elvitegravir^53^) mutation, detected in ∼10% of pre-treatment viremic participants in the 2012 through 2017 survey rounds and in 20% (95% CI 13.89% - 28.8%) in the partial 2019 survey data. The most common NNRTI-resistance mutation was rtK103N, found in 7.1% (95% CI 5.05% - 9.97%) of pre-treatment viremic PLHIV in the 2017 survey, a 4.26-fold (95% CI 2.11 - 8.59) increase compared to the 2012 survey. The next most prevalent NNRTI mutation, rtE138A, which is associated with 2.5-fold reduced susceptibility to RPV^54^, was present in only 2.77% (95% CI 1.59% – 4.85%) of pre-treatment viremic PLHIV and its prevalence remained stable compared to the 2012 survey (*p-*value = 0.49). NRTI resistance mutations were rare compared to NNRTI mutations. Genotypes associated with intermediate/high-level INSTI resistance were identified in 16 pre-treatment viremic participant-visits (<1%), the majority of which harbored inE92G (*n*=13), which confers resistance to Elvitegravir, a drug that is not routinely used in Uganda (appendix p 4). Mutations conferring intermediate/high-level resistance to DTG were not observed.

**Figure 3:**
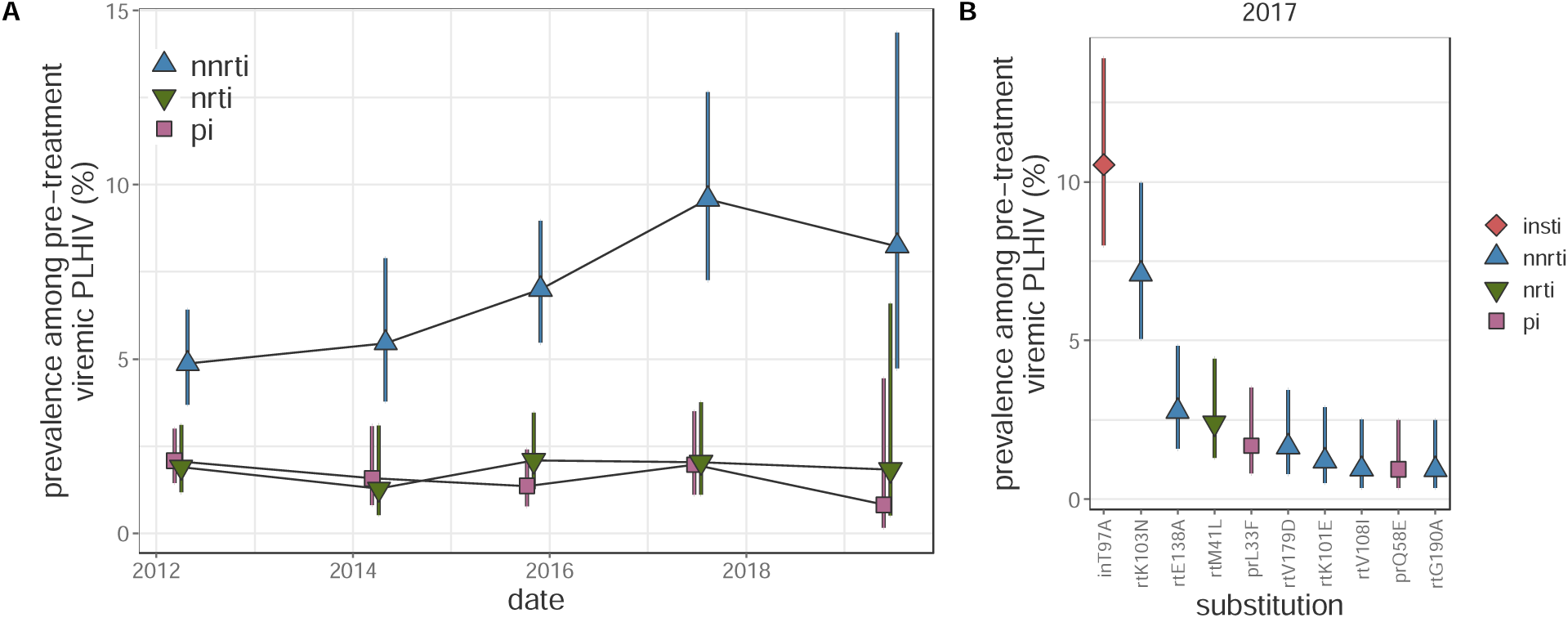
Longitudinal trends in HIV drug resistance among pre-treatment viremic Rakai Community Cohort Study participants, 2012-2019. (A) Estimated prevalence of NNRTI, NRTI, and PI resistance among pre-treatment viremic PLHIV. For visual clarity, points are jittered along the x-axis. (B) Prevalence in the 2017 survey of the 10 most prevalent substitutions in pre-treatment viremic PLHIV sorted by prevalence. Estimates were generated using Poisson regression with robust standard errors with survey round as a predictor variable. General estimating equations with the best fit correlation structure by QIC value (NNRTI: exchangeable, NRTI: exchangeable, PI: AR1, substitutions: independent to ensure convergence) were used to account for repeated measures from the same participant Error bars indicate the Wald 95% confidence interval for the mean value within each category. PLHIV = people living with HIV. NNRTI = non-nucleoside reverse transcriptase inhibitors. NRTI = nucleoside reverse transcriptase inhibitors. PI = integrase inhibitors.

### Prevalence of resistance among treatment-experienced viremic PLHIV

Prevalence of NNRTI and NRTI resistance was substantially higher among treatment-experienced participants as compared to pre-treatment PLHIV. In 2017, 51.49% (95 CI 46.24%-57.34%) and 36.46% (95% CI 30.06%-44.22%) of treatment-experienced participants with viremia harbored NNRTI and NRTI resistant viruses, respectively (figure 4A and appendix p 49). NRTI resistance was 1.62 (95% CI 1.03 – 2.56, appendix pp 50-51) times more common among participants aged 25-34 years compared to 15-24 year-olds (*p*-value = 0.037). While NNRTI resistance remained stable between the 2015 and 2017 survey rounds, NRTI resistance decreased by more than quarter (prevalence ratio 0.73%, 95% CI 0.58 - 0.92, *p-*value = 0.0084). Only 2.13% (95% CI 0.81%-5.63%) of treatment-experienced viremic participants in the 2017 survey round had viruses with PI resistance.

**Figure 4:**
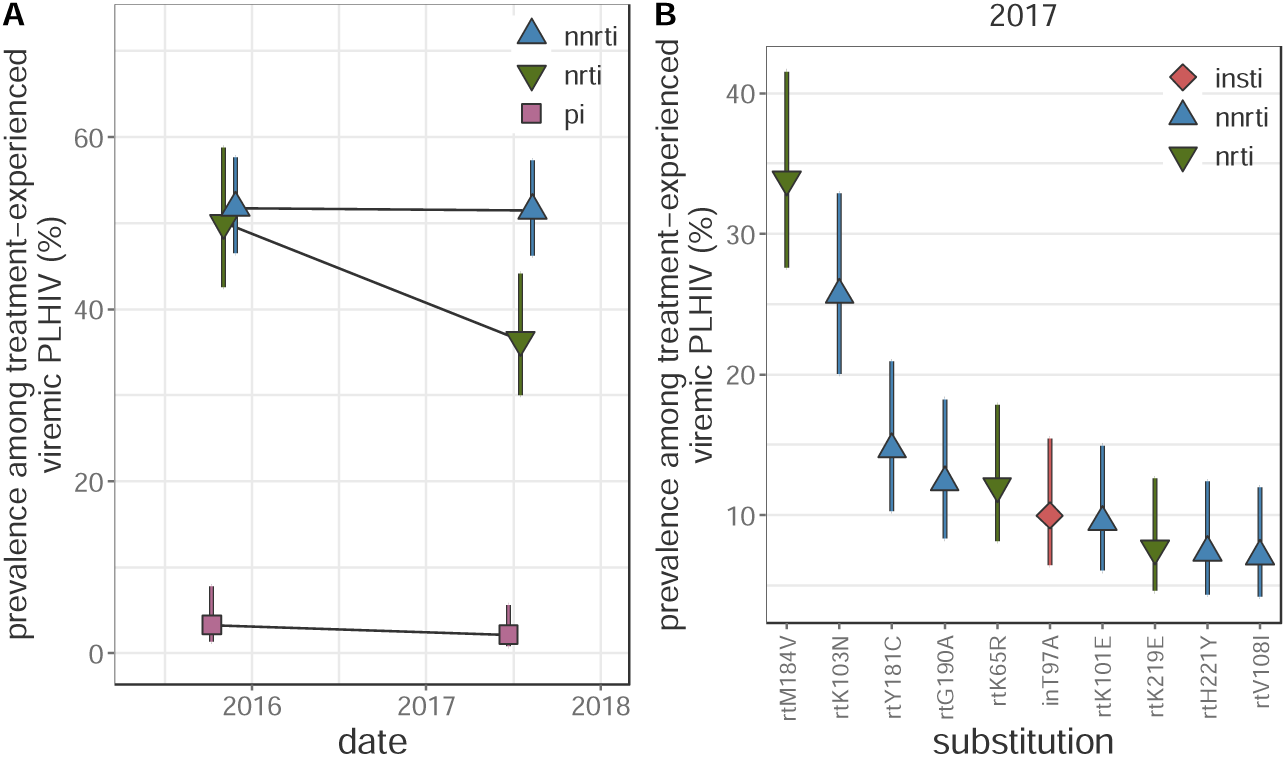
Longitudinal trends in HIV drug resistance among treatment-experienced viremic Rakai Community Cohort Study participants, 2015-2017. (A) Estimated prevalence of NNRTI, NRTI, and PI resistance among treatment-experienced viremic PLHIV. For visual clarity, points are jittered along the x-axis. (C) Prevalence of the 10 most prevalent drug resistance mutations in treatment-experienced viremic PLHIV in the 2017 survey round, sorted by prevalence. Estimates were generated using Poisson regression with robust standard errors with survey round as a predictor variable. General estimating equations with the best fit correlation structure by QIC value (NNRTI and PI: exchangeable, NRTI: independent, mutations: independent to ensure convergence) were used to account for repeated measures from the same participant Error bars indicate the Wald 95% confidence interval for the mean value within each category. PLHIV = people living with HIV. NNRTI = non-nucleoside reverse transcriptase inhibitors. NRTI = nucleoside reverse transcriptase inhibitors. PI = integrase inhibitors.

The resistance-associated mutations observed among treatment-experienced viremic participants differed considerably compared to pre-treatment viremic participants (appendix pp 46, 52). NRTI resistance among treatment-experienced viremic participants in the 2017 survey was most frequently due to the rtM184V (33.89%, 95% CI 27.61% - 41.59%), rtK65R (12.07% 95% CI 8.14% - 17.9%), and rtK219E (7.64%, 95% CI 4.6% - 12.67%) substitutions, which were rarely observed among pre-treatment PLHIV. On the contrary, the most prevalent NNRTI-associated substitution among treatment-experienced viremic participants was rtK103N (25.68%, 95% CI 20.03 - 32.92%), however rtY181C (14.67%, 95% CI 10.27% - 20.95%) and rtG190A (12.34%, 95% CI 8.33% - 18.27%) were also frequently observed. inT97A was observed at a similar prevalence as among pre-treatment viremic participants (9.96%, 95% CI 6.41 - 15.48%). Only four participant-visits contributed by viremic treatment-experienced PLHIV (<1%) harbored INSTI resistance mutations, which were each observed only once (inG163K, inG163R, inR263K, inS147G) and not associated with DTG resistance.

## Discussion

In this study, we report on trends in HIV drug resistance from a longitudinal, population-based cohort in southern Uganda between 2012 and 2019, a period marked by the substantial expansion of ART programs. Despite a doubling in the prevalence of NNRTI resistance among pre-treatment PLHIV, we observed an overall decline in the population prevalence of pre-treatment HIV drug-resistant viremia, alongside increasing ART uptake and viral suppression among PLHIV. By the end of the analysis period, the population prevalence of NNRTI resistance was 0.78% and NRTI resistance was 0.46%, with most resistance stemming from dual-class NNRTI/NRTI resistance in treatment-experienced viremic individuals. Notably, dual-class resistance remained relatively uncommon among pre-treatment viremic PLHIV, and resistance trends for NRTIs and PIs in this group remained stable throughout the ART scale-up, despite a substantial burden of NRTI resistance in treatment-experienced individuals. We also observed a relatively high background prevalence of the INSTI-resistance mutation inT97A. Overall, these findings provide important insights into the evolving dynamics of HIV drug resistance during ART scale-up in a high-burden East African population and may help guide future surveillance and HIV epidemic control efforts in the region.

Consistent with previous studies, we observed an increase in the prevalence of NNRTI resistance among viremic pre-treatment individuals living with HIV, supporting the recent shift to DTG-based regimens^1,11,12^. However, a key finding of this population-based analysis is that, concurrent with this rise in NNRTI resistance, we observed a substantial decline in the overall population prevalence of pre-treatment HIV, likely driven by both increased treatment initiation and declining HIV incidence^18–20^. Importantly, this decrease in pre-treatment HIV has outpaced the rise in NNRTI resistance, resulting in a more than 50% reduction in the population prevalence of pre-treatment HIV with NNRTI resistance over the study period. By the end of the survey period, most viremic individuals with NNRTI-resistant HIV were those with prior treatment experience.

We also find a lower burden of NNRTI and NRTI resistance among viremic treatment-experienced PLHIV in this study as compared to clinic-based studies^1,12^. This is likely because our population-based study design includes PLHIV who remain viremic because they are not actively engaged in care, despite past treatment exposure. In comparison, clinic-based studies may disproportionately enroll people who remain viremic due to sub-optimal adherence or are in more advanced stages of disease, and thus more likely to have drug resistance, whereas population-based sampling includes people lost to clinic-based care and no longer using treatment altogether.

Both NNRTI and NRTI therapies remain important components of the current and future ART landscape. Current first-line DTG-regimens incorporate two NRTIs (e.g. TDF and 3TC) and DTG treatment failure is more likely among those with NRTI resistance^55,56^. We here observe rtM184V (>1000-fold reduced susceptibility to 3TC^57^) and rtK65R (five-fold reduced susceptibility to TDF^58^) in 34% and 12% of viremic treatment-experienced PLHIV. In contrast, these mutations were observed only rarely among pre-treatment PLHIV. Further, we identify a number of mutations associated with reduced susceptibility to the RPV (e.g. rtK101E, rtE138A, rtY181C, and rtG190A), an NNRTI given in combination with CAB as part of long-lasting injectable therapies, in 10-12% of viremic treatment-experienced PLHIV.

As this study pre-dates the scale-up of DTG, we do not observe major DTG resistance-conferring mutations. Approximately 10% of viremic participants harbored inT97A, which is a polymorphic mutation most common in subtype A and in isolation confers two-fold resistance to EVG but not to other INSTIs^59^. The observed prevalence of inT97A in this study is an order of magnitude higher than in a population-based cohort in South Africa^17^ and about twice as prevalence as globally-sampled INSTI-naïve PLHIV^59^. Further, we observe a significant increase in the prevalence of inT97A among pre-treatment viremic PLHIV in the 2019 survey round. As inT97A is repeatedly selected for in subjects failing DTG therapy^60^ and in combination with other mutations (e.g. inG140S and inQ148H) can significantly increase DTG resistance^61–63^, we recommend continued monitoring.

There are important limitations of this work. Due to unknown HIV serostatus among non-participating residents of RCCS communities, we did not generalize our results beyond study participants. Younger individuals, men, and residents of trading communities are less likely to participate in RCCS surveys.^20^ Further, only self-reported treatment status was available, which may have led to the misclassification of some participants. Prior work in this cohort demonstrated that 11% of self-reported ART-naïve participants had antivirals in their blood.^64^ Given the significant differences observed in the mutational profiles of pre-treatment as compared to treatment-experienced PLHIV and the consistency of these results with estimates of the fitness impact of mutations in the absence of treatment,^65^ we expect minimal misclassification bias. While we lack data on individual-level ART regimens, first-line therapy in this setting is highly consistent across individuals. As this work is based on sequencing of viral RNA, we could only identify resistance among viremic PLHIV. Consequently, our population prevalence estimates are an underestimate as some PLHIV with resistance may be suppressed through second-line therapy or were transiently suppressed following treatment initiation. The latter may be more pronounced in recent survey rounds as treatment scale-up has increased the proportion of recent treatment initiators.

Despite the population-based study design, viral load data and sequence data was available for only a subset of participants due to budgetary and logistical constraints. We consequently restrict analyses to survey rounds where sufficient data is available to generate reliable inferences and use imputation to account for missing viral load data. Despite this missingness, deep-sequence data was available for 4,072 participant-visits, which is considerably more than a recent population-based study in South Africa (*n*=1,097),^17^ clinic-based studies in sub-Saharan Africa (*n*=972),^12^ and WHO surveys in Uganda (*n*=372)^1^. Further, we utilized detailed demographic data on survey participants to account for the role of potential biases in sequence data availability. However, we cannot rule out potential residual biases in our estimates.

In summary, this study adds critical context to our understanding of the HIV epidemic in southern Uganda and to the impact of treatment expansion on the population burden of HIV resistance. We show that following ART scale-up, most resistance is contributed by treatment-experienced PLHIV, which may inform interventions aimed at reducing transmitted HIV resistance. The relatively high prevalence of NRTI and NRTI-resistance among treatment-experienced PLHIV and of inT97A among all viremic PLHIV is concerning in light of the roll-out of DTG+ TDF+3TC and CAB+RPV regimens in sub-Saharan Africa. Overall, these findings stress the importance of continued viral sequence-based monitoring of resistance mutations among PLHIV, particularly those with previous treatment exposure, during the roll-out of novel HIV ART regimens.

## Data Availability

Code to reproduce all analyses and visualizations as well as de-identified resistance and limited patient metadata are available at https://github.com/m-a-martin/rccs_hiv_resistance_r15_r19.

## Acknowledgements

We thank the Rakai Health Sciences Program and the participants of the Rakai Community Cohort Study for making this research possible. We thank members of the PANGEA-HIV consortium, the Johns Hopkins University Infectious Disease Dynamics group, and the NIAID Laboratory of Immunoregulation International HIV/STD Section at the Johns Hopkins University School of Medicine for helpful feedback on this work. This work was supported by the National Institutes of Health (NIH) National Institute of Allergy and Infectious Diseases (NIAID) (U01AI075115, R01AI087409, U01AI100031, R01AI110324, R01AI114438, K25AI114461, R01AI123002, K01AI125086, R01AI128779, R01AI143333, R21AI145682, R01AI155080, ZIAAI001040), NIH National Institute of Child Health and Development (R01HD050180, R01HD070769, R01HD091003), NIH National Heart, Lung, and Blood Institute (R01HL152813), the Fogarty International Center (D43TW009578, D43TW010557), the Johns Hopkins University Center for AIDS Research (P30AI094189), the Bill & Melinda Gates Foundation (OPP1084362, INV-007573, INV-035619, INV-060259, INV-075093), the U.S. President’s Emergency Plan for AIDS Relief through the Centers for Disease Control and Prevention (NU2GGH000817), and in part by the Division of Intramural Research, NIAID, NIH.

## Data sharing

Code to reproduce all analyses and visualizations as well as de-identified resistance and limited patient metadata are available at https://github.com/m-a-martin/rccs_hiv_resistance_r15_r19. Due to privacy concerns we are unable to share individual-level data on community of residence.

## Contributors

M.A.M.: Conceptualization, data curation, formal analysis, methodology, project administration, validation, visualization, writing – original draft, writing – review & editing

S.J.R.: Conceptualization, investigation, funding acquisition, resources, writing – review & editing

C.S.: Investigation, writing – review & editing

B.T.F.: Data curation, investigation, formal analysis, software, writing – review & editing

F.N.: Investigation, writing – review & editing

T.C.Q.: Conceptualization, funding acquisition, investigation, resources, writing – review & editing

S.A.K.: Conceptualization, writing – review & editing M.N.: Investigation, writing – review & editing

E.N.K.: Investigation, writing – review & editing G.K.: Investigation, writing – review & editing

R.S.: Investigation, writing – review & editing R.K.G.: Conceptualization, writing – review & editing

L.A.D: Funding acquisition, resources, writing – review & editing

J.K.: Conceptualization, investigation, funding acquisition, resources, writing – review & editing

O.R.: Conceptualization, funding acquisition, investigation, writing – review & editing

C.F.: Conceptualization, funding acquisition, investigation, writing – review & editing

R.M.G.: Conceptualization, investigation, writing – review & editing

D.B.: Conceptualization, data curation, formal analysis, funding acquisition, investigation, methodology, project administration, resources, software, supervision, writing – review & editing

M.K.G.: Conceptualization, data curation, formal analysis, funding acquisition, investigation, project administration, resources, supervision, writing – original draft, writing – review & editing

