## Appendix for "Population dynamics of HIV drug resistance during treatment scale-up in Uganda: a population-based longitudinal study"

**Table of Contents**

P1…Table of contents

P2…Supplementary Table 1: Rakai Community Cohort Study interview date ranges

P3…Supplementary Figure 1: Rakai Community Cohort Study interview dates

P4…Supplementary Table 2: Recommended ART regimen among RCCS participants by year

P5-11…Supplementary methods

P12-13…Supplementary Table 3: Summary of outcome measures

P14…Supplementary Table 4: Drugs to which resistance was predicted

P15-17…Supplementary Table 5: Sampling probability model for the probability that genotyping data is available among all viremic PLHIV

P18…Supplementary Table 6: Demographics of study participants by survey round

P19…Supplementary Table 7: Demographics of participants living with HIV by survey round

P20…Supplementary Table 8: Prevalence of PLHIV, viremic PLHIV, viremic pre-treatment PLHIV, and viremic treatment-experienced PLHIV

P21…Supplementary Table 9: Number of participant-visits contributed by PLHIV with history of treatment

P22…Supplementary Table 10: Number of viremic participant-visits from which deep-sequence based identification of drug resistance mutations was attempted.

P23…Supplementary Table 11: Technology used to generate sequence data by survey round

P24…Supplementary Table 12: Number of viremic participant-visits with attempted deep-sequence based identification of drug resistance mutations that had successful genotyping for at least one drug

P25… Supplementary Table 13: Deep-sequencing quality summary statistics among participant-visits with successful genotyping for at least one drug

P26…Supplementary Table 14: Number of viremic participant-visits with attempted deep-sequence based identification of drug resistance mutations that had successful genotyping for all INSTIs

P27…Supplementary Table 15: Number of viremic participant-visits with attempted deep-sequence based identification of drug resistance mutations that had successful genotyping for all NNRTIs

P28…Supplementary Table 16: Number of viremic participant-visits with attempted deep-sequence based identification of drug resistance mutations that had successful genotyping for all NRTIs

P29…Supplementary Table 17: Number of viremic participant-visits with attempted deep-sequence based identification of drug resistance mutations that had successful genotyping for all PIs

P30…Supplementary Table 18: Number of viremic participant-visits with attempted deep-sequence based identification of drug resistance mutations that had successful genotyping for all NRTIs, NNRTIs, and PIs

P31…Supplementary Table 19: Number of viremic participant-visits with attempted deep-sequence based identification of drug resistance mutations that had successful genotyping for all drugs

P32-33…Supplementary Table 20: Number of viremic participant-visits that had resistant genotypes for each drug class

P34…Supplementary Table 21: Population prevalence of viremic NNRTI, NRTI, and PI resistance by survey round

P35…Supplementary Table 22: Association between age, community type, and sex and the population prevalence of viremic NNRTI, NRTI and PI resistance adjusted by survey round

P36-37…Supplementary Table 23: Population prevalence of viremic NNRTI, NRTI, and PI resistance stratified by survey round and age, community type, and sex

P38…Supplementary Table 24: Population prevalence of pre-treatment viremic NNRTI, NRTI, and PI resistance by survey round

P39…Supplementary Table 25: Population prevalence of treatment-experienced viremic NNRTI, NRTI, and PI resistance by survey round

P40… Supplementary Table 26: Population prevalence of viremic multi-class resistance by survey round

P41…Supplementary Table 27: Prevalence of NNRTI, NRTI, and PI resistance among pre-treatment viremic PLHIV by survey round

P42-43…Supplementary Table 28: Association between age, community type, and sex and the prevalence of NNRTI, NRTI and PI resistance among pre-treatment viremic PLHIV adjusted by survey round

P44…Supplementary Table 29: Prevalence of NRTI resistance among pre-treatment viremic PLHIV stratified by sex

P45…Supplementary Table 30: Count of amino acid mutations observed among viremic PLHIV

P46-47…Supplementary Table 31: Prevalence of amino acid mutations among viremic pre-treatment PLHIV by survey round

P48…Supplementary Table 32: Prevalence of NNRTI, NRTI, and PI resistance among viremic treatment-experienced PLHIV by survey round

P49…Supplementary Table 33: Association between age, community type, and sex and the prevalence of NNRTI, NRTI and PI resistance among treatment-experienced viremic PLHIV adjusted by survey round

P50…Supplementary Table 34: Prevalence of NNRTI, NRTI, and PI resistance among treatment-experienced viremic PLHIV by survey round and age, community type, and sex

P51-52…Supplementary Table 35: Prevalence of amino acid mutations among treatment-experienced viremic PLHIV

P53…Supplementary references

**Supplementary Table 1: Rakai Community Cohort Study survey dates**

| **Round** | **Survey start date** | **Survey mid date** | **Survey end date** |
| --- | --- | --- | --- |
| 1 | 1994-11-05 | 1995-03-20 | 1995-07-18 |
| 2 | 1995-01-12 | 1996-01-10 | 1996-05-01 |
| 3 | 1996-01-12 | 1996-11-20 | 1997-04-23 |
| 4 | 1997-06-06 | 1997-11-21 | 1998-05-12 |
| 5 | 1997-08-05 | 1998-08-06 | 1999-03-15 |
| 6 | 1999-04-06 | 1999-09-07 | 2000-02-09 |
| 7 | 2000-03-20 | 2000-08-08 | 2001-02-19 |
| 8 | 2001-04-03 | 2001-10-23 | 2002-05-31 |
| 9 | 2002-07-15 | 2003-01-28 | 2003-08-01 |
| 10 | 2003-09-26 | 2004-04-23 | 2004-11-23 |
| 11 | 2005-01-13 | 2005-10-13 | 2006-06-30 |
| 12 | 2006-03-31 | 2007-06-28 | 2008-04-24 |
| 13 | 2008-06-17 | 2009-03-17 | 2009-12-04 |
| 14 | 2010-01-18 | 2010-09-24 | 2011-06-21 |
| 15 | 2011-08-10 | 2012-04-02 | 2013-05-29 |
| 16 | 2013-07-08 | 2014-04-07 | 2015-01-28 |
| 17 | 2015-02-23 | 2015-11-03 | 2016-09-02 |
| 18 | 2016-10-03 | 2017-07-17 | 2018-05-21 |
| 19 | 2018-06-19 | 2019-06-20 | 2020-11-04 |

Start, mid (median), and end dates for each RCCS survey round. Viral load and sequencing data used in this study were collected during R15-19. R19 sequence data was generated for participant visits through 2019-05-17.

**Supplementary Figure 1: Rakai Community Cohort Study interview dates**

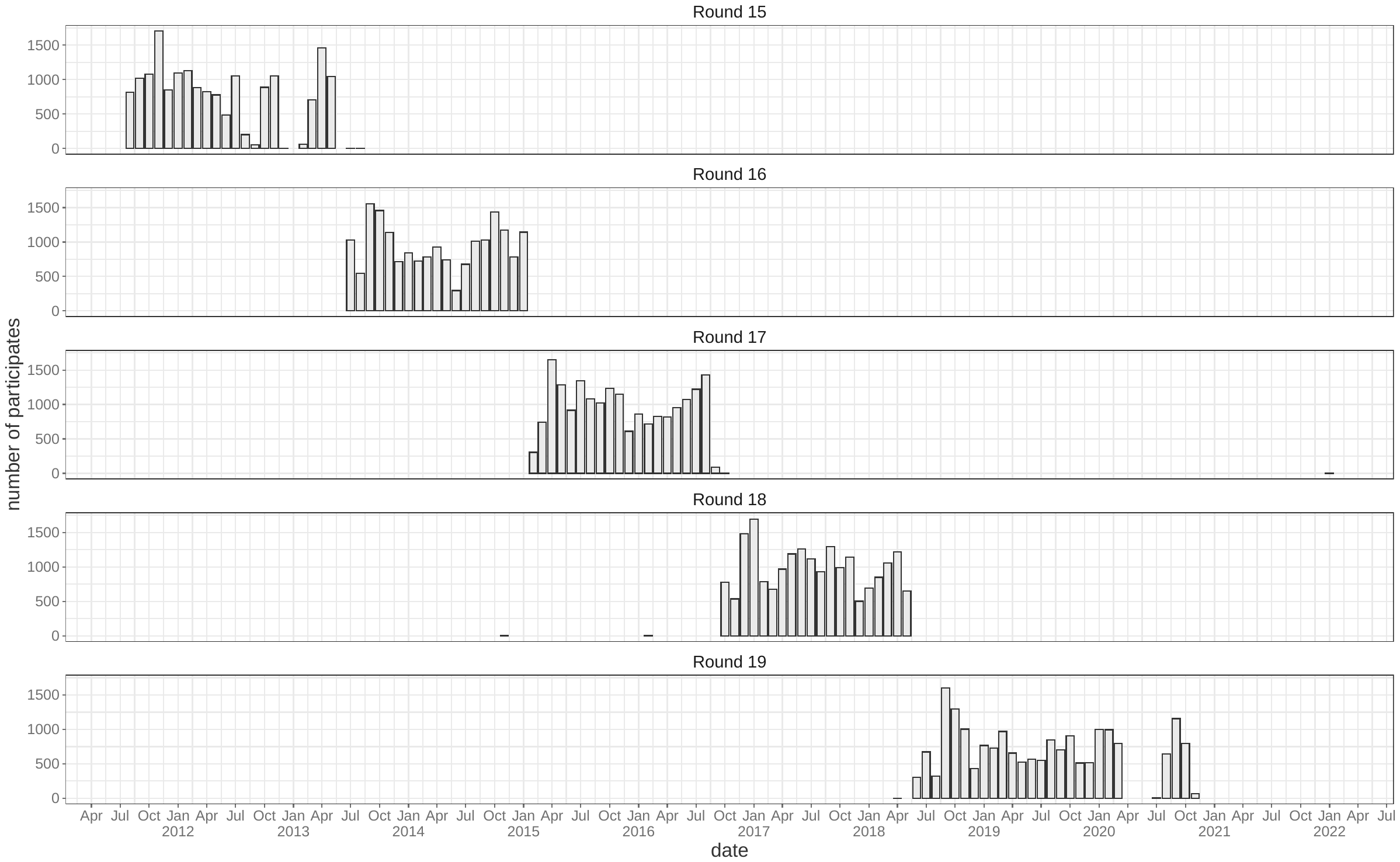

Number of interviews conducted each month, stratified by Rakai Community Cohort Study round.

**Supplementary Table 2: Recommended ART regimen among RCCS participants by year**

| **Year** | **First line regimen** | **Second line regimen** |
| --- | --- | --- |
| 2012 | **Preferred** | **Preferred** |
|  | TDF/3TC + NVP | AZT + 3TC + ATV/r |
|  | TDF/3TC + EFV | AZT + 3TC + LPV/r |
|  | **Alternative** | **Alternative** |
|  | AZT/3TC + NVP | TDF + 3TC + ATV/r |
|  | AZT/3TC + EFV | TDF + 3TC + LPV/r |
| 2016 | **Preferred** | **Preferred** |
|  | TDF+3TC+EFV | 2 NRTIs and ritonavir-boosted atazanavir (ATV/r) |
|  |  | e.g. |
|  | **Alternative** **(only if EFV is contraindicated)** | After failing on TDF + 3TC or ABC+3TC based regimen: AZT+3TC / ATV/r |
|  | TDF+3TC+DTG |  |
|  |  | Αφτερ φαιλινγ ον ΑΖΤ+3ΤΧ βασεδ ρεγιμεν: ΤΔΦ + 3ΤΧ/ ΑΤς/ρ |
| 2018 | **Preferred** | **Preferred** |
|  | TDF+3TC+DTG | AZT+3TC+ATV/r |
|  | **Alternative** | **Alternative** |
|  | TDF+3TC+EFV, ABC+3TC+DTG | AZT+3TC+LPV/r |
|  |  | AZT+3TC+DTG |
|  | **Adult women and adolescent girls of childbearing potential who are pregnant, intend to get pregnant or not on effective contraception** |  |
|  | TDF+3TC+EFV |  |
|  | **Alternative** |  |
|  | TDF+3TC+ATVr or ABC+3TC+EFV |  |
| 2022 | **Preferred** | **Preferred** |
|  | TDF + 3TC + DTG | AZT+3TC+ATV/r |
|  |  | TDF+3TC+ATV/r |
|  | **Alternatives** |  |
|  | TDF + 3TC + EFV400 | **Alternative** |
|  | ABC + 3TC +DTG | **AZT+3TC+LPV/r** |
|  | ABC +3TC +EFV400 | **TDF+3TC+LPV/r** |
|  | TDF +3TC + ATV/r or ABC |  |
|  | 3TC + ATV/r |  |

**Supplementary Methods**

**HIV deep sequencing**

For samples taken at participant visits during the 2012 and 2014 survey rounds RNA extraction was performed at University College London Hospital (London, United Kingdom) using the QIAsymphony SP workstation with the QIAsymphony DSP Virus/Pathogen Kit (Qiagen, Hilden, Germany). cDNA from four overlapping amplicons covering the HIV-1 genome was generated using in a one-step reverse-transcription protocol using universal primers^1^. Following gel electrophoresis confirmation of amplification samples were sequenced at the Wellcome Trust Sanger Institute (Hinxton, United Kingdom) on Illumina MiSeq and HiSeq platforms.

For the remaining survey rounds, RNA extraction was performed as above at the Oxford Genomics Centre (Oxford, United Kingdom) followed by library preparation with the SMARTer Stranded Total RNA-Seq kit v2 – Pico Input Mammalian (Clontech, TakaRa Bio). Streptavidin-conjugated beads were used to isolate fragments shorter than 400 nucleotides and PCR amplification and purification was performed with the Agencourt AMPure XMP (Beckman Coulter) following the veSEQ-HIV protocol.^2^The Oxford Genomics Centre generated 350 to 600 base pair reads using the Illumina NovaSeq 6000 instrument. A subset of poorly-sequenced pre-treatment samples from the 2012 and 2014 survey rounds were also sequenced using this protocol (appendix p 23).

Sequencing reads were filtered with trimmomatic^3^ to remove any reads with less than 50 bases, remove leading and trailing bases if their quality score was below 20, and cut reads when the average quality score in a 4 base sliding window was less than 20.

**Data attributes**

For each participant-visit $i$, we assign the following attributes:

- $S_{i}$: Categorical variable indicating which survey round participant-visit $i$ was contributed towards
- $H_{i}$: Binary variable indicating whether participant-visit $i$ was contributed by a person living with HIV (PLHIV) as determined by a validated rapid-test algorithm (*Methods*)*.*
- $Q_{i}:$ Binary variable indicating whether a viral load measurement is available for participant-visit $i$. Among samples with $Q_{i}=1$, viral loads were measured using the Abbott real-time m2000 assay (Abbott Laboratories).
- $V_{i}$: binary variable indicating whether participant-visit $i$ is both seropositive and viremic. Among participant-visits with an available viral load measurement, viremia was defined as $\geq1000$ copies/mL. Viral load measurements were routinely conducted for PLHIV in the 2014 through 2019 survey rounds and missing data is minimal (see: *Results*). In the 2012 survey round, viral load measurements were only available for residents of Fishing communities. We impute missing 2012 survey viral loads among pre-treatment PLHIV (*see below).* Treatment-experienced participant-visits contributed to the 2012 survey round are not considered. Assumed to be 0 for all $i$ with $H_{i}$ = 0 or $Q_{i}=0$.
- $T_{i}$: binary variable indicating whether participant-visit $i$ is treatment experienced. Treatment-experienced is defined as having reported being on treatment during the survey round that participant visit $i$ was contributed to or any earlier survey rounds.
- $R_{i}^{c}$: binary variable indicating whether participant-visit $i$ has available resistance prediction to drug class $c$ (NNRTI, NRTI, PI, joint NNRTI/NRTI/PI, and joint INSTI/NNRTI/NRTI/PI). Additionally, $R_{i}^{any}$indicates available resistance prediction to at least one drug (i.e. exclude sequenced samples with insufficient coverage to predict resistance for all included drugs). Assumed to be 0 for all $i$ with $V_{i}$ = 0 or $H_{i}$ = 0.
- $D_{i}^{c}$: binary variable indicating whether participant-visit $i$ has predicted viremic resistance to drug class $c$. Assumed to be 0 for all $i$ with $V_{i}$ = 0 or $H_{i}$ = 0.
- $M_{m,i}$: binary vector for each participant-visit indicating presence/absence of each resistance conferring mutation $m$.

We note that individual participants can contribute up to one participant-visit per survey round, conditional on being a resident of a participating community during that survey.

**Imputation of missing round 2012 pre-treatment viral loads**

Viral load measurements were conducted for only a subset of participants in the 2012 survey round. We here focus on participant-visits contributed by viremic PLHIV. Consequently, to avoid discarding a large proportion of 2012 participant-visits we impute $V_{i}$ for all pre-treatment $i$ contributed to the 2012 survey round with $Q_{i}=0$. As we do not attempt to estimate the prevalence of treatment-experienced viremic resistance in the 2012 survey round we do not do any imputation with for those $i$ with $Q_{i}=0, T_{i}=1$.

Specifically, because sequencing success depends on sample viral load^2^, we impute missing $V_{i}$ based on observed $V_{i}$ among 2012 survey participants with viral load measurements conditional on availability of resistance prediction to at least one drug ($R_{i}^{any}$).

We first calculate:

$$P\left( V=1 | R^{any}=1,Q=1, S=2012 \right)= \frac{\sum_{i\in2012} {{R_{i}^{any}V}_{i}Q}_{i}}{\sum_{i\in2012} V_{i}Q_{i}}$$

$$P\left( V=1 | R^{any}=0,Q=1, S=2012 \right)= \frac{\sum_{i\in2012} {{{(\sim R}_{i}^{any})V}_{i}Q}_{i}}{\sum_{i\in2012} V_{i}Q_{i}}$$

Where $\sim R_{i}^{any}$ indicates “not $R_{i}^{any}$”.

And then impute missing viral loads as:

$$\left( V_{i} | {R_{i}^{any}=1, Q}_{i}=0, S=2012 \right)\sim\text{Bernoulli}(P\left( V=1 | R^{any}=1,Q=1, S=2012 \right))$$

$$\left( V_{i} | {R_{i}^{any}=0, Q}_{i}=0, S=2012 \right)\sim\text{Bernoulli}(P\left( V=1 | R^{any}=0,Q=1, S=2012 \right))$$

**Sampling probabilities**

For all outcome measures that relied on sequence-based resistance predictions we used inverse probability weighting to account for the fact that sequence data was not available for all participant-visits contributed by viremic PLHIV and that resistance prediction was unsuccessful for some sequenced participant-visits. First, among all viremic participant-visits ($V_{i}=1$) we estimate the probability that resistance prediction is available for each drug class ($c$, NNRTI, NRTI, PI individually and composite NNRTI&NRTI&PI (NNP for short) and INSTI&NNRTI&NRTI&PI (INNP for short)) based on epidemiological covariates ($X_{i}$). This analysis is stratified by survey round, $s$.

We define $\pi_{i}^{s,c}$ as the probability that resistance prediction to class $c$ is available for participant-visit $i$ conditional on $i$ being contributed to survey round $s$ and $V_{i}=1$. In mathematical notation: $\pi_{i}^{s,c}= P(R^{c}=1|{X_{i}, V}_{i}=1, {H_{i}=1, S}_{i}=s)$. We estimate $\pi_{i}^{s,c}$ and all $\beta$ using robust Poisson regression for each survey round ($s$):

$$\text{log}\left( \frac{\pi_{i}^{s,c}}{1-\pi_{i}^{s,c}} \right)= \beta_{0}^{s,c}+ \beta_{1}^{s,c}X_{i}^{\text{treatment-experienced}}+ \beta_{2}^{s,c}X_{i}^{\text{missing viral load}}+ \beta_{3}^{s,c}X_{i}^{\text{viral load}}+\beta_{4}^{rsc}X_{i}^{\text{trading}}+\beta_{5}^{s,c}X_{i}^{\text{fishing}}+\beta_{6}^{s,c}X_{i}^{\text{25-34 yrs. }}+ \beta_{7}^{s,c}X_{i}^{\text{35-49 yrs. }}+ \beta_{8}^{s,c}X_{i}^{\text{male }}+ \varepsilon_{i}$$

Where $X_{i}^{j}$are binary indicator variables with the exception of $X_{i}^{viral load}$, which is the log_10_ viral load (copies/mL) for participant-visit $i$ and is 0 for all $i$with missing viral loads ($X_{i}^{\text{missing viral load}}=1$). Sampling probability model estimates are provided in table S5 (appendix pp 14-16).

Sampling weights are calculated based on sampling probabilities for each outcome of interest depending on the population under study, as described below.

**Outcome measures**

A summary of primary outcome measures and corresponding figure references is provided in table S2 (appendix pp 11-12). Outcomes below are listed in order of their appearance in the *Results* section.

We estimate each quantity using a robust Poisson regression with survey round as a predictor variable using general estimating equations (GEE) to account for repeated measures within participants. The correlation structure that minimizes Quasi Information Criterion (QIC) for each outcome was chosen. Estimates stratified by epidemiological covariates used the correlation structure from the corresponding non-stratified analysis.

*Population prevalence of all PLHIV, viremic PLHIV, viremic pre-treatment PLHIV, and viremic treatment-experienced PLHIV among study participants*

Here, the population under study is all contributed participant-visits. The quantities of interest are the prevalence of all PLHIV, all viremic PLHIV, all viremic pre-treatment PLHIV, and viremic treatment-experienced PLHIV among study participants in each survey round $s$. In mathematical notation, we estimate the following quantities ($q$) for all $s$:

- Prevalence of all PLHIV: $P(H=1|S=s)$
- Prevalence of viremic PLHIV: $P(V=1, H=1|S=s)$
- Prevalence of viremic pre-treatment PLHIV: $P(T=0, V=1,H=1|S=s)$
- Prevalence of viremic treatment-experienced PLHIV: $P(T=1,V=1,H=1|S=s)$

Due to missing viral loads, particularly among treatment-experienced PLHIV, we did not attempt to estimate the prevalence of viremic PLHIV or the prevalence of viremic treatment-experienced PLHIV in the 2012 survey round.

As these outcomes did not depend on the availability to sequence-based resistance prediction, inverse probability weighting was not employed.

*Population prevalence of viremic NNRTI, NRTI, and PI resistance among study participants*

Here, the population under study is all contributed participant-visits. The quantities of interest are the prevalence of NNRTI, NRTI, and PI resistance among study participants in each survey round $s$. We estimate the following quantities for all $s$:

- Prevalence of viremic NNRTI resistance: $P(D^{NNRTI}=1, V=1, H=1|S=s)$
- Prevalence of viremic NRTI resistance: $P(D^{NRTI}=1, V=1, H=1|S=s)$
- Prevalence of viremic PI resistance: $P(D^{PI}=1, V=1, H=1|S=s)$

Due to missing sequence-data among treatment-experienced PLHIV in the 2012, 2014, and 2019 survey rounds this analysis was restricted to the 2015 and 2017 rounds. Bivariate models were used to evaluate whether age category, sex, community type, or treatment status was associated with each outcome. Significant covariates were further analyzed in stratified analysis.

Viremic participant-visits lacking resistance prediction were dropped and viremic participant-visits with resistance prediction were upweighted accordingly using inverse probability weighting. Specifically, participant-visits in round $s$ were assigned weight for resistance class $c$ ($w_{i}^{s,c}$) according to:

$$w_{i}^{s,c}= \left\{ \begin{aligned} 1, &H_{i}=0 \\ 1, &V_{i}=0, H_{i}=1 \\ 0, &R_{i}^{c}=0, V_{i}=1, H_{i}=1 \\ \left( \sum_{j\in s} V_{j}H_{j} \right)\frac{\frac{1}{\pi_{i}^{s,c}}}{\sum_{j\in s} \left( R_{j}^{c}\frac{1}{\pi_{j}^{s,c}} \right)}, &R_{i}^{c}=1, V_{i}=1, H_{i}=1 \end{aligned} \right.$$

Where $i\in s$ indicates all participant-visits contributed to round $s$. Normalizing by the sum of all weights contributed to round $s$ and multiplying by the total number of viremic PLHIV in round $s$ ensures that the sum of the weights is equal to the total number of viremic PLHIV in round $s$ ($\sum_{i\in s} w_{i}^{,s,c}=\sum_{i\in s} V_{i}H_{i}$).

*Population prevalence of pre-treatment viremic resistance among study participants*

Here, the population under study is all contributed participant-visits. The quantity of interest is the prevalence of NNRTI, NRTI, and PI pre-treatment viremic resistance among study participants in each survey round $s$. We estimate the following quantities for all $s$:

- Prevalence of pre-treatment viremic NNRTI resistance: $P(D^{NNRTI}=1, T=0, V=1, H=1|S=s)$
- Prevalence of pre-treatment viremic NRTI resistance: $P(D^{NRTI}=1, T=0, V=1, H=1|S=s)$
- Prevalence of pre-treatment viremic PI resistance: $P(D^{PI}=1, T=0, V=1, H=1|S=s)$

Pre-treatment viremic participant-visits lacking resistance prediction were dropped and pre-treatment viremic participant-visits with resistance prediction were upweighted accordingly using inverse probability weighting. Treatment-experienced viremic participant-visits were assigned a weight of 1. Specifically, participant-visits in round $s$ were assigned weight for resistance class $c$ ($w_{i}^{s,c}$) according to:

$$w_{i}^{s,c}= \left\{ \begin{aligned} 1, &H_{i}=0 \\ 1, &V_{i}=0, H_{i}=1 \\ 1, &{T_{i}=1, V}_{i}=1, H_{i}=1 \\ 0, &R_{i}^{c}=0, {T_{i}=0, V}_{i}=1, H_{i}=1 \\ \left( \sum_{j\in s} \left( {(\sim T_{j})V}_{j}H_{j} \right) \right)\frac{\frac{1}{\pi_{i}^{s,c}}}{\sum_{j\in s} \left( {(\sim T_{j})R}_{j}^{c}\frac{1}{\pi_{j}^{s,c}} \right)}, &R_{i}^{c}=1, {T_{i}=0, V}_{i}=1, H_{i}=1 \end{aligned} \right.$$

Where $\sim T_{j}$ is 1 when $T_{j}=0$.

*Population prevalence of treatment-experienced viremic resistance among study participants*

Here, the population under study is all contributed participant-visits. The quantity of interest is the prevalence of NNRTI, NRTI, and PI treatment-experienced viremic resistance among study participants in each survey round $s$. We estimate the following quantities for all $s$:

- Prevalence of treatment-experienced viremic NNRTI resistance:
   $P(D^{NNRTI}=1, T=1, V=1, H=1|S=s)$
- Prevalence of treatment-experienced viremic NRTI resistance:

$P(D^{NRTI}=1, T=1, V=1, H=1|S=s)$

- Prevalence of treatment-experienced viremic PI resistance:

$P(D^{PI}=1, T=1, V=1, H=1|S=s)$

Due to missing sequence-data among treatment-experienced PLHIV in the 2012, 2014, and 2019 survey rounds this analysis was restricted to the 2015 and 2017 rounds.

Treatment-experienced viremic participant-visits lacking resistance prediction were dropped and treatment-experienced viremic participant-visits with resistance prediction were upweighted accordingly using inverse probability weighting. Treatment-experienced viremic participant-visits were assigned a weight of 1. Specifically, participant-visits in round $s$ were assigned weight for resistance class $c$ ($w_{i}^{s,c}$) according to:

$$w_{i}^{s,c}= \left\{ \begin{aligned} 1, &H_{i}=0 \\ 1, &V_{i}=0, H_{i}=1 \\ 1, &{T_{i}=0, V}_{i}=1, H_{i}=1 \\ 0, &R_{i}^{c}=0, {T_{i}=1, V}_{i}=1, H_{i}=1 \\ \left( \sum_{j\in s} T_{j}V_{j}H_{j} \right)\frac{\frac{1}{\pi_{i}^{s,c}}}{\sum_{j\in s} \left( {T_{j}R}_{j}^{c}\frac{1}{\pi_{j}^{s,c}} \right)}, &R_{i}^{c}=1, {T_{i}=1, V}_{i}=1, H_{i}=1 \end{aligned} \right.$$

*Population prevalence of viremic multi-class resistance among study participants*

Here, the population under study is all contributed participant-visits. The quantity of interest is the prevalence of combinations of multi-class NNRTI, NRTI, and PI resistance among study participants in each survey round $s$. We estimate the following quantities for all $s$:

- Prevalence of viremic NNRTI mono-resistance:

$P(D^{NNRTI}=1, D^{NRTI}=0, D^{PI}=0,V=1,H=1|S=s)$

- Prevalence of viremic NRTI mono-resistance:

$P(D^{NNRTI}=0, D^{NRTI}=1, D^{PI}=0,V=1,H=1|S=s)$

- Prevalence of viremic PI mono-resistance:

$P(D^{NNRTI}=0, D^{NRTI}=0, D^{PI}=1,V=1,H=1|S=s)$

- Prevalence of viremic NNRTI/NRTI dual-resistance:

$P(D^{NNRTI}=1, D^{NRTI}=1, D^{PI}=0,V=1,H=1|S=s)$

- Prevalence of viremic NNRTI/PI dual-resistance:

$P(D^{NNRTI}=1, D^{NRTI}=0, D^{PI}=1,V=1,H=1|S=s)$

- Prevalence of viremic NRTI/PI dual-resistance:

$P(D^{NNRTI}=0, D^{NRTI}=1, D^{PI}=1,V=1,H=1|S=s)$

- Prevalence of viremic triple-resistance:

$P(D^{NNRTI}=1, D^{NRTI}=1, D^{PI}=1,V=1,H=1|S=s)$

Due to missing sequence-data among treatment-experienced PLHIV in the 2012, 2014, and 2019 survey rounds this analysis was restricted to the 2015 and 2017 rounds.

Viremic participant-visits lacking resistance prediction were dropped and viremic participant-visits with resistance prediction were upweighted accordingly using inverse probability weighting. For all quantities, we use the sampling probabilities calculated based on the availability of resistance prediction to NNRTIs, NRTI and PIs, $\pi_{i}^{s,NNP}$. Specifically, participant-visits in round $s$ were assigned weights ($w_{i}^{s,NNP}$) according to:

$$w_{i}^{s,NNP}= \left\{ \begin{aligned} 1, &H_{i}=0 \\ 1, &V_{i}=0, H_{i}=1 \\ 0, &R_{i}^{NNP}=0, V_{i}=1, H_{i}=1 \\ \left( \sum_{j\in s} V_{i}H_{i} \right)\frac{\frac{1}{\pi_{i}^{s,NNP}}}{\sum_{j\in s} \left( R_{j}^{NNP}\frac{1}{\pi_{j}^{s,NNP}} \right)}, &R_{i}^{NNP}=1, V_{i}=1, H_{i}=1 \end{aligned} \right.$$

*Prevalence of resistance among pre-treatment viremic participants*

Here, the population under study is pre-treatment viremic participant-visits (all $i$ with ${T_{i}=0, V}_{i}=1,H_{i}=1$). The quantity of interest is the prevalence of NNRTI, NRTI, and PI resistance among pre-treatment viremic participants in each survey round $s$. We estimate the following quantities for all $s$:

- Prevalence of NNRTI resistance among pre-treatment viremic participant-visits:
   $P\left( D^{NNRTI}=1 \right|T=0, V=1, H=1, S=s)$
- Prevalence of NRTI resistance among pre-treatment viremic participant-visits:

$P(D^{NRTI}=1 |T=0,V=1,H=1,S=s)$

- Prevalence of PI resistance among pre-treatment viremic participant-visits:

$P\left( D^{PI}=1 \right|T=0, V=1,H=1,S=s)$

We additionally estimate each quantity stratified by sex, age category, and community type. Bivariate models were used to evaluate whether age category, sex, or community type was associated with each outcome. Significant covariates were further analyzed in stratified analysis.

Pre-treatment viremic participant-visits lacking resistance prediction were dropped and pre-treatment viremic participant-visits with resistance prediction were upweighted accordingly using inverse probability weighting. Specifically, participant-visits in round $s$ were assigned weight for resistance class $c$ ($w_{i}^{s,c}$) according to:

$$w_{i}^{s,c}= \left\{ \begin{aligned} 0, &H_{i}=0 \\ 0, &V_{i}=0, H_{i}=1 \\ 0, &{T_{i}=1, V}_{i}=1, H_{i}=1 \\ 0, &R_{i}^{c}=0, {T_{i}=0, V}_{i}=1, H_{i}=1 \\ \left( \sum_{i\in s} \left( \left( \sim T_{i} \right)V_{i}H_{i} \right) \right)\frac{\frac{1}{\pi_{i}^{s,c}}}{\sum_{i\in s} \left( {{(\sim T}_{i})R}_{i}^{c}\frac{1}{\pi_{i}^{s,c}} \right)}, &R_{i}^{c}=1, {T_{i}=0, V}_{i}=1, H_{i}=1 \end{aligned} \right.$$

*Prevalence of resistance conferring mutations among pre-treatment viremic participants*

Here, the population under study is pre-treatment viremic participant-visits (${{\mathrm{all}i\mathrm{with}T}_{i}=0, V}_{i}=1,H_{i}=1$). The quantity of interest is the prevalence of each resistance mutations $m$ among pre-treatment viremic participants in each survey round $s$. For each mutation, we estimate the following quantity for all $s$:

- Prevalence of $m$ among pre-treatment viremic participant-visits:
   $P(M_{m}=1|T=0,V=1,H=1,S=s)$

Pre-treatment viremic participant-visits lacking resistance prediction were dropped and pre-treatment viremic participant-visits with resistance prediction were upweighted accordingly using inverse probability weighting. For all mutations, we use the sampling probabilities calculated based on the availability of resistance prediction to INSTIs, NNRTIs, NRTIs, and PIs, $\pi_{i}^{s,INNP}$. Specifically, participant-visits in round $s$ were assigned weights ($w_{i}^{s,INNP}$) according to:

$$w_{i}^{s,INNP}= \left\{ \begin{aligned} 0, &H_{i}=0 \\ 0, &V_{i}=0, H_{i}=1 \\ 0, &{T_{i}=1, V}_{i}=1, H_{i}=1 \\ 0, &R_{i}^{INNP}=0, {T_{i}=0, V}_{i}=1, H_{i}=1 \\ \left( \sum_{j\in s} \left( \left( \sim T_{j} \right)V_{j}H_{j} \right) \right)\frac{\frac{1}{\pi_{i}^{s,INNP}}}{\sum_{j\in s} \left( {{(\sim T}_{j})R}_{j}^{INNP}\frac{1}{\pi_{j}^{s,INNP}} \right)}, &R_{i}^{INNP}=1, {T_{i}=0, V}_{i}=1, H_{i}=1 \end{aligned} \right.$$

*Prevalence of resistance among treatment-experienced viremic participants*

Here, the population under study is treatment-experienced viremic participant-visits (all $i$with ${T_{i}=1, V}_{i}=1,H_{i}=1$). The quantity of interest is the prevalence of NNRTI, NRTI, and PI resistance among treatment-experienced viremic participants in each survey round $s$. We estimate the following quantities for all $s$:

- Prevalence of NNRTI resistance among treatment-experienced viremic participant-visits:
   $P\left( D^{NNRTI}=1 \right|T=1, V=1, H=1, S=s)$
- Prevalence of NRTI resistance among treatment-experienced viremic participant-visits:

$P(D^{NRTI}=1 |T=1,V=1,H=1,S=s)$

- Prevalence of PI resistance among treatment-experienced viremic participant-visits:

$P\left( D^{PI}=1 \right|T=1, V=1,H=1,S=s)$

Due to missing sequence-data among treatment-experienced PLHIV in the 2012, 2014, and 2019 survey rounds this analysis was restricted to the 2015 and 2017 rounds. Bivariate models were used to evaluate whether age category, sex, or community type was associated with each outcome. Significant covariates were further analyzed in stratified analysis.

Treatment-experienced viremic participant-visits lacking resistance prediction were dropped and treatment-experienced viremic participant-visits with resistance prediction were upweighted accordingly using inverse probability weighting. Specifically, participant-visits in round $s$ were assigned weight for resistance class $c$ ($w_{i}^{s,c}$) according to:

$$w_{i}^{s,c}= \left\{ \begin{aligned} 0, &H_{i}=0 \\ 0, &V_{i}=0, H_{i}=1 \\ 0, &{T_{i}=0, V}_{i}=1, H_{i}=1 \\ 0, &R_{i}^{c}=0, {T_{i}=1, V}_{i}=1, H_{i}=1 \\ \left( \sum_{j\in s} \left( T_{j}V_{j}H_{j} \right) \right)\frac{\frac{1}{\pi_{i}^{s,c}}}{\sum_{j\in s} \left( {T_{j}R}_{j}^{c}\frac{1}{\pi_{j}^{s,c}} \right)}, &R_{i}^{c}=1, {T_{i}=1, V}_{i}=1, H_{i}=1 \end{aligned} \right.$$

*Prevalence of resistance conferring mutationsamong treatment-experienced viremic participants*

Here, the population under study is treatment-experienced viremic participant-visits (${{\mathrm{all}i\mathrm{with}T}_{i}=1, V}_{i}=1,H_{i}=1$). The quantity of interest is the prevalence of each resistance mutation $m$ among pre-treatment viremic participants in each survey round $s$. For each mutation, we estimate the following quantity for all $s$:

- Prevalence of $m$ among treatment-experienced viremic participant-visits:
   $P(M_{m}=1|T=1,V=1,H=1,S=s)$

Due to missing sequence-data among treatment-experienced PLHIV in the 2012, 2014, and 2019 survey rounds this analysis was restricted to the 2015 and 2017 rounds.

Treatment-experienced viremic participant-visits lacking resistance prediction were dropped and treatment-experienced viremic participant-visits with resistance prediction were upweighted accordingly using inverse probability weighting. For all mutations, we use the sampling probabilities calculated based on the availability of resistance prediction to INSTIs, NNRTIs, NRTIs, and PIs, $\pi_{i}^{s,INNP}$. Specifically, participant-visits in round $s$ were assigned weights ($w_{i}^{s,INNP}$) according to:

$$w_{i}^{s,INNP}= \left\{ \begin{aligned} 0, &H_{i}=0 \\ 0, &V_{i}=0, H_{i}=1 \\ 0, &{T_{i}=0, V}_{i}=1, H_{i}=1 \\ 0, &R_{i}^{INNP}=0, {T_{i}=1, V}_{i}=1, H_{i}=1 \\ \left( \sum_{j\in s} \left( T_{j}V_{j}H_{j} \right) \right)\frac{\frac{1}{\pi_{i}^{s,INNP}}}{\sum_{j\in s} \left( {T_{j}R}_{j}^{INNP}\frac{1}{\pi_{j}^{s,INNP}} \right)}, &R_{i}^{INNP}=1, {T_{i}=1, V}_{i}=1, H_{i}=1 \end{aligned} \right.$$

**Supplementary Table 3: Summary of outcome measures**

| **Measure** | **Model outcome (Numerator)** | **Population (Denominator)** | **Pseudo-population (IPW population)** | **Model predictors** | **Sub-group analyses** | **Corresponding figure** |
| --- | --- | --- | --- | --- | --- | --- |
| population prevalence of PLHIV in RCCS | HIV seropositive | all RCCS participants | all RCCS participants | survey round (2012 [REF], 2014, 2015, 2017, 2019) | .. | Fig 1A |
| population prevalence of viremic PLHIV in RCCS | viremic and HIV seropositive | all RCCS participants | all RCCS participants | survey round (2014 [REF], 2015, 2017, 2019) | .. | Fig 1A |
| population prevalence of viremic pre-treatment PLHIV in RCCS | pre-treatment, viremic, and HIV seropositive | all RCCS participants | all RCCS participants | survey round (2012 [REF], 2014, 2015, 2017, 2019) | .. | Fig 1A |
| population prevalence of viremic treatment-experienced PLHIV in RCCS | treatment-experienced, viremic, and HIV seropositive | all RCCS participants | all RCCS participants | survey round (2014 [REF], 2015, 2017, 2019) | .. | Fig 1A |
| population prevalence of viremic NNRTI, NRTI, and PI resistance | viremic NNRTI, NRTI, and PI resistance | viremic PLHIV with resistance prediction to all NNRTIs, NRTIs, or PIs; non-viremic PLHIV; participants not living with HIV | all RCCS participants | survey round (2015 [REF] and 2017) | Age category ((14,24] [REF], (24,34], and (34,49]) Community type (agrarian [REF], fishing, and trading) Sex (female [REF], male) | Fig 1B |
| population prevalence of viremic pre-treatment NNRTI, NRTI, and PI resistance | viremic pre-treatment NNRTI, NRTI, and PI resistance | viremic pre-treatment PLHIV with resistance prediction to all NNRTIs, NRTIs, or PIs; non-viremic pre-treatment PLHIV; treatment-experienced PLHIV; participants not living with HIV | all RCCS participants | survey round (2012 [REF], 2014, 2015, 2017, 2019) | .. | Fig 1C |
| population prevalence of viremic treatment-experienced NNRTI, NRTI, and PI resistance | viremic treatment-experienced NNRTI, NRTI, and PI resistance | viremic treatment-experienced PLHIV with resistance prediction to all all NNRTIs, NRTIs, or PIs; non-viremic treatment-experienced PLHIV; pre-treatment PLHIV; participants not living with HIV | all RCCS participants | survey round (2015 [REF] and 2017) | .. | Fig 1D |
| population prevalence of multi-class resistance | viremic NNRTI, NRTI, PI, NNRTI/NRTI, NNRTI/PI, NRTI/PI, and NNRTI/NRTI/PI resistance | viremic PLHIV with resistance prediction to all NNRTIs, NRTIs, and PIs; non-viremic PLHIV; participants not living with HIV | all RCCS participants | survey round (2015 [REF] and 2017) | .. | Fig 2A |
| prevalence of NNRTI, NRTI, and PI resistance among viremic pre-treatment PLHIV | viremic NNRTI, NRTI, and PI resistance | viremic pre-treatment PLHIV with resistance prediction to all NNRTIs, NRTIs, or PIs. | viremic pre-treatment PLHIV | survey round (2012 [REF], 2014, 2015, 2017, 2019) | Age category ((14,24] [REF], (24,34], and (34,49]) Community type (agrarian [REF], fishing, and trading) Sequencing technology (amplicon v. veSeq-HIV) Sex (female [REF], male) | Fig 3A |
| prevalence of resistance mutations among viremic pre-treatment PLHIV | mutation presence | viremic pre-treatment PLHIV with resistance prediction to all INSTIs, NNRTIs, NRTIs, and PIs. | viremic pre-treatment PLHIV | survey round (2012 [REF], 2014, 2015, 2017, 2019) | .. | Fig 3B |
| prevalence of NNRTI, NRTI, and PI resistance among viremic treatment-experienced PLHIV | viremic NNRTI, NRTI, and PI resistance | viremic treatment-experienced PLHIV with resistance prediction to all NNRTI, NRTI, or PIs. | viremic treatment-experienced PLHIV | survey round (2015 [REF] and 2017) | Age category ((14,24] [REF], (24,34], and (34,49]) Community type (agrarian [REF], fishing, and trading) Sex (female [REF], male) | Fig 4A |
| prevalence of resistance mutations among viremic treatment-experienced PLHIV | mutation presence | viremic treatment-experienced PLHIV with resistance prediction to all INSTIs, NNRTIs, NRTIs, and PIs. | viremic treatment-experienced PLHIV | survey round (2015 [REF] and 2017) | .. | Fig 4B |

Measure refers to how a given analysis is referred to in text, figures, and tables. The model outcome is the participant-visit level characteristic which is being modeled in each analysis. The population is the data among which a given outcome is modeled. Pseudo-population is the population which is being approximated using IPW. Model predictors are the variables included in each model. Interaction included with model predictors in bivariate analyses. PLHIV = people living with HIV. NNRTI = non-nucleoside reverse transcriptase inhibitors. NRTI = nucleoside reverse transcriptase inhibitors. PI = protease inhibitors. INSTI = integrase strand transfer inhibitors. IPW = inverse probability weighting.

**Supplementary Table 4: Drugs to which resistance was predicted**

| **Generic name** | **Abbreviation** | **Class** |
| --- | --- | --- |
| bictegravir | BIC | INSTI |
| cabotegravir | CAB | INSTI |
| dolutegravir | DTG | INSTI |
| elvitegravir | EVG | INSTI |
| raltegravir | RAL | INSTI |
| doravirine | DOR | NNRTI |
| efavirenz | EFV | NNRTI |
| Etravirine | ETR | NNRTI |
| nevirapine | NVP | NNRTI |
| rilpivirine | RPV | NNRTI |
| abacavir | ABC | NRTI |
| didanosine | DDI | NRTI |
| emtricitabine | FTC | NRTI |
| lamivudine | 3TC | NRTI |
| stavudine | D4T | NRTI |
| tenofovir | TDF | NRTI |
| zidovudine | AZT | NRTI |
| atazanavir | ATV | PI |
| darunavir | DRV | PI |
| fosamprenavir | FPV | PI |
| indinavir | IDV | PI |
| lopinavir | LPV | PI |
| nelfinavir | NFV | PI |
| saquinavir | SQV | PI |
| tipranavir | TPV | PI |

INSTI = integrase strand transfer inhibitor. NNRTI = non-nucleoside reverse transcriptase inhibitor. NRTI = nucleoside reverse transcriptase inhibitor. PI = protease inhibitor.

**Supplementary Table 5: Sampling probability model for the probability that genotyping data is available among all viremic PLHIV**

|  |  | **NNRTI** | |  | **NRTI** | |  | **PI** | |  | **NNRTI/NRTI/PI** | |  | **INSTI/NRTI/PI** | |
| --- | --- | --- | --- | --- | --- | --- | --- | --- | --- | --- | --- | --- | --- | --- | --- |
| **Survey round** | **Variable** | **Coeff. (95% CI)** | ***p*-value** |  | **Coeff. (95% CI)** | ***p*-value** |  | **Coeff. (95% CI)** | ***p*-value** |  | **Coeff. (95% CI)** | ***p*-value** |  | **Coeff. (95% CI)** | ***p*-value** |
| 2012 | (Intercept) | 0.04 (0.02, 0.06) | <0.0001 |  | 0.03 (0.02, 0.05) | <0.0001 |  | 0.13 (0.09, 0.17) | <0.0001 |  | 0.03 (0.02, 0.04) | <0.0001 |  | 0 (0, 0.01) | <0.0001 |
|  | Age category |  |  |  |  |  |  |  |  |  |  |  |  |  |  |
|  | (14,24] | ref | ref |  | ref | ref |  | ref | ref |  | ref | ref |  | ref | ref |
|  | (24,34] | 1 (0.89, 1.12) | 1 |  | 1.01 (0.91, 1.13) | 0.83 |  | 1.02 (0.94, 1.1) | 0.65 |  | 0.99 (0.88, 1.12) | 0.89 |  | 0.94 (0.77, 1.15) | 0.55 |
|  | (34,49] | 0.98 (0.86, 1.11) | 0.74 |  | 1 (0.89, 1.14) | 0.94 |  | 1.03 (0.94, 1.12) | 0.54 |  | 0.97 (0.85, 1.11) | 0.68 |  | 0.92 (0.74, 1.15) | 0.47 |
|  | Community type |  |  |  |  |  |  |  |  |  |  |  |  |  |  |
|  | Agrarian | ref | ref |  | ref | ref |  | ref | ref |  | ref | ref |  | ref | ref |
|  | Fishing | 0.85 (0.72, 0.99) | 0.038 |  | 0.9 (0.77, 1.06) | 0.2 |  | 1.07 (0.95, 1.2) | 0.24 |  | 0.85 (0.72, 1) | 0.056 |  | 0.91 (0.66, 1.24) | 0.54 |
|  | Trading | 0.96 (0.85, 1.09) | 0.5 |  | 0.97 (0.86, 1.1) | 0.66 |  | 1 (0.92, 1.1) | 0.93 |  | 0.96 (0.84, 1.09) | 0.54 |  | 1.01 (0.81, 1.25) | 0.94 |
|  | Sex |  |  |  |  |  |  |  |  |  |  |  |  |  |  |
|  | F | ref | ref |  | ref | ref |  | ref | ref |  | ref | ref |  | ref | ref |
|  | M | 1.05 (0.96, 1.15) | 0.25 |  | 1.06 (0.98, 1.16) | 0.16 |  | 1.04 (0.98, 1.11) | 0.17 |  | 1.07 (0.97, 1.17) | 0.17 |  | 1.07 (0.92, 1.25) | 0.38 |
|  | Missing VL | 14.74 (9.73, 22.35) | <0.0001 |  | 16.35 (10.91, 24.49) | <0.0001 |  | 5.23 (3.93, 6.96) | <0.0001 |  | 18.49 (12.03, 28.43) | <0.0001 |  | 125.82 (60.52, 261.57) | <0.0001 |
|  | Log_10_ VL | 1.88 (1.72, 2.04) | <0.0001 |  | 1.91 (1.76, 2.07) | <0.0001 |  | 1.46 (1.38, 1.55) | <0.0001 |  | 1.97 (1.8, 2.15) | <0.0001 |  | 2.89 (2.51, 3.34) | <0.0001 |
| 2014 | (Intercept) | 0.02 (0.01, 0.04) | <0.0001 |  | 0.02 (0.01, 0.03) | <0.0001 |  | 0.05 (0.03, 0.08) | <0.0001 |  | 0.02 (0.01, 0.03) | <0.0001 |  | 0 (0, 0.01) | <0.0001 |
|  | Age category |  |  |  |  |  |  |  |  |  |  |  |  |  |  |
|  | (14,24] | ref | ref |  | ref | ref |  | ref | ref |  | ref | ref |  | ref | ref |
|  | (24,34] | 0.74 (0.62, 0.89) | 0.0013 |  | 0.76 (0.63, 0.92) | 0.0039 |  | 0.75 (0.64, 0.87) | 3e-04 |  | 0.75 (0.62, 0.91) | 0.0034 |  | 0.76 (0.58, 0.98) | 0.032 |
|  | (34,49] | 0.67 (0.55, 0.83) | 0.0002 |  | 0.73 (0.59, 0.9) | 0.0028 |  | 0.7 (0.58, 0.83) | <0.0001 |  | 0.7 (0.57, 0.87) | 0.001 |  | 0.73 (0.55, 0.97) | 0.028 |
|  | Community type |  |  |  |  |  |  |  |  |  |  |  |  |  |  |
|  | Agrarian | ref | ref |  | ref | ref |  | ref | ref |  | ref | ref |  | ref | ref |
|  | Fishing | 0.69 (0.58, 0.82) | <0.0001 |  | 0.77 (0.65, 0.92) | 0.0033 |  | 0.74 (0.64, 0.87) | 0.00019 |  | 0.75 (0.63, 0.9) | 0.0019 |  | 0.54 (0.42, 0.68) | <0.0001 |
|  | Trading | 0.99 (0.82, 1.2) | 0.91 |  | 1.06 (0.87, 1.29) | 0.56 |  | 1.11 (0.94, 1.32) | 0.22 |  | 1.06 (0.87, 1.3) | 0.55 |  | 0.99 (0.77, 1.26) | 0.92 |
|  | Sex |  |  |  |  |  |  |  |  |  |  |  |  |  |  |
|  | F | ref | ref |  | ref | ref |  | ref | ref |  | ref | ref |  | ref | ref |
|  | M | 0.85 (0.72, 0.99) | 0.037 |  | 0.86 (0.73, 1.01) | 0.058 |  | 0.9 (0.78, 1.03) | 0.14 |  | 0.86 (0.73, 1.01) | 0.071 |  | 0.87 (0.7, 1.07) | 0.18 |
|  | Log_10_ VL | 2.11 (1.9, 2.35) | <0.0001 |  | 2.14 (1.93, 2.39) | <0.0001 |  | 1.78 (1.61, 1.96) | <0.0001 |  | 2.2 (1.97, 2.45) | <0.0001 |  | 2.81 (2.46, 3.22) | <0.0001 |
| 2015 | (Intercept) | 0.42 (0.35, 0.52) | <0.0001 |  | 0.29 (0.23, 0.36) | <0.0001 |  | 0.4 (0.32, 0.49) | <0.0001 |  | 0.24 (0.19, 0.31) | <0.0001 |  | 0.24 (0.18, 0.3) | <0.0001 |
|  | Age category |  |  |  |  |  |  |  |  |  |  |  |  |  |  |
|  | (14,24] | ref | ref |  | ref | ref |  | ref | ref |  | ref | ref |  | ref | ref |
|  | (24,34] | 0.97 (0.9, 1.04) | 0.32 |  | 0.91 (0.84, 0.98) | 0.019 |  | 0.95 (0.89, 1.02) | 0.13 |  | 0.93 (0.85, 1.01) | 0.096 |  | 0.94 (0.86, 1.03) | 0.18 |
|  | (34,49] | 0.97 (0.89, 1.04) | 0.38 |  | 0.95 (0.87, 1.03) | 0.21 |  | 0.98 (0.91, 1.06) | 0.65 |  | 0.97 (0.89, 1.07) | 0.58 |  | 0.98 (0.9, 1.08) | 0.72 |
|  | Community type |  |  |  |  |  |  |  |  |  |  |  |  |  |  |
|  | Agrarian | ref | ref |  | ref | ref |  | ref | ref |  | ref | ref |  | ref | ref |
|  | Fishing | 1.03 (0.96, 1.09) | 0.42 |  | 1.01 (0.94, 1.09) | 0.74 |  | 1.01 (0.95, 1.08) | 0.65 |  | 1 (0.93, 1.07) | 0.93 |  | 1 (0.93, 1.07) | 0.92 |
|  | Trading | 1.01 (0.94, 1.09) | 0.76 |  | 1.06 (0.97, 1.15) | 0.18 |  | 1.04 (0.96, 1.12) | 0.35 |  | 1.05 (0.96, 1.14) | 0.32 |  | 1.03 (0.94, 1.13) | 0.55 |
|  | Sex |  |  |  |  |  |  |  |  |  |  |  |  |  |  |
|  | F | ref | ref |  | ref | ref |  | ref | ref |  | ref | ref |  | ref | ref |
|  | M | 0.97 (0.91, 1.02) | 0.23 |  | 0.97 (0.91, 1.04) | 0.38 |  | 0.94 (0.89, 1) | 0.041 |  | 0.96 (0.9, 1.03) | 0.22 |  | 0.96 (0.89, 1.02) | 0.2 |
|  | Treatment |  |  |  |  |  |  |  |  |  |  |  |  |  |  |
|  | Treatment-experienced | ref | ref |  | ref | ref |  | ref | ref |  | ref | ref |  | ref | ref |
|  | Pre-treatment | 1.07 (1, 1.16) | 0.061 |  | 1.05 (0.97, 1.14) | 0.2 |  | 1.1 (1.02, 1.19) | 0.014 |  | 1.1 (1.01, 1.2) | 0.036 |  | 1.1 (1.01, 1.21) | 0.032 |
|  | Log_10_ VL | 1.17 (1.12, 1.22) | <0.0001 |  | 1.28 (1.22, 1.34) | <0.0001 |  | 1.19 (1.14, 1.24) | <0.0001 |  | 1.31 (1.25, 1.38) | <0.0001 |  | 1.32 (1.25, 1.39) | <0.0001 |
| 2017 | (Intercept) | 0.76 (0.62, 0.92) | 0.0049 |  | 0.64 (0.51, 0.79) | <0.0001 |  | 0.76 (0.62, 0.92) | 0.0048 |  | 0.63 (0.51, 0.79) | <0.0001 |  | 0.63 (0.51, 0.79) | <0.0001 |
|  | Age category |  |  |  |  |  |  |  |  |  |  |  |  |  |  |
|  | (14,24] | ref | ref |  | ref | ref |  | ref | ref |  | ref | ref |  | ref | ref |
|  | (24,34] | 0.98 (0.91, 1.05) | 0.5 |  | 0.94 (0.88, 1.02) | 0.14 |  | 0.96 (0.9, 1.04) | 0.31 |  | 0.95 (0.88, 1.02) | 0.15 |  | 0.95 (0.88, 1.02) | 0.15 |
|  | (34,49] | 0.93 (0.86, 1.02) | 0.12 |  | 0.9 (0.82, 0.98) | 0.021 |  | 0.93 (0.86, 1.01) | 0.1 |  | 0.9 (0.82, 0.98) | 0.018 |  | 0.9 (0.82, 0.98) | 0.018 |
|  | Community type |  |  |  |  |  |  |  |  |  |  |  |  |  |  |
|  | Agrarian | ref | ref |  | ref | ref |  | ref | ref |  | ref | ref |  | ref | ref |
|  | Fishing | 1.03 (0.96, 1.11) | 0.44 |  | 1.05 (0.97, 1.14) | 0.2 |  | 1.04 (0.97, 1.12) | 0.3 |  | 1.05 (0.97, 1.14) | 0.19 |  | 1.05 (0.97, 1.14) | 0.19 |
|  | Trading | 1.03 (0.94, 1.11) | 0.55 |  | 1.04 (0.95, 1.13) | 0.42 |  | 1.04 (0.96, 1.13) | 0.36 |  | 1.03 (0.94, 1.13) | 0.53 |  | 1.03 (0.94, 1.13) | 0.53 |
|  | Sex |  |  |  |  |  |  |  |  |  |  |  |  |  |  |
|  | F | ref | ref |  | ref | ref |  | ref | ref |  | ref | ref |  | ref | ref |
|  | M | 0.97 (0.92, 1.04) | 0.42 |  | 0.97 (0.91, 1.04) | 0.36 |  | 0.97 (0.91, 1.03) | 0.37 |  | 0.97 (0.9, 1.03) | 0.3 |  | 0.97 (0.9, 1.03) | 0.3 |
|  | Treatment |  |  |  |  |  |  |  |  |  |  |  |  |  |  |
|  | Treatment-experienced | ref | ref |  | ref | ref |  | ref | ref |  | ref | ref |  | ref | ref |
|  | Pre-treatment | 1.07 (0.99, 1.15) | 0.089 |  | 1.04 (0.97, 1.13) | 0.26 |  | 1.06 (0.99, 1.14) | 0.12 |  | 1.05 (0.97, 1.14) | 0.2 |  | 1.05 (0.97, 1.14) | 0.2 |
|  | Log10 VL | 1.02 (0.98, 1.07) | 0.26 |  | 1.07 (1.02, 1.12) | 0.0053 |  | 1.03 (0.98, 1.07) | 0.24 |  | 1.07 (1.02, 1.12) | 0.0052 |  | 1.07 (1.02, 1.12) | 0.0052 |
| 2019 | (Intercept) | 1.21 (0.48, 3.06) | 0.69 |  | 1.04 (0.4, 2.65) | 0.94 |  | 1.52 (0.61, 3.75) | 0.37 |  | 1.04 (0.4, 2.65) | 0.94 |  | 1.04 (0.4, 2.65) | 0.94 |
|  | Age category |  |  |  |  |  |  |  |  |  |  |  |  |  |  |
|  | (14,24] | ref | ref |  | ref | ref |  | ref | ref |  | ref | ref |  | ref | ref |
|  | (24,34] | 0.94 (0.67, 1.34) | 0.75 |  | 0.98 (0.69, 1.41) | 0.92 |  | 0.93 (0.66, 1.29) | 0.65 |  | 0.98 (0.69, 1.41) | 0.92 |  | 0.98 (0.69, 1.41) | 0.92 |
|  | (34,49] | 0.96 (0.66, 1.4) | 0.83 |  | 0.97 (0.66, 1.44) | 0.9 |  | 0.94 (0.66, 1.36) | 0.76 |  | 0.97 (0.66, 1.44) | 0.9 |  | 0.97 (0.66, 1.44) | 0.9 |
|  | Community type |  |  |  |  |  |  |  |  |  |  |  |  |  |  |
|  | Agrarian | ref | ref |  | ref | ref |  | ref | ref |  | ref | ref |  | ref | ref |
|  | Fishing | 1.3 (0.95, 1.77) | 0.1 |  | 1.25 (0.91, 1.72) | 0.18 |  | 1.37 (1.01, 1.86) | 0.042 |  | 1.25 (0.91, 1.72) | 0.18 |  | 1.25 (0.91, 1.72) | 0.18 |
|  | Trading | 0.92 (0.64, 1.33) | 0.67 |  | 0.93 (0.65, 1.34) | 0.7 |  | 0.92 (0.64, 1.32) | 0.64 |  | 0.93 (0.65, 1.34) | 0.7 |  | 0.93 (0.65, 1.34) | 0.7 |
|  | Sex |  |  |  |  |  |  |  |  |  |  |  |  |  |  |
|  | F | ref | ref |  | ref | ref |  | ref | ref |  | ref | ref |  | ref | ref |
|  | M | 0.89 (0.68, 1.18) | 0.42 |  | 0.89 (0.67, 1.18) | 0.42 |  | 0.91 (0.69, 1.19) | 0.49 |  | 0.89 (0.67, 1.18) | 0.42 |  | 0.89 (0.67, 1.18) | 0.42 |
|  | Log10 VL | 0.78 (0.64, 0.96) | 0.017 |  | 0.81 (0.66, 0.99) | 0.038 |  | 0.75 (0.61, 0.91) | 0.0035 |  | 0.81 (0.66, 0.99) | 0.038 |  | 0.81 (0.66, 0.99) | 0.038 |

Estimates were generated using robust log-Poisson regression specific to each survey round (row sections) and outcome measure (columns). 2012, 2014, and 2019 survey rounds include only pre-treatment PLHIV. 95% confidence intervals indicate the Wald confidence interval around the mean value in each category. *p*-values that coefficients are different from 0 at the $\alpha=0.05$ level were calculated using the Wald method. PLHIV = people living with HIV. NNRTI = non-nucleoside reverse transcriptase inhibitors. NRTI = nucleoside reverse transcriptase inhibitors. PI = protease inhibitors.

**Supplementary Table 6: Demographics of study participants by survey round**

|  | **Survey round** | | | | |
| --- | --- | --- | --- | --- | --- |
|  | **2012** | **2014** | **2015** | **2017** | **2019** |
| Overall | 17167 | 17992 | 19336 | 19803 | 19324 |
| Age (median [IQR]) | 28 [13] | 29 [15] | 29 [14] | 29 [16] | 29 [17] |
| Age Category |  |  |  |  |  |
| (14,24] | 6026 (35.1%) | 6401 (35.58%) | 6818 (35.26%) | 6974 (35.22%) | 6691 (34.63%) |
| (24,34] | 6429 (37.45%) | 6291 (34.97%) | 6414 (33.17%) | 6370 (32.17%) | 5929 (30.68%) |
| (34,50] | 4712 (27.45%) | 5300 (29.46%) | 6104 (31.57%) | 6459 (32.62%) | 6704 (34.69%) |
| Community type |  |  |  |  |  |
| Agrarian | 7877 (45.88%) | 8197 (45.56%) | 8808 (45.55%) | 8747 (44.17%) | 8848 (45.79%) |
| Fishing | 3876 (22.58%) | 3930 (21.84%) | 4238 (21.92%) | 4756 (24.02%) | 4081 (21.12%) |
| Trading | 5414 (31.54%) | 5865 (32.6%) | 6290 (32.53%) | 6300 (31.81%) | 6395 (33.09%) |
| Sex |  |  |  |  |  |
| F | 9240 (53.82%) | 9672 (53.76%) | 10452 (54.05%) | 10552 (53.28%) | 10448 (54.07%) |
| M | 7927 (46.18%) | 8320 (46.24%) | 8884 (45.95%) | 9251 (46.72%) | 8876 (45.93%) |

Percentages represent the proportion of total participant-visits in each survey round belonging to each sub-category.

**Supplementary Table 7: Demographics of participants living with HIV by survey round**

|  | **Survey round** | | | | |
| --- | --- | --- | --- | --- | --- |
|  | **2012** | **2014** | **2015** | **2017** | **2019** |
| Overall | 3498 | 3388 | 3615 | 3636 | 3323 |
| Age (median [IQR]) | 32 [10] | 33 [12] | 34 [11] | 34 [11] | 36 [12] |
| Age Category |  |  |  |  |  |
| (14,24] | 531 (15.18%) | 478 (14.11%) | 453 (12.53%) | 372 (10.23%) | 264 (7.94%) |
| (24,34] | 1663 (47.54%) | 1469 (43.36%) | 1481 (40.97%) | 1453 (39.96%) | 1205 (36.26%) |
| (34,50] | 1304 (37.28%) | 1441 (42.53%) | 1681 (46.5%) | 1811 (49.81%) | 1854 (55.79%) |
| Community type |  |  |  |  |  |
| Agrarian | 1091 (31.19%) | 1081 (31.91%) | 1141 (31.56%) | 1072 (29.48%) | 1067 (32.11%) |
| Fishing | 1599 (45.71%) | 1519 (44.83%) | 1597 (44.18%) | 1736 (47.74%) | 1500 (45.14%) |
| Trading | 808 (23.1%) | 788 (23.26%) | 877 (24.26%) | 828 (22.77%) | 756 (22.75%) |
| Sex |  |  |  |  |  |
| F | 2169 (62.01%) | 2103 (62.07%) | 2297 (63.54%) | 2242 (61.66%) | 2125 (63.95%) |
| M | 1329 (37.99%) | 1285 (37.93%) | 1318 (36.46%) | 1394 (38.34%) | 1198 (36.05%) |

Percentages represent the proportion of total participant-visits in each survey round belonging to each sub-category.

**Supplementary Table 8: Prevalence of PLHIV, viremic PLHIV, viremic pre-treatment PLHIV, and viremic treatment-experienced PLHIV by survey round**

|  | **PLHIV** | | | | |  | **Viremic PLHIV** | | | |
| --- | --- | --- | --- | --- | --- | --- | --- | --- | --- | --- |
| **Survey round** | ***n*** | **Obs (%)** | **Prev. % (95% CI)** | **Prev. ratio (95% CI)** | ***p*-value** |  | **Obs (%)** | **Prev. % (95% CI)** | **Prev. ratio (95% CI)** | ***p*-value** |
| 2012 | 17167 | 3498 (20.38%) | 20.38 (19.78, 20.99) | ref | ref |  |  |  |  |  |
| 2014 | 17992 | 3388 (18.83%) | 18.83 (18.27, 19.41) | 0.92 (0.9, 0.95) | <0.0001 |  | 1464 (8.14%) | 8.14 (7.75, 8.55) | ref | ref |
| 2015 | 19336 | 3615 (18.7%) | 18.7 (18.15, 19.25) | 0.92 (0.89, 0.95) | <0.0001 |  | 1026 (5.31%) | 5.31 (5, 5.63) | 0.65 (0.61, 0.69) | <0.0001 |
| 2017 | 19803 | 3636 (18.36%) | 18.36 (17.83, 18.91) | 0.9 (0.87, 0.93) | <0.0001 |  | 728 (3.68%) | 3.68 (3.42, 3.95) | 0.45 (0.42, 0.49) | <0.0001 |
| 2019 | 19324 | 3323 (17.2%) | 17.2 (16.67, 17.74) | 0.84 (0.81, 0.87) | <0.0001 |  | 453 (2.34%) | 2.34 (2.14, 2.57) | 0.29 (0.26, 0.32) | <0.0001 |
|  | **Viremic pre-treatment PLHIV** | | | | |  | **Viremic treatment-experienced PLHIV** | | | |
| **Survey round** | ***n*** | **Obs (%)** | **Prev. % (95% CI)** | **Prev. ratio (95% CI)** | ***p*-value** |  | **Obs (%)** | **Prev. % (95% CI)** | **Prev. ratio (95% CI)** | ***p*-value** |
| 2012 | 17167 | 1985 (11.56%) | 11.56 (11.09, 12.05) | ref | ref |  |  |  |  |  |
| 2014 | 17992 | 1318 (7.33%) | 7.33 (6.95, 7.72) | 0.63 (0.6, 0.67) | <0.0001 |  | 146 (0.81%) | 0.81 (0.69, 0.95) | ref | ref |
| 2015 | 19336 | 822 (4.25%) | 4.25 (3.98, 4.55) | 0.37 (0.34, 0.4) | <0.0001 |  | 204 (1.06%) | 1.06 (0.92, 1.21) | 1.3 (1.09, 1.56) | 0.0041 |
| 2017 | 19803 | 502 (2.53%) | 2.53 (2.33, 2.76) | 0.22 (0.2, 0.24) | <0.0001 |  | 226 (1.14%) | 1.14 (1, 1.3) | 1.41 (1.16, 1.71) | 0.00058 |
| 2019 | 19324 | 288 (1.49%) | 1.49 (1.33, 1.67) | 0.13 (0.11, 0.15) | <0.0001 |  | 165 (0.85%) | 0.85 (0.73, 0.99) | 1.05 (0.85, 1.31) | 0.64 |

Estimates were generated using log-Poisson regression with robust standard errors with survey round as a predictor variable. Generalized estimating equations with correlation structure selection by Quasi Information Criterion for each outcome (all independent). Confidence intervals indicate the Wald confidence interval around the mean value in each category. *p*-values that coefficients are different from 0 at the $\alpha=0.05$ level were calculated using the Wald method. PLHIV = people living with HIV.

**Supplementary Table 9: Number of participant-visits contributed by PLHIV with history of treatment**

|  | **PLHIV** | |
| --- | --- | --- |
|  | **Participant-visits** | **Treatment-experienced** |
| Overall | 17460 | 10745 (61.54%) |
| Survey round |  |  |
| 2012 | 3498 | 876 (25.04%) |
| 2014 | 3388 | 1674 (49.41%) |
| 2015 | 3615 | 2451 (67.8%) |
| 2017 | 3636 | 2911 (80.06%) |
| 2019 | 3323 | 2833 (85.25%) |

Percentages represent the proportion of total participant-visits contributed by PLHIV in each survey round with self-reported history of antiretroviral therapy.

**Supplementary Table 10: Number of viremic participant-visits from which deep-sequence based identification of drug resistance mutations was attempted**

|  | **Viremic PLWHIV** | | |  | **Viremic pre-treatment PLWHIV** | | |  | **Viremic treatment-experienced PLWHIV** | | |
| --- | --- | --- | --- | --- | --- | --- | --- | --- | --- | --- | --- |
|  | **Participant-visits** | **Deep-sequence based DRM attempted (%)** | ***p-*value** |  | **Participant-visits** | **Deep-sequence based DRM attempted (%)** | ***p-*value** |  | **Participant-visits** | **Deep-sequence based DRM attempted (%)** | ***p-*value** |
| Overall | 5691 | 4523 (79.48%) |  |  | 4915 | 4094 (83.3%) |  |  | 776 | 429 (55.28%) |  |
| Age Category |  |  | 0.019 |  |  |  | 0.076 |  |  |  | 0.82 |
| (14,24] | 1111 | 950 (85.51%) |  |  | 997 | 884 (88.67%) |  |  | 114 | 66 (57.89%) |  |
| (24,34] | 2811 | 2235 (79.51%) |  |  | 2453 | 2034 (82.92%) |  |  | 358 | 201 (56.15%) |  |
| (34,50] | 1769 | 1338 (75.64%) |  |  | 1465 | 1176 (80.27%) |  |  | 304 | 162 (53.29%) |  |
| Community type |  |  | 0.55 |  |  |  | 0.26 |  |  |  | 0.069 |
| Agrarian | 1621 | 1287 (79.4%) |  |  | 1417 | 1190 (83.98%) |  |  | 204 | 97 (47.55%) |  |
| Fishing | 2860 | 2246 (78.53%) |  |  | 2454 | 1998 (81.42%) |  |  | 406 | 248 (61.08%) |  |
| Trading | 1210 | 990 (81.82%) |  |  | 1044 | 906 (86.78%) |  |  | 166 | 84 (50.6%) |  |
| Sex |  |  | 0.14 |  |  |  | 0.055 |  |  |  | 0.77 |
| F | 3008 | 2440 (81.12%) |  |  | 2564 | 2198 (85.73%) |  |  | 444 | 242 (54.5%) |  |
| M | 2683 | 2083 (77.64%) |  |  | 2351 | 1896 (80.65%) |  |  | 332 | 187 (56.33%) |  |
| Survey round |  |  | 0.0005 |  |  |  | 0.0005 |  |  |  | 0.0005 |
| 2012 | 2020 | 1971 (97.57%) |  |  | 1985 | 1969 (99.19%) |  |  | 35 | 2 (5.71%) |  |
| 2014 | 1464 | 798 (54.51%) |  |  | 1318 | 794 (60.24%) |  |  | 146 | 4 (2.74%) |  |
| 2015 | 1026 | 948 (92.4%) |  |  | 822 | 764 (92.94%) |  |  | 204 | 184 (90.2%) |  |
| 2017 | 728 | 635 (87.23%) |  |  | 502 | 445 (88.65%) |  |  | 226 | 190 (84.07%) |  |
| 2019 | 453 | 171 (37.75%) |  |  | 288 | 122 (42.36%) |  |  | 165 | 49 (29.7%) |  |
| Viral load (log10 copies/mL) |  |  | 0.0005 |  |  |  | 0.0005 |  |  |  | 0.76 |
| (3,4] | 2100 | 1647 (78.43%) |  |  | 1747 | 1445 (82.71%) |  |  | 353 | 202 (57.22%) |  |
| (4,5] | 2231 | 1696 (76.02%) |  |  | 1898 | 1515 (79.82%) |  |  | 333 | 181 (54.35%) |  |
| (5,Inf] | 598 | 428 (71.57%) |  |  | 508 | 382 (75.2%) |  |  | 90 | 46 (51.11%) |  |
| missing | 762 | 752 (98.69%) |  |  | 762 | 752 (98.69%) |  |  |  |  |  |

Percentages show the percentage of all participant-visits within a category (columns) that have successful genotyping for at least one drug. *P-*values calculated using $\chi^{2}$ goodness-of-fit test with Monte Carlo simulation. Viremic PLHIV is a sum of viremic pre-treatment PLHIV and viremic treatment-experienced PLHIV. Viral load measurements were available for only a subset of PLHIV in the 2012 survey roads. Missing viral load measurements for pre-treatment PLHIV in the 2012 survey round were imputed. All other participant-visits with missing viral load measurements were dropped. PLHIV = people living with HIV.

**Supplementary Table 11: Sequencing approach used to generate sequence data by survey round**

|  | **Survey round** | | | | |
| --- | --- | --- | --- | --- | --- |
|  | **2012** | **2014** | **2015** | **2017** | **2019** |
| Overall | 2005 | 799 | 948 | 636 | 171 |
| Sequencing approach |  |  |  |  |  |
| amplicon | 1867 (94.72%) | 607 (76.07%) | 6 (0.63%) |  |  |
| veSeq-HIV | 104 (5.28%) | 191 (23.93%) | 942 (99.37%) | 635 (100%) | 171 (100%) |

**Supplementary Table 12: Number of viremic participant-visits with attempted deep-sequence based identification of drug resistance mutations that had successful genotyping for at least one drug**

|  | **Viremic PLWHIV** | | |  | **Viremic pre-treatment PLWHIV** | | |  | **Viremic treatment-experienced PLWHIV** | | |
| --- | --- | --- | --- | --- | --- | --- | --- | --- | --- | --- | --- |
|  | **Participant-visits** | **Genotype data for ≥1 drug (%)** | ***p-*value** |  | **Participant-visits** | **Genotype data for ≥1 drug (%)** | ***p-*value** |  | **Participant-visits** | **Genotype data for ≥1 drug (%)** | ***p-*value** |
| Overall | 4523 | 4072 (90.03%) |  |  | 4094 | 3655 (89.28%) |  |  | 429 | 417 (97.2%) |  |
| Age Category |  |  | 0.83 |  |  |  | 0.84 |  |  |  | 0.98 |
| (14,24] | 950 | 840 (88.42%) |  |  | 884 | 775 (87.67%) |  |  | 66 | 65 (98.48%) |  |
| (24,34] | 2235 | 2020 (90.38%) |  |  | 2034 | 1824 (89.68%) |  |  | 201 | 196 (97.51%) |  |
| (34,50] | 1338 | 1212 (90.58%) |  |  | 1176 | 1056 (89.8%) |  |  | 162 | 156 (96.3%) |  |
| Community type |  |  | 0.83 |  |  |  | 0.84 |  |  |  | 0.98 |
| Agrarian | 1287 | 1172 (91.06%) |  |  | 1190 | 1077 (90.5%) |  |  | 97 | 95 (97.94%) |  |
| Fishing | 2246 | 2023 (90.07%) |  |  | 1998 | 1781 (89.14%) |  |  | 248 | 242 (97.58%) |  |
| Trading | 990 | 877 (88.59%) |  |  | 906 | 797 (87.97%) |  |  | 84 | 80 (95.24%) |  |
| Sex |  |  | 0.37 |  |  |  | 0.31 |  |  |  | 0.88 |
| F | 2440 | 2168 (88.85%) |  |  | 2198 | 1931 (87.85%) |  |  | 242 | 237 (97.93%) |  |
| M | 2083 | 1904 (91.41%) |  |  | 1896 | 1724 (90.93%) |  |  | 187 | 180 (96.26%) |  |
| Survey round |  |  | 0.0015 |  |  |  | 0.0005 |  |  |  | 0.97 |
| 2012 | 1971 | 1694 (85.95%) |  |  | 1969 | 1693 (85.98%) |  |  | 2 | 1 (50%) |  |
| 2014 | 798 | 657 (82.33%) |  |  | 794 | 653 (82.24%) |  |  | 4 | 4 (100%) |  |
| 2015 | 948 | 921 (97.15%) |  |  | 764 | 744 (97.38%) |  |  | 184 | 177 (96.2%) |  |
| 2017 | 635 | 630 (99.21%) |  |  | 445 | 443 (99.55%) |  |  | 190 | 187 (98.42%) |  |
| 2019 | 171 | 170 (99.42%) |  |  | 122 | 122 (100%) |  |  | 49 | 48 (97.96%) |  |
| Viral load (log10 copies/mL) |  |  | 0.0005 |  |  |  | 0.0005 |  |  |  | 0.91 |
| (3,4] | 1647 | 1359 (82.51%) |  |  | 1445 | 1167 (80.76%) |  |  | 202 | 192 (95.05%) |  |
| (4,5] | 1696 | 1630 (96.11%) |  |  | 1515 | 1451 (95.78%) |  |  | 181 | 179 (98.9%) |  |
| (5,Inf] | 428 | 421 (98.36%) |  |  | 382 | 375 (98.17%) |  |  | 46 | 46 (100%) |  |
| missing | 752 | 662 (88.03%) |  |  | 752 | 662 (88.03%) |  |  |  |  |  |
| Sequencing approach |  |  | 0.0005 |  |  |  | 0.0005 |  |  |  | 0.83 |
| amplicon | 2480 | 2106 (84.92%) |  |  | 2474 | 2101 (84.92%) |  |  | 6 | 5 (83.33%) |  |
| veSEQ-HIV | 2043 | 1966 (96.23%) |  |  | 1620 | 1554 (95.93%) |  |  | 423 | 412 (97.4%) |  |

Excludes participant-visits on which deep-sequence based identification of drug resistance mutations was not attempted. Percentages show the percentage of all participant-visits within a category (columns) that have successful genotyping for at least one drug. *P-*values calculated using $\chi^{2}$ goodness-of-fit test with Monte Carlo simulation. Viremic PLHIV is a sum of viremic pre-treatment PLHIV and viremic treatment-experienced PLHIV. Viral load measurements were available for only a subset of PLHIV in the 2012 survey roads. Missing viral load measurements for pre-treatment PLHIV in the 2012 survey round were imputed. All other participant-visits with missing viral load measurements were dropped. PLHIV = people living with HIV.

**Supplementary Table 13: Deep-sequencing quality summary statistics among participant-visits with successful genotyping for at least one drug**

|  | **Quantile** | | | | |
| --- | --- | --- | --- | --- | --- |
|  | **0.025** | **0.25** | **0.5** | **0.75** | **0.975** |
| HIV reads | 2025 | 32681 | 97973 | 236689 | 2210972.5 |
| Mapped reads | 316 | 4893 | 17912 | 49666 | 168641 |
| Insert size (quantile) | |  |  |  |  |
| 0.05 | 81 | 125 | 134 | 153 | 178 |
| 0.5 | 185.5 | 240 | 267 | 366 | 419 |
| 0.95 | 467.5 | 517 | 567 | 949 | 1104 |
| Duplicate reads (%) | 32.42 | 57.67 | 74.65 | 91.07 | 95.75 |
| Length | 1287 | 3544 | 7215 | 8938 | 9040 |

**Supplementary Table 14: Number of viremic participant-visits with attempted deep-sequence based identification of drug resistance mutations that had successful genotyping for all INSTIs**

|  | **Viremic PLWHIV** | | |  | **Viremic pre-treatment PLWHIV** | | |  | **Viremic treatment-experienced PLWHIV** | | |
| --- | --- | --- | --- | --- | --- | --- | --- | --- | --- | --- | --- |
|  | **Participant-visits** | **Genotype data for all INSTIs (%)** | ***p-*value** |  | **Participant-visits** | **Genotype data for all INSTIs (%)** | ***p-*value** |  | **Participant-visits** | **Genotype data for all INSTIs (%)** | ***p-*value** |
| Overall | 4523 | 2578 (57%) |  |  | 4094 | 2168 (52.96%) |  |  | 429 | 410 (95.57%) |  |
| Age Category |  |  | 0.21 |  |  |  | 0.41 |  |  |  | 0.96 |
| (14,24] | 950 | 527 (55.47%) |  |  | 884 | 463 (52.38%) |  |  | 66 | 64 (96.97%) |  |
| (24,34] | 2235 | 1248 (55.84%) |  |  | 2034 | 1054 (51.82%) |  |  | 201 | 194 (96.52%) |  |
| (34,50] | 1338 | 803 (60.01%) |  |  | 1176 | 651 (55.36%) |  |  | 162 | 152 (93.83%) |  |
| Community type |  |  | 0.008 |  |  |  | 0.0005 |  |  |  | 0.86 |
| Agrarian | 1287 | 794 (61.69%) |  |  | 1190 | 700 (58.82%) |  |  | 97 | 94 (96.91%) |  |
| Fishing | 2246 | 1205 (53.65%) |  |  | 1998 | 965 (48.3%) |  |  | 248 | 240 (96.77%) |  |
| Trading | 990 | 579 (58.48%) |  |  | 906 | 503 (55.52%) |  |  | 84 | 76 (90.48%) |  |
| Sex |  |  | 0.01 |  |  |  | 0.0025 |  |  |  | 0.89 |
| F | 2440 | 1326 (54.34%) |  |  | 2198 | 1093 (49.73%) |  |  | 242 | 233 (96.28%) |  |
| M | 2083 | 1252 (60.11%) |  |  | 1896 | 1075 (56.7%) |  |  | 187 | 177 (94.65%) |  |
| Survey round |  |  | 0.0005 |  |  |  | 0.0005 |  |  |  | 0.56 |
| 2012 | 1971 | 537 (27.25%) |  |  | 1969 | 537 (27.27%) |  |  | 2 | 0 (0%) |  |
| 2014 | 798 | 335 (41.98%) |  |  | 794 | 333 (41.94%) |  |  | 4 | 2 (50%) |  |
| 2015 | 948 | 909 (95.89%) |  |  | 764 | 735 (96.2%) |  |  | 184 | 174 (94.57%) |  |
| 2017 | 635 | 627 (98.74%) |  |  | 445 | 441 (99.1%) |  |  | 190 | 186 (97.89%) |  |
| 2019 | 171 | 170 (99.42%) |  |  | 122 | 122 (100%) |  |  | 49 | 48 (97.96%) |  |
| Viral load (log10 copies/mL) |  |  | 0.0005 |  |  |  | 0.0005 |  |  |  | 0.86 |
| (3,4] | 1647 | 816 (49.54%) |  |  | 1445 | 628 (43.46%) |  |  | 202 | 188 (93.07%) |  |
| (4,5] | 1696 | 1152 (67.92%) |  |  | 1515 | 976 (64.42%) |  |  | 181 | 176 (97.24%) |  |
| (5,Inf] | 428 | 362 (84.58%) |  |  | 382 | 316 (82.72%) |  |  | 46 | 46 (100%) |  |
| missing | 752 | 248 (32.98%) |  |  | 752 | 248 (32.98%) |  |  |  |  |  |
| Sequencing approach |  |  | 0.0005 |  |  |  | 0.0005 |  |  |  | 0.3 |
| amplicon | 2480 | 629 (25.36%) |  |  | 2474 | 626 (25.3%) |  |  | 6 | 3 (50%) |  |
| veSEQ-HIV | 2043 | 1949 (95.4%) |  |  | 1620 | 1542 (95.19%) |  |  | 423 | 407 (96.22%) |  |

Excludes participant-visits on which deep-sequence based identification of drug resistance mutations was not attempted. Percentages show the percentage of all participant-visits within a category (columns) that have successful genotyping for all INSTIs. *P-*values calculated using $\chi^{2}$ goodness-of-fit test with Monte Carlo simulation. Viremic PLHIV is a sum of viremic pre-treatment PLHIV and viremic treatment-experienced PLHIV. Viral load measurements were available for only a subset of PLHIV in the 2012 survey roads. Missing viral load measurements for pre-treatment PLHIV in the 2012 survey round were imputed. All other participant-visits with missing viral load measurements were dropped. PLHIV = people living with HIV. DRM = drug resistance mutations. INSTI = integrase strand transfer inhibitor.

**Supplementary Table 15: Number of viremic participant-visits with attempted deep-sequence based identification of drug resistance mutations that had successful genotyping for all NNRTIs**

|  | **Viremic PLWHIV** | | |  | **Viremic pre-treatment PLWHIV** | | |  | **Viremic treatment-experienced PLWHIV** | | |
| --- | --- | --- | --- | --- | --- | --- | --- | --- | --- | --- | --- |
|  | **Participant-visits** | **Genotype data for all NNRTIs (%)** | ***p-*value** |  | **Participant-visits** | **Genotype data for all NNRTIs (%)** | ***p-*value** |  | **Participant-visits** | **Genotype data for all NNRTIs (%)** | ***p-*value** |
| Overall | 4523 | 3050 (67.43%) |  |  | 4094 | 2656 (64.88%) |  |  | 429 | 394 (91.84%) |  |
| Age Category |  |  | 0.78 |  |  |  | 0.89 |  |  |  | 0.94 |
| (14,24] | 950 | 629 (66.21%) |  |  | 884 | 566 (64.03%) |  |  | 66 | 63 (95.45%) |  |
| (24,34] | 2235 | 1502 (67.2%) |  |  | 2034 | 1317 (64.75%) |  |  | 201 | 185 (92.04%) |  |
| (34,50] | 1338 | 919 (68.68%) |  |  | 1176 | 773 (65.73%) |  |  | 162 | 146 (90.12%) |  |
| Community type |  |  | 0.22 |  |  |  | 0.088 |  |  |  | 0.96 |
| Agrarian | 1287 | 912 (70.86%) |  |  | 1190 | 822 (69.08%) |  |  | 97 | 90 (92.78%) |  |
| Fishing | 2246 | 1481 (65.94%) |  |  | 1998 | 1252 (62.66%) |  |  | 248 | 229 (92.34%) |  |
| Trading | 990 | 657 (66.36%) |  |  | 906 | 582 (64.24%) |  |  | 84 | 75 (89.29%) |  |
| Sex |  |  | 0.009 |  |  |  | 0.005 |  |  |  | 1 |
| F | 2440 | 1574 (64.51%) |  |  | 2198 | 1352 (61.51%) |  |  | 242 | 222 (91.74%) |  |
| M | 2083 | 1476 (70.86%) |  |  | 1896 | 1304 (68.78%) |  |  | 187 | 172 (91.98%) |  |
| Survey round |  |  | 0.0005 |  |  |  | 0.0005 |  |  |  | 0.84 |
| 2012 | 1971 | 972 (49.32%) |  |  | 1969 | 971 (49.31%) |  |  | 2 | 1 (50%) |  |
| 2014 | 798 | 425 (53.26%) |  |  | 794 | 422 (53.15%) |  |  | 4 | 3 (75%) |  |
| 2015 | 948 | 865 (91.24%) |  |  | 764 | 705 (92.28%) |  |  | 184 | 160 (86.96%) |  |
| 2017 | 635 | 623 (98.11%) |  |  | 445 | 439 (98.65%) |  |  | 190 | 184 (96.84%) |  |
| 2019 | 171 | 165 (96.49%) |  |  | 122 | 119 (97.54%) |  |  | 49 | 46 (93.88%) |  |
| Viral load (log10 copies/mL) |  |  | 0.0005 |  |  |  | 0.0005 |  |  |  | 0.41 |
| (3,4] | 1647 | 895 (54.34%) |  |  | 1445 | 723 (50.03%) |  |  | 202 | 172 (85.15%) |  |
| (4,5] | 1696 | 1356 (79.95%) |  |  | 1515 | 1179 (77.82%) |  |  | 181 | 177 (97.79%) |  |
| (5,Inf] | 428 | 393 (91.82%) |  |  | 382 | 348 (91.1%) |  |  | 46 | 45 (97.83%) |  |
| missing | 752 | 406 (53.99%) |  |  | 752 | 406 (53.99%) |  |  |  |  |  |
| Sequencing approach |  |  | 0.0005 |  |  |  | 0.0005 |  |  |  | 0.56 |
| amplicon | 2480 | 1215 (48.99%) |  |  | 2474 | 1211 (48.95%) |  |  | 6 | 4 (66.67%) |  |
| veSEQ-HIV | 2043 | 1835 (89.82%) |  |  | 1620 | 1445 (89.2%) |  |  | 423 | 390 (92.2%) |  |

Excludes participant-visits on which deep-sequence based identification of drug resistance mutations was not attempted. Percentages show the percentage of all participant-visits within a category (columns) that have successful genotyping for all NNRTIs. *P-*values calculated using $\chi^{2}$ goodness-of-fit test with Monte Carlo simulation. Viremic PLHIV is a sum of viremic pre-treatment PLHIV and viremic treatment-experienced PLHIV. Viral load measurements were available for only a subset of PLHIV in the 2012 survey roads. Missing viral load measurements for pre-treatment PLHIV in the 2012 survey round were imputed. All other participant-visits with missing viral load measurements were dropped. PLHIV = people living with HIV. NNRTI = non-nucleoside reverse transcriptase inhibitor.

**Supplementary Table 16: Number of viremic participant-visits with attempted deep-sequence based identification of drug resistance mutations that had successful genotyping for all NRTIs**

|  | **Viremic PLWHIV** | | |  | **Viremic pre-treatment PLWHIV** | | |  | **Viremic treatment-experienced PLWHIV** | | |
| --- | --- | --- | --- | --- | --- | --- | --- | --- | --- | --- | --- |
|  | **Participant-visits** | **Genotype data for all NRTIs (%)** | ***p-*value** |  | **Participant-visits** | **Genotype data for all NRTIs (%)** | ***p-*value** |  | **Participant-visits** | **Genotype data for all NRTIs (%)** | ***p-*value** |
| Overall | 4523 | 3009 (66.53%) |  |  | 4094 | 2626 (64.14%) |  |  | 429 | 383 (89.28%) |  |
| Age Category |  |  | 0.55 |  |  |  | 0.71 |  |  |  | 0.96 |
| (14,24] | 950 | 620 (65.26%) |  |  | 884 | 559 (63.24%) |  |  | 66 | 61 (92.42%) |  |
| (24,34] | 2235 | 1472 (65.86%) |  |  | 2034 | 1294 (63.62%) |  |  | 201 | 178 (88.56%) |  |
| (34,50] | 1338 | 917 (68.54%) |  |  | 1176 | 773 (65.73%) |  |  | 162 | 144 (88.89%) |  |
| Community type |  |  | 0.53 |  |  |  | 0.39 |  |  |  | 0.94 |
| Agrarian | 1287 | 884 (68.69%) |  |  | 1190 | 795 (66.81%) |  |  | 97 | 89 (91.75%) |  |
| Fishing | 2246 | 1473 (65.58%) |  |  | 1998 | 1254 (62.76%) |  |  | 248 | 219 (88.31%) |  |
| Trading | 990 | 652 (65.86%) |  |  | 906 | 577 (63.69%) |  |  | 84 | 75 (89.29%) |  |
| Sex |  |  | 0.004 |  |  |  | 0.001 |  |  |  | 1 |
| F | 2440 | 1543 (63.24%) |  |  | 2198 | 1327 (60.37%) |  |  | 242 | 216 (89.26%) |  |
| M | 2083 | 1466 (70.38%) |  |  | 1896 | 1299 (68.51%) |  |  | 187 | 167 (89.3%) |  |
| Survey round |  |  | 0.0005 |  |  |  | 0.0005 |  |  |  | 0.71 |
| 2012 | 1971 | 998 (50.63%) |  |  | 1969 | 997 (50.63%) |  |  | 2 | 1 (50%) |  |
| 2014 | 798 | 420 (52.63%) |  |  | 794 | 417 (52.52%) |  |  | 4 | 3 (75%) |  |
| 2015 | 948 | 817 (86.18%) |  |  | 764 | 665 (87.04%) |  |  | 184 | 152 (82.61%) |  |
| 2017 | 635 | 612 (96.38%) |  |  | 445 | 430 (96.63%) |  |  | 190 | 182 (95.79%) |  |
| 2019 | 171 | 162 (94.74%) |  |  | 122 | 117 (95.9%) |  |  | 49 | 45 (91.84%) |  |
| Viral load (log10 copies/mL) |  |  | 0.0005 |  |  |  | 0.0005 |  |  |  | 0.097 |
| (3,4] | 1647 | 844 (51.24%) |  |  | 1445 | 684 (47.34%) |  |  | 202 | 160 (79.21%) |  |
| (4,5] | 1696 | 1356 (79.95%) |  |  | 1515 | 1179 (77.82%) |  |  | 181 | 177 (97.79%) |  |
| (5,Inf] | 428 | 397 (92.76%) |  |  | 382 | 351 (91.88%) |  |  | 46 | 46 (100%) |  |
| missing | 752 | 412 (54.79%) |  |  | 752 | 412 (54.79%) |  |  |  |  |  |
| Sequencing approach |  |  | 0.0005 |  |  |  | 0.0005 |  |  |  | 1 |
| amplicon | 2480 | 1266 (51.05%) |  |  | 2474 | 1261 (50.97%) |  |  | 6 | 5 (83.33%) |  |
| veSEQ-HIV | 2043 | 1743 (85.32%) |  |  | 1620 | 1365 (84.26%) |  |  | 423 | 378 (89.36%) |  |

Excludes participant-visits on which deep-sequence based identification of drug resistance mutations was not attempted. Percentages show the percentage of all participant-visits within a category (columns) that have successful genotyping for all NRTIs. *P-*values calculated using $\chi^{2}$ goodness-of-fit test with Monte Carlo simulation. Viremic PLHIV is a sum of viremic pre-treatment PLHIV and viremic treatment-experienced PLHIV. Viral load measurements were available for only a subset of PLHIV in the 2012 survey roads. Missing viral load measurements for pre-treatment PLHIV in the 2012 survey round were imputed. All other participant-visits with missing viral load measurements were dropped. PLHIV = people living with HIV. NRTI = nucleoside reverse transcriptase inhibitors

**Supplementary Table 17: Number of viremic participant-visits with attempted deep-sequence based identification of drug resistance mutations that had successful genotyping for all PIs**

|  | **Viremic PLWHIV** | | |  | **Viremic pre-treatment PLWHIV** | | |  | **Viremic treatment-experienced PLWHIV** | | |
| --- | --- | --- | --- | --- | --- | --- | --- | --- | --- | --- | --- |
|  | **Participant-visits** | **Genotype data for all PIs (%)** | ***p-*value** |  | **Participant-visits** | **Genotype data for all PIs (%)** | ***p-*value** |  | **Participant-visits** | **Genotype data for all PIs (%)** | ***p-*value** |
| Overall | 4523 | 3520 (77.82%) |  |  | 4094 | 3127 (76.38%) |  |  | 429 | 393 (91.61%) |  |
| Age Category |  |  | 0.57 |  |  |  | 0.62 |  |  |  | 0.94 |
| (14,24] | 950 | 722 (76%) |  |  | 884 | 659 (74.55%) |  |  | 66 | 63 (95.45%) |  |
| (24,34] | 2235 | 1730 (77.4%) |  |  | 2034 | 1548 (76.11%) |  |  | 201 | 182 (90.55%) |  |
| (34,50] | 1338 | 1068 (79.82%) |  |  | 1176 | 920 (78.23%) |  |  | 162 | 148 (91.36%) |  |
| Community type |  |  | 0.71 |  |  |  | 0.83 |  |  |  | 0.92 |
| Agrarian | 1287 | 1005 (78.09%) |  |  | 1190 | 914 (76.81%) |  |  | 97 | 91 (93.81%) |  |
| Fishing | 2246 | 1764 (78.54%) |  |  | 1998 | 1536 (76.88%) |  |  | 248 | 228 (91.94%) |  |
| Trading | 990 | 751 (75.86%) |  |  | 906 | 677 (74.72%) |  |  | 84 | 74 (88.1%) |  |
| Sex |  |  | 0.031 |  |  |  | 0.018 |  |  |  | 0.92 |
| F | 2440 | 1834 (75.16%) |  |  | 2198 | 1611 (73.29%) |  |  | 242 | 223 (92.15%) |  |
| M | 2083 | 1686 (80.94%) |  |  | 1896 | 1516 (79.96%) |  |  | 187 | 170 (90.91%) |  |
| Survey round |  |  | 0.0005 |  |  |  | 0.0005 |  |  |  | 0.71 |
| 2012 | 1971 | 1362 (69.1%) |  |  | 1969 | 1361 (69.12%) |  |  | 2 | 1 (50%) |  |
| 2014 | 798 | 502 (62.91%) |  |  | 794 | 499 (62.85%) |  |  | 4 | 3 (75%) |  |
| 2015 | 948 | 863 (91.03%) |  |  | 764 | 706 (92.41%) |  |  | 184 | 157 (85.33%) |  |
| 2017 | 635 | 624 (98.27%) |  |  | 445 | 439 (98.65%) |  |  | 190 | 185 (97.37%) |  |
| 2019 | 171 | 169 (98.83%) |  |  | 122 | 122 (100%) |  |  | 49 | 47 (95.92%) |  |
| Viral load (log10 copies/mL) |  |  | 0.0005 |  |  |  | 0.0005 |  |  |  | 0.26 |
| (3,4] | 1647 | 1079 (65.51%) |  |  | 1445 | 910 (62.98%) |  |  | 202 | 169 (83.66%) |  |
| (4,5] | 1696 | 1505 (88.74%) |  |  | 1515 | 1327 (87.59%) |  |  | 181 | 178 (98.34%) |  |
| (5,Inf] | 428 | 412 (96.26%) |  |  | 382 | 366 (95.81%) |  |  | 46 | 46 (100%) |  |
| missing | 752 | 524 (69.68%) |  |  | 752 | 524 (69.68%) |  |  |  |  |  |
| Sequencing approach |  |  | 0.0005 |  |  |  | 0.0005 |  |  |  | 1 |
| amplicon | 2480 | 1674 (67.5%) |  |  | 2474 | 1669 (67.46%) |  |  | 6 | 5 (83.33%) |  |
| veSEQ-HIV | 2043 | 1846 (90.36%) |  |  | 1620 | 1458 (90%) |  |  | 423 | 388 (91.73%) |  |

Excludes participant-visits on which deep-sequence based identification of drug resistance mutations was not attempted. Percentages show the percentage of all participant-visits within a category (columns) that have successful genotyping for all PIs. *P-*values calculated using $\chi^{2}$ goodness-of-fit test with Monte Carlo simulation. Viremic PLHIV is a sum of viremic pre-treatment PLHIV and viremic treatment-experienced PLHIV. Viral load measurements were available for only a subset of PLHIV in the 2012 survey roads. Missing viral load measurements for pre-treatment PLHIV in the 2012 survey round were imputed. All other participant-visits with missing viral load measurements were dropped. PLWHIV = people living with HIV. PI = protease inhibitor.

**Supplementary Table 18: Number of viremic participant-visits with attempted deep-sequence based identification of drug resistance mutations that had successful genotyping for all NNRTIs, NRTIs, and PIs**

|  | **Viremic PLWHIV** | | |  | **Viremic pre-treatment PLWHIV** | | |  | **Viremic treatment-experienced PLWHIV** | | |
| --- | --- | --- | --- | --- | --- | --- | --- | --- | --- | --- | --- |
|  | **Participant-visits** | **Genotype data for all NNRTIs, NRTIs, and PIs (%)** | ***p-*value** |  | **Participant-visits** | **Genotype data for all NNRTIs, NRTIs, and PIs (%)** | ***p-*value** |  | **Participant-visits** | **Genotype data for all NNRTIs, NRTIs, and PIs (%)** | ***p-*value** |
| Overall | 4523 | 2910 (64.34%) |  |  | 4094 | 2536 (61.94%) |  |  | 429 | 374 (87.18%) |  |
| Age Category |  |  | 0.64 |  |  |  | 0.83 |  |  |  | 0.96 |
| (14,24] | 950 | 602 (63.37%) |  |  | 884 | 543 (61.43%) |  |  | 66 | 59 (89.39%) |  |
| (24,34] | 2235 | 1424 (63.71%) |  |  | 2034 | 1251 (61.5%) |  |  | 201 | 173 (86.07%) |  |
| (34,50] | 1338 | 884 (66.07%) |  |  | 1176 | 742 (63.1%) |  |  | 162 | 142 (87.65%) |  |
| Community type |  |  | 0.29 |  |  |  | 0.16 |  |  |  | 0.9 |
| Agrarian | 1287 | 865 (67.21%) |  |  | 1190 | 777 (65.29%) |  |  | 97 | 88 (90.72%) |  |
| Fishing | 2246 | 1412 (62.87%) |  |  | 1998 | 1197 (59.91%) |  |  | 248 | 215 (86.69%) |  |
| Trading | 990 | 633 (63.94%) |  |  | 906 | 562 (62.03%) |  |  | 84 | 71 (84.52%) |  |
| Sex |  |  | 0.0055 |  |  |  | 0.0035 |  |  |  | 1 |
| F | 2440 | 1491 (61.11%) |  |  | 2198 | 1280 (58.23%) |  |  | 242 | 211 (87.19%) |  |
| M | 2083 | 1419 (68.12%) |  |  | 1896 | 1256 (66.24%) |  |  | 187 | 163 (87.17%) |  |
| Survey round |  |  | 0.0005 |  |  |  | 0.0005 |  |  |  | 0.46 |
| 2012 | 1971 | 932 (47.29%) |  |  | 1969 | 931 (47.28%) |  |  | 2 | 1 (50%) |  |
| 2014 | 798 | 403 (50.5%) |  |  | 794 | 400 (50.38%) |  |  | 4 | 3 (75%) |  |
| 2015 | 948 | 802 (84.6%) |  |  | 764 | 658 (86.13%) |  |  | 184 | 144 (78.26%) |  |
| 2017 | 635 | 611 (96.22%) |  |  | 445 | 430 (96.63%) |  |  | 190 | 181 (95.26%) |  |
| 2019 | 171 | 162 (94.74%) |  |  | 122 | 117 (95.9%) |  |  | 49 | 45 (91.84%) |  |
| Viral load (log10 copies/mL) |  |  | 0.0005 |  |  |  | 0.0005 |  |  |  | 0.061 |
| (3,4] | 1647 | 805 (48.88%) |  |  | 1445 | 652 (45.12%) |  |  | 202 | 153 (75.74%) |  |
| (4,5] | 1696 | 1323 (78.01%) |  |  | 1515 | 1147 (75.71%) |  |  | 181 | 176 (97.24%) |  |
| (5,Inf] | 428 | 390 (91.12%) |  |  | 382 | 345 (90.31%) |  |  | 46 | 45 (97.83%) |  |
| missing | 752 | 392 (52.13%) |  |  | 752 | 392 (52.13%) |  |  |  |  |  |
| Sequencing approach |  |  | 0.0005 |  |  |  | 0.0005 |  |  |  | 0.68 |
| amplicon | 2480 | 1183 (47.7%) |  |  | 2474 | 1179 (47.66%) |  |  | 6 | 4 (66.67%) |  |
| veSEQ-HIV | 2043 | 1727 (84.53%) |  |  | 1620 | 1357 (83.77%) |  |  | 423 | 370 (87.47%) |  |

Excludes participant-visits on which deep-sequence based identification of drug resistance mutations was not attempted. Percentages show the percentage of all participant-visits within a category (columns) that have successful genotyping for all NNRTIs, NRTIs, and PIs. *P-*values calculated using $\chi^{2}$ goodness-of-fit test with Monte Carlo simulation. Viremic PLHIV is a sum of viremic pre-treatment PLHIV and viremic treatment-experienced PLHIV. Viral load measurements were available for only a subset of PLHIV in the 2012 survey roads. Missing viral load measurements for pre-treatment PLHIV in the 2012 survey round were imputed. All other participant-visits with missing viral load measurements were dropped. PLHIV = people living with HIV. NNRTI = non-nucleoside reverse transcriptase inhibitors. NRTI = nucleoside reverse transcriptase inhibitors. PI = protease inhibitors.

**Supplementary Table 19: Number of viremic participant-visits with attempted deep-sequence based identification of drug resistance mutations that had successful deep-sequence based genotyping for all drugs**

|  | **Viremic PLWHIV** | | |  | **Viremic pre-treatment PLWHIV** | | |  | **Viremic treatment-experienced PLWHIV** | | |
| --- | --- | --- | --- | --- | --- | --- | --- | --- | --- | --- | --- |
|  | **Participant-visits** | **Genotype data for all drugs (%)** | ***p-*value** |  | **Participant-visits** | **Genotype data for all drugs (%)** | ***p-*value** |  | **Participant-visits** | **Genotype data for all drugs (%)** | ***p-*value** |
| Overall | 4523 | 2323 (51.36%) |  |  | 4094 | 1952 (47.68%) |  |  | 429 | 371 (86.48%) |  |
| Age Category |  |  | 0.39 |  |  |  | 0.62 |  |  |  | 0.96 |
| (14,24] | 950 | 479 (50.42%) |  |  | 884 | 420 (47.51%) |  |  | 66 | 59 (89.39%) |  |
| (24,34] | 2235 | 1126 (50.38%) |  |  | 2034 | 953 (46.85%) |  |  | 201 | 173 (86.07%) |  |
| (34,50] | 1338 | 718 (53.66%) |  |  | 1176 | 579 (49.23%) |  |  | 162 | 139 (85.8%) |  |
| Community type |  |  | 0.089 |  |  |  | 0.012 |  |  |  | 0.81 |
| Agrarian | 1287 | 706 (54.86%) |  |  | 1190 | 618 (51.93%) |  |  | 97 | 88 (90.72%) |  |
| Fishing | 2246 | 1105 (49.2%) |  |  | 1998 | 890 (44.54%) |  |  | 248 | 215 (86.69%) |  |
| Trading | 990 | 512 (51.72%) |  |  | 906 | 444 (49.01%) |  |  | 84 | 68 (80.95%) |  |
| Sex |  |  | 0.0015 |  |  |  | 0.0005 |  |  |  | 0.92 |
| F | 2440 | 1170 (47.95%) |  |  | 2198 | 962 (43.77%) |  |  | 242 | 208 (85.95%) |  |
| M | 2083 | 1153 (55.35%) |  |  | 1896 | 990 (52.22%) |  |  | 187 | 163 (87.17%) |  |
| Survey round |  |  | 0.0005 |  |  |  | 0.0005 |  |  |  | 0.2 |
| 2012 | 1971 | 482 (24.45%) |  |  | 1969 | 482 (24.48%) |  |  | 2 | 0 (0%) |  |
| 2014 | 798 | 269 (33.71%) |  |  | 794 | 267 (33.63%) |  |  | 4 | 2 (50%) |  |
| 2015 | 948 | 799 (84.28%) |  |  | 764 | 656 (85.86%) |  |  | 184 | 143 (77.72%) |  |
| 2017 | 635 | 611 (96.22%) |  |  | 445 | 430 (96.63%) |  |  | 190 | 181 (95.26%) |  |
| 2019 | 171 | 162 (94.74%) |  |  | 122 | 117 (95.9%) |  |  | 49 | 45 (91.84%) |  |
| Viral load (log10 copies/mL) |  |  | 0.0005 |  |  |  | 0.0005 |  |  |  | 0.056 |
| (3,4] | 1647 | 656 (39.83%) |  |  | 1445 | 504 (34.88%) |  |  | 202 | 152 (75.25%) |  |
| (4,5] | 1696 | 1090 (64.27%) |  |  | 1515 | 916 (60.46%) |  |  | 181 | 174 (96.13%) |  |
| (5,Inf] | 428 | 356 (83.18%) |  |  | 382 | 311 (81.41%) |  |  | 46 | 45 (97.83%) |  |
| missing | 752 | 221 (29.39%) |  |  | 752 | 221 (29.39%) |  |  |  |  |  |
| Sequencing approach | |  | 0.0005 |  |  |  | 0.0005 |  |  |  | 0.19 |
| amplicon | 2480 | 597 (24.07%) |  |  | 2474 | 595 (24.05%) |  |  | 6 | 2 (33.33%) |  |
| veSEQ-HIV | 2043 | 1726 (84.48%) |  |  | 1620 | 1357 (83.77%) |  |  | 423 | 369 (87.23%) |  |

Excludes participant-visits on which deep-sequence based identification of drug resistance mutations was not attempted. Percentages show the percentage of all participant-visits within a category (columns) that have successful genotyping for all INSTIs, NNRTIs, NRTIs, and PIs. *P-*values calculated using $\chi^{2}$ goodness-of-fit test with Monte Carlo simulation. Viremic PLHIV is a sum of viremic pre-treatment PLHIV and viremic treatment-experienced PLHIV. Viral load measurements were available for only a subset of PLHIV in the 2012 survey roads. Missing viral load measurements for pre-treatment PLHIV in the 2012 survey round were imputed. All other participant-visits with missing viral load measurements were dropped. PLHIV = people living with HIV. INSTI = integrase strand transfer inhibitor. PI = protease inhibitor. NNRTI = non-nucleoside reverse transcriptase inhibitors. NRTI = nucleoside reverse transcriptase inhibitors. PI = protease inhibitors.

**Supplementary Table 20: Number of viremic participant-visits with resistant genotypes**

|  |  |  | **Survey Round** | | | | | |
| --- | --- | --- | --- | --- | --- | --- | --- | --- |
| **Treatment-status** | **Class** | **Resistance** | **all** | **2012** | **2014** | **2015** | **2017** | **2019** |
| All | INSTI | All | 2578 | 537 | 335 | 909 | 627 | 170 |
|  |  | Susceptible | 2546 (98.76%) | 530 (98.7%) | 322 (96.12%) | 901 (99.12%) | 623 (99.36%) | 170 (100%) |
|  |  | Low | 13 (0.5%) | 3 (0.56%) | 1 (0.3%) | 7 (0.77%) | 2 (0.32%) | 0 (0%) |
|  |  | Intermediate/high | 19 (0.74%) | 4 (0.74%) | 12 (3.58%) | 1 (0.11%) | 2 (0.32%) | 0 (0%) |
|  | NNRTI | All | 3050 | 972 | 425 | 865 | 623 | 165 |
|  |  | Susceptible | 2564 (84.07%) | 891 (91.67%) | 381 (89.65%) | 698 (80.69%) | 469 (75.28%) | 125 (75.76%) |
|  |  | Low | 106 (3.48%) | 33 (3.4%) | 16 (3.76%) | 30 (3.47%) | 24 (3.85%) | 3 (1.82%) |
|  |  | Intermediate/high | 380 (12.46%) | 48 (4.94%) | 28 (6.59%) | 137 (15.84%) | 130 (20.87%) | 37 (22.42%) |
|  | NRTI | All | 3009 | 998 | 420 | 817 | 612 | 162 |
|  |  | Susceptible | 2743 (91.16%) | 962 (96.39%) | 406 (96.67%) | 711 (87.03%) | 525 (85.78%) | 139 (85.8%) |
|  |  | Low | 63 (2.09%) | 17 (1.7%) | 8 (1.9%) | 19 (2.33%) | 13 (2.12%) | 6 (3.7%) |
|  |  | Intermediate/high | 203 (6.75%) | 19 (1.9%) | 6 (1.43%) | 87 (10.65%) | 74 (12.09%) | 17 (10.49%) |
|  | PI | All | 3520 | 1362 | 502 | 863 | 624 | 169 |
|  |  | Susceptible | 3407 (96.79%) | 1316 (96.62%) | 482 (96.02%) | 842 (97.57%) | 603 (96.63%) | 164 (97.04%) |
|  |  | Low | 47 (1.34%) | 17 (1.25%) | 12 (2.39%) | 7 (0.81%) | 8 (1.28%) | 3 (1.78%) |
|  |  | Intermediate/high | 66 (1.88%) | 29 (2.13%) | 8 (1.59%) | 14 (1.62%) | 13 (2.08%) | 2 (1.18%) |
| Pre-treatment | INSTI | All | 2168 | 537 | 333 | 735 | 441 | 122 |
|  |  | Susceptible | 2142 (98.8%) | 530 (98.7%) | 320 (96.1%) | 731 (99.46%) | 439 (99.55%) | 122 (100%) |
|  |  | Low | 10 (0.46%) | 3 (0.56%) | 1 (0.3%) | 4 (0.54%) | 2 (0.45%) | 0 (0%) |
|  |  | Intermediate/high | 16 (0.74%) | 4 (0.74%) | 12 (3.6%) | 0 (0%) | 0 (0%) | 0 (0%) |
|  | NNRTI | All | 2656 | 971 | 422 | 705 | 439 | 119 |
|  |  | Susceptible | 2383 (89.72%) | 890 (91.66%) | 378 (89.57%) | 628 (89.08%) | 381 (86.79%) | 106 (89.08%) |
|  |  | Low | 94 (3.54%) | 33 (3.4%) | 16 (3.79%) | 27 (3.83%) | 15 (3.42%) | 3 (2.52%) |
|  |  | Intermediate/high | 179 (6.74%) | 48 (4.94%) | 28 (6.64%) | 50 (7.09%) | 43 (9.79%) | 10 (8.4%) |
|  | NRTI | All | 2626 | 997 | 417 | 665 | 430 | 117 |
|  |  | Susceptible | 2518 (95.89%) | 961 (96.39%) | 403 (96.64%) | 633 (95.19%) | 411 (95.58%) | 110 (94.02%) |
|  |  | Low | 60 (2.28%) | 17 (1.71%) | 8 (1.92%) | 19 (2.86%) | 11 (2.56%) | 5 (4.27%) |
|  |  | Intermediate/high | 48 (1.83%) | 19 (1.91%) | 6 (1.44%) | 13 (1.95%) | 8 (1.86%) | 2 (1.71%) |
|  | PI | All | 3127 | 1361 | 499 | 706 | 439 | 122 |
|  |  | Susceptible | 3027 (96.8%) | 1315 (96.62%) | 479 (95.99%) | 690 (97.73%) | 423 (96.36%) | 120 (98.36%) |
|  |  | Low | 44 (1.41%) | 17 (1.25%) | 12 (2.4%) | 7 (0.99%) | 7 (1.59%) | 1 (0.82%) |
|  |  | Intermediate/high | 56 (1.79%) | 29 (2.13%) | 8 (1.6%) | 9 (1.27%) | 9 (2.05%) | 1 (0.82%) |
| Treatment-experienced | INSTI | All | 410 |  | 2 | 174 | 186 | 48 |
|  |  | Susceptible | 404 (98.54%) |  | 2 (100%) | 170 (97.7%) | 184 (98.92%) | 48 (100%) |
|  |  | Low | 3 (0.73%) |  | 0 (0%) | 3 (1.72%) | 0 (0%) | 0 (0%) |
|  |  | Intermediate/high | 3 (0.73%) |  | 0 (0%) | 1 (0.57%) | 2 (1.08%) | 0 (0%) |
|  | NNRTI | All | 394 | 1 | 3 | 160 | 184 | 46 |
|  |  | Susceptible | 181 (45.94%) | 1 (100%) | 3 (100%) | 70 (43.75%) | 88 (47.83%) | 19 (41.3%) |
|  |  | Low | 12 (3.05%) | 0 (0%) | 0 (0%) | 3 (1.88%) | 9 (4.89%) | 0 (0%) |
|  |  | Intermediate/high | 201 (51.02%) | 0 (0%) | 0 (0%) | 87 (54.37%) | 87 (47.28%) | 27 (58.7%) |
|  | NRTI | All | 383 | 1 | 3 | 152 | 182 | 45 |
|  |  | Susceptible | 225 (58.75%) | 1 (100%) | 3 (100%) | 78 (51.32%) | 114 (62.64%) | 29 (64.44%) |
|  |  | Low | 3 (0.78%) | 0 (0%) | 0 (0%) | 0 (0%) | 2 (1.1%) | 1 (2.22%) |
|  |  | Intermediate/high | 155 (40.47%) | 0 (0%) | 0 (0%) | 74 (48.68%) | 66 (36.26%) | 15 (33.33%) |
|  | PI | All | 393 | 1 | 3 | 157 | 185 | 47 |
|  |  | Susceptible | 380 (96.69%) | 1 (100%) | 3 (100%) | 152 (96.82%) | 180 (97.3%) | 44 (93.62%) |
|  |  | Low | 3 (0.76%) | 0 (0%) | 0 (0%) | 0 (0%) | 1 (0.54%) | 2 (4.26%) |
|  |  | Intermediate/high | 10 (2.54%) | 0 (0%) | 0 (0%) | 5 (3.18%) | 4 (2.16%) | 1 (2.13%) |

INSTI = integrase strand transfer inhibitor. PI = protease inhibitor. NNRTI = non-nucleoside reverse transcriptase inhibitors. NRTI = nucleoside reverse transcriptase inhibitors. PI = protease inhibitors.

**Supplementary Table 21: Population prevalence of viremic NNRTI, NRTI, and PI resistance by survey round**

|  | **NNRTI** | | | | |  | **NRTI** | | | | |  | **PI** | | | | |
| --- | --- | --- | --- | --- | --- | --- | --- | --- | --- | --- | --- | --- | --- | --- | --- | --- | --- |
| **Survey round** | ***n*** | **Obs (%)** | **Prev. (95% CI)** | **Prev. ratio (95% CI)** | ***p*-value** |  | ***n*** | **Obs (%)** | **Prev. (95% CI)** | **Prev. ratio (95% CI)** | ***p*-value** |  | ***n*** | **Obs (%)** | **Prev. (95% CI)** | **Prev. ratio (95% CI)** | ***p*-value** |
| 2015 | 19175 | 137 (0.71%) | 0.88 (0.74, 1.04) | ref | ref |  | 19127 | 87 (0.45%) | 0.61 (0.49, 0.75) | ref | ref |  | 19173 | 14 (0.07%) | 0.09 (0.05, 0.15) | ref | ref |
| 2017 | 19698 | 130 (0.66%) | 0.79 (0.66, 0.93) | 0.89 (0.71, 1.12) | 0.32 |  | 19687 | 74 (0.38%) | 0.46 (0.37, 0.58) | 0.76 (0.57, 1.01) | 0.063 |  | 19699 | 13 (0.07%) | 0.08 (0.04, 0.13) | 0.87 (0.42, 1.81) | 0.71 |

Estimates were generated using Poisson regression with robust standard errors with survey round as a predictor variable. Generalized estimating equations with correlation structure selection by Quasi Information Criterion value (NNRTI: independent, NRTI independent, PI: independent) were used to account for repeat participants across study rounds. 95% confidence intervals indicate the Wald confidence interval around the mean value in each category. *p*-values that coefficients are different from 0 at the $\alpha=0.05$ level were calculated using the Wald method. NNRTI = non-nucleoside reverse transcriptase inhibitors. NRTI = nucleoside reverse transcriptase inhibitors. PI = integrase inhibitors.

**Supplementary Table 22: Association between age, community type, and sex and the population prevalence of viremic NNRTI, NRTI and PI resistance adjusted by survey round**

|  |  | **NNRTI** | |  | **NRTI** | |  | **PI** | |
| --- | --- | --- | --- | --- | --- | --- | --- | --- | --- |
| **Covariate** | **Variable** | **Coeff. % (95% CI)** | ***p*-value** |  | **Coeff. % (95% CI)** | ***p*-value** |  | **Coeff. % (95% CI)** | ***p*-value** |
| Age | Intercept | 0 (0, 0.01) | ref |  | 0 (0, 0) | ref |  | 0 (0, 0) | ref |
|  | 2015 | 0.9 (0.72, 1.12) | 0.34 |  | 0.77 (0.57, 1.02) | 0.068 |  | 0.88 (0.42, 1.82) | 0.73 |
|  | age (24,34] | 3.04 (2.18, 4.25) | <0.0001 |  | 5.55 (3.3, 9.31) | <0.0001 |  | 2.81 (1.06, 7.46) | 0.039 |
|  | age (34,49] | 1.99 (1.39, 2.84) | 0.00018 |  | 3.69 (2.16, 6.31) | <0.0001 |  | 1.44 (0.48, 4.33) | 0.51 |
| Community type | Intercept | 0.01 (0, 0.01) | ref |  | 0 (0, 0.01) | ref |  | 0 (0, 0) | ref |
|  | 2015 | 0.86 (0.69, 1.08) | 0.2 |  | 0.74 (0.56, 0.98) | 0.038 |  | 0.84 (0.4, 1.73) | 0.63 |
|  | Fishing | 3.34 (2.48, 4.51) | <0.0001 |  | 3.19 (2.15, 4.73) | <0.0001 |  | 5.33 (1.91, 14.87) | 0.0014 |
|  | Trading | 1.04 (0.73, 1.48) | 0.82 |  | 1.08 (0.69, 1.69) | 0.74 |  | 1.68 (0.53, 5.34) | 0.38 |
| Sex | Intercept | 0.01 (0.01, 0.01) | ref |  | 0.01 (0.01, 0.01) | ref |  | 0 (0, 0) | ref |
|  | 2015 | 0.89 (0.71, 1.12) | 0.33 |  | 0.76 (0.57, 1.02) | 0.065 |  | 0.87 (0.42, 1.81) | 0.71 |
|  | Male | 0.74 (0.57, 0.96) | 0.022 |  | 0.68 (0.48, 0.96) | 0.028 |  | 0.88 (0.4, 1.92) | 0.74 |

**E**stimates were generated using bivariate Poisson regression with robust standard errors with survey round and an epidemiological covariate as predictor variables. Generalized estimating equations with correlation structure selection by Quasi Information Criterion value based on the univariate analysis (NNRTI: independent, NRTI independent, PI: independent) were used to account for repeat participants across study rounds. 95% confidence intervals indicate the Wald confidence interval around the mean value in each category. *p*-values that coefficients are different from 0 at the $\alpha=0.05$ level were calculated using the Wald method. NNRTI = non-nucleoside reverse transcriptase inhibitors. NRTI = nucleoside reverse transcriptase inhibitors. PI = integrase inhibitors.

**Supplementary Table 23: Population prevalence of viremic NNRTI, NRTI, and PI resistance stratified by survey round and age, community type, and sex**

|  |  | **NNRTI** | | | | |  | **NRTI** | | | | |  | **PI** | | | | |
| --- | --- | --- | --- | --- | --- | --- | --- | --- | --- | --- | --- | --- | --- | --- | --- | --- | --- | --- |
| **Covariate** | **Strata** | ***n*** | **Obs (%)** | **Prev. (95% CI)** | **Prev. ratio (95% CI)** | ***p*-value** |  | ***n*** | **Obs (%)** | **Prev. (95% CI)** | **Prev. ratio (95% CI)** | ***p*-value** |  | ***n*** | **Obs (%)** | **Prev. (95% CI)** | **Prev. ratio (95% CI)** | ***p*-value** |
| Age category | age (14,24] |  |  |  |  |  |  |  |  |  |  |  |  |  |  |  |  |  |
|  | 2015 | 6791 | 29 (0.43%) | 0.51 (0.35, 0.73) | ref | ref |  | 6786 | 10 (0.15%) | 0.19 (0.1, 0.35) | ref | ref |  | 6792 | 1 (0.01%) | 0.01 (0, 0.09) | ref | ref |
|  | 2017 | 6958 | 20 (0.29%) | 0.33 (0.21, 0.51) | 0.65 (0.37, 1.15) | 0.14 |  | 6958 | 8 (0.11%) | 0.14 (0.07, 0.27) | 0.73 (0.29, 1.86) | 0.51 |  | 6959 | 5 (0.07%) | 0.08 (0.03, 0.19) | 6.49 (0.76, 55.53) | 0.09 |
|  | age (24,34] |  |  |  |  |  |  |  |  |  |  |  |  |  |  |  |  |  |
|  | 2015 | 6334 | 63 (0.99%) | 1.22 (0.95, 1.56) | ref | ref |  | 6301 | 42 (0.67%) | 0.91 (0.67, 1.24) | ref | ref |  | 6327 | 8 (0.13%) | 0.16 (0.08, 0.31) | ref | ref |
|  | 2017 | 6321 | 72 (1.14%) | 1.34 (1.06, 1.68) | 1.1 (0.81, 1.5) | 0.55 |  | 6315 | 45 (0.71%) | 0.87 (0.65, 1.16) | 0.95 (0.65, 1.39) | 0.79 |  | 6320 | 6 (0.09%) | 0.11 (0.05, 0.24) | 0.7 (0.26, 1.88) | 0.49 |
|  | age (34,49] |  |  |  |  |  |  |  |  |  |  |  |  |  |  |  |  |  |
|  | 2015 | 6050 | 45 (0.74%) | 0.94 (0.7, 1.26) | ref | ref |  | 6040 | 35 (0.58%) | 0.77 (0.55, 1.07) | ref | ref |  | 6054 | 5 (0.08%) | 0.1 (0.04, 0.24) | ref | ref |
|  | 2017 | 6419 | 38 (0.59%) | 0.73 (0.53, 1) | 0.78 (0.52, 1.17) | 0.23 |  | 6414 | 21 (0.33%) | 0.42 (0.28, 0.65) | 0.55 (0.32, 0.94) | 0.028 |  | 6420 | 2 (0.03%) | 0.04 (0.01, 0.15) | 0.38 (0.07, 1.98) | 0.25 |
| Comm. type | Agrarian |  |  |  |  |  |  |  |  |  |  |  |  |  |  |  |  |  |
|  | 2015 | 8757 | 42 (0.48%) | 0.6 (0.45, 0.82) | ref | ref |  | 8742 | 27 (0.31%) | 0.43 (0.29, 0.62) | ref | ref |  | 8757 | 4 (0.05%) | 0.06 (0.02, 0.16) | ref | ref |
|  | 2017 | 8714 | 34 (0.39%) | 0.47 (0.34, 0.66) | 0.78 (0.51, 1.2) | 0.26 |  | 8709 | 19 (0.22%) | 0.28 (0.18, 0.43) | 0.65 (0.37, 1.11) | 0.12 |  | 8713 | 1 (0.01%) | 0.01 (0, 0.1) | 0.22 (0.02, 1.99) | 0.18 |
|  | Fishing |  |  |  |  |  |  |  |  |  |  |  |  |  |  |  |  |  |
|  | 2015 | 4164 | 66 (1.59%) | 1.88 (1.48, 2.39) | ref | ref |  | 4135 | 36 (0.87%) | 1.14 (0.82, 1.59) | ref | ref |  | 4158 | 7 (0.17%) | 0.2 (0.09, 0.41) | ref | ref |
|  | 2017 | 4706 | 68 (1.44%) | 1.71 (1.35, 2.16) | 0.91 (0.67, 1.23) | 0.54 |  | 4703 | 42 (0.89%) | 1.09 (0.8, 1.47) | 0.95 (0.64, 1.41) | 0.8 |  | 4707 | 8 (0.17%) | 0.19 (0.1, 0.39) | 0.99 (0.38, 2.54) | 0.98 |
|  | Trading |  |  |  |  |  |  |  |  |  |  |  |  |  |  |  |  |  |
|  | 2015 | 6254 | 29 (0.46%) | 0.6 (0.41, 0.86) | ref | ref |  | 6250 | 24 (0.38%) | 0.5 (0.34, 0.75) | ref | ref |  | 6258 | 3 (0.05%) | 0.05 (0.02, 0.16) | ref | ref |
|  | 2017 | 6278 | 28 (0.45%) | 0.53 (0.36, 0.76) | 0.88 (0.52, 1.48) | 0.64 |  | 6275 | 13 (0.21%) | 0.26 (0.15, 0.44) | 0.51 (0.26, 1) | 0.048 |  | 6279 | 4 (0.06%) | 0.07 (0.03, 0.2) | 1.45 (0.32, 6.63) | 0.63 |
| Sex | F |  |  |  |  |  |  |  |  |  |  |  |  | .. | .. | .. | .. | .. |
|  | 2015 | 10368 | 85 (0.82%) | 1.01 (0.81, 1.24) | ref | ref |  | 10340 | 54 (0.52%) | 0.7 (0.53, 0.91) | ref | ref |  | .. | .. | .. | .. | .. |
|  | 2017 | 10505 | 79 (0.75%) | 0.89 (0.72, 1.11) | 0.89 (0.66, 1.18) | 0.41 |  | 10499 | 48 (0.46%) | 0.56 (0.42, 0.75) | 0.81 (0.56, 1.16) | 0.25 |  | .. | .. | .. | .. | .. |
|  | M |  |  |  |  |  |  |  |  |  |  |  | .. | .. | .. | .. | .. | .. |
|  | 2015 | 8807 | 52 (0.59%) | 0.73 (0.56, 0.96) | ref | ref |  | 8787 | 33 (0.38%) | 0.51 (0.36, 0.71) | ref | ref |  | .. | .. | .. | .. | .. |
|  | 2017 | 9193 | 51 (0.55%) | 0.66 (0.51, 0.87) | 0.91 (0.63, 1.3) | 0.6 |  | 9188 | 26 (0.28%) | 0.35 (0.24, 0.52) | 0.69 (0.43, 1.11) | 0.13 |  | .. | .. | .. | .. | .. |

Estimates were generated using bivariate Poisson regression with robust standard errors with survey round as a predictor variable stratified by covariate. Estimates only generated for combination of drug and covariate that were significant in the bivariate analysis. Generalized estimating equations with correlation structure selection by Quasi Information Criterion value based on the univariate analysis (NNRTI: independent, NRTI independent, PI: independent) were used to account for repeat participants across study rounds. 95% confidence intervals indicate the Wald confidence interval around the mean value in each category. *p*-values that coefficients are different from 0 at the $\alpha=0.05$ level were calculated using the Wald method. NNRTI = non-nucleoside reverse transcriptase inhibitors. NRTI = nucleoside reverse transcriptase inhibitors. PI = integrase inhibitors.

**Table 24: Population prevalence of viremic pre-treatment NNRTI, NRTI, and PI resistance by survey round**

|  | **NNRTI** | | | | |  | **NRTI** | | | | |  | **PI** | | | | |
| --- | --- | --- | --- | --- | --- | --- | --- | --- | --- | --- | --- | --- | --- | --- | --- | --- | --- |
| **Survey round** | ***n*** | **Obs (%)** | **Prev. % (95% CI)** | **Prev. ratio (95% CI)** | ***p*-value** |  | ***n*** | **Obs (%)** | **Prev. % (95% CI)** | **Prev. ratio (95% CI)** | ***p*-value** |  | ***n*** | **Obs (%)** | **Prev. % (95% CI)** | **Prev. ratio (95% CI)** | ***p*-value** |
| 2012 | 16153 | 48 (0.3%) | 0.56 (0.42, 0.75) | ref | ref |  | 16179 | 19 (0.12%) | 0.24 (0.15, 0.37) | ref | ref |  | 16543 | 29 (0.18%) | 0.25 (0.17, 0.36) | ref | ref |
| 2014 | 17096 | 28 (0.16%) | 0.4 (0.27, 0.59) | 0.71 (0.44, 1.16) | 0.17 |  | 17091 | 6 (0.04%) | 0.1 (0.04, 0.22) | 0.41 (0.16, 1.08) | 0.071 |  | 17173 | 8 (0.05%) | 0.11 (0.05, 0.23) | 0.43 (0.19, 0.96) | 0.04 |
| 2015 | 19219 | 50 (0.26%) | 0.3 (0.23, 0.39) | 0.54 (0.36, 0.8) | 0.0023 |  | 19179 | 13 (0.07%) | 0.09 (0.05, 0.15) | 0.38 (0.19, 0.75) | 0.0057 |  | 19220 | 9 (0.05%) | 0.05 (0.03, 0.1) | 0.21 (0.1, 0.46) | <0.0001 |
| 2017 | 19740 | 43 (0.22%) | 0.25 (0.18, 0.33) | 0.44 (0.29, 0.68) | 0.00015 |  | 19731 | 8 (0.04%) | 0.05 (0.02, 0.1) | 0.21 (0.09, 0.47) | 0.0002 |  | 19740 | 9 (0.05%) | 0.05 (0.03, 0.1) | 0.21 (0.1, 0.43) | <0.0001 |
| 2019 | 19155 | 10 (0.05%) | 0.12 (0.06, 0.22) | 0.21 (0.11, 0.43) | <0.0001 |  | 19153 | 2 (0.01%) | 0.03 (0.01, 0.11) | 0.11 (0.03, 0.49) | 0.0036 |  | 19158 | 1 (0.01%) | 0.01 (0, 0.08) | 0.05 (0.01, 0.34) | 0.0025 |

Estimates were generated using Poisson regression with robust standard errors with survey round as a predictor variable. Generalized estimating equations with correlation structure selection by Quasi Information Criterion value (NNRTI: independent, NRTI: independent, PI: independent) were used to account for repeat participants across study rounds. 95% confidence intervals indicate the Wald confidence interval around the mean value in each category. *p*-values that coefficients are different from 0 at the $\alpha=0.05$ level were calculated using the Wald method. NNRTI = non-nucleoside reverse transcriptase inhibitors. NRTI = nucleoside reverse transcriptase inhibitors. PI = integrase inhibitors.

**Supplementary Table 25: Population prevalence of viremic treatment-experienced NNRTI, NRTI, and PI resistance by survey round**

|  | **NNRTI** | | | | |  | **NRTI** | | | | |  | **PI** | | | | |
| --- | --- | --- | --- | --- | --- | --- | --- | --- | --- | --- | --- | --- | --- | --- | --- | --- | --- |
| **Survey round** | ***n*** | **Obs (%)** | **Prev. % (95% CI)** | **Prev. ratio (95% CI)** | ***p*-value** |  | ***n*** | **Obs (%)** | **Prev. % (95% CI)** | **Prev. ratio (95% CI)** | ***p*-value** |  | ***n*** | **Obs (%)** | **Prev. % (95% CI)** | **Prev. ratio (95% CI)** | ***p*-value** |
| 2015 | 19292 | 87 (0.45%) | 0.58 (0.47, 0.72) | ref | ref |  | 19284 | 74 (0.38%) | 0.53 (0.42, 0.66) | ref | ref |  | 19289 | 5 (0.03%) | 0.03 (0.01, 0.08) | ref | ref |
| 2017 | 19761 | 87 (0.44%) | 0.54 (0.44, 0.66) | 0.92 (0.7, 1.22) | 0.57 |  | 19759 | 66 (0.33%) | 0.42 (0.33, 0.53) | 0.79 (0.58, 1.07) | 0.13 |  | 19762 | 4 (0.02%) | 0.02 (0.01, 0.06) | 0.7 (0.19, 2.62) | 0.6 |

Estimates were generated using Poisson regression with robust standard errors with survey round as a predictor variable. Generalized estimating equations with correlation structure selection by Quasi Information Criterion value (NNRTI: independent, NRTI: independent, PI: independent) were used to account for repeat participants across study rounds. 95% confidence intervals indicate the Wald confidence interval around the mean value in each category. *p*-values that coefficients are different from 0 at the $\alpha=0.05$ level were calculated using the Wald method. NNRTI = non-nucleoside reverse transcriptase inhibitors. NRTI = nucleoside reverse transcriptase inhibitors. PI = integrase inhibitors.

**Supplementary Table 26: Population prevalence of viremic multi-class resistance by survey round**

|  |  | **NNRTI** | | | |  | **NRTI** | | | |  | **PI** | | | |
| --- | --- | --- | --- | --- | --- | --- | --- | --- | --- | --- | --- | --- | --- | --- | --- |
| **Survey round** | ***n*** | **Obs (%)** | **Prev. % (95% CI)** | **Prev. ratio (95% CI)** | ***p*-value** |  | **Obs (%)** | **Prev. % (95% CI)** | **Prev. ratio (95% CI)** | ***p*-value** |  | **Obs (%)** | **Prev. % (95% CI)** | **Prev. ratio (95% CI)** | ***p*-value** |
| 2015 | 19336 | 50 (0.26%) | 0.33 (0.25, 0.44) | ref | ref |  | 2 (0.01%) | 0.01 (0, 0.05) | ref | ref |  | 8 (0.04%) | 0.05 (0.02, 0.1) | ref | ref |
| 2017 | 19803 | 54 (0.27%) | 0.32 (0.25, 0.42) | 0.97 (0.67, 1.4) | 0.86 |  | 0 (0%) | 0 (0, 0) | 0 (0, 0) | <0.0001 |  | 7 (0.04%) | 0.04 (0.02, 0.09) | 0.84 (0.3, 2.35) | 0.74 |
|  |  | **NNRTI & NRTI** | | | |  | **NNRTI & PI** | | | |  | **NRTI & PI** | | | |
| **Survey round** | ***n*** | **Obs (%)** | **Prev. % (95% CI)** | **Prev. ratio (95% CI)** | ***p*-value** |  | **Obs (%)** | **Prev. % (95% CI)** | **Prev. ratio (95% CI)** | ***p*-value** |  | **Obs (%)** | **Prev. % (95% CI)** | **Prev. ratio (95% CI)** | ***p*-value** |
| 2015 | 19336 | 74 (0.38%) | 0.54 (0.43, 0.68) | ref | ref |  | 0 (0%) | 0 (NA, NA) | ref | ref |  | 1 (0.01%) | 0.01 (0, 0.04) | ref | ref |
| 2017 | 19803 | 70 (0.35%) | 0.44 (0.35, 0.56) | 0.82 (0.6, 1.11) | 0.2 |  | 2 (0.01%) | 0.01 (0, 0.05) | NA (NA, NA) |  |  | 1 (0.01%) | 0.01 (0, 0.04) | 0.94 (0.06, 14.96) | 0.96 |
|  |  | **NNRTI, NRTI, & PI** | | | |  |  |  |  |  |  |  |  |  |  |
| **Survey round** | ***n*** | **Obs (%)** | **Prev. % (95% CI)** | **Prev. ratio (95% CI)** | ***p*-value** |  |  |  |  |  |  |  |  |  |  |
| 2015 | 19336 | 5 (0.03%) | 0.04 (0.02, 0.09) | ref | ref |  |  |  |  |  |  |  |  |  |  |
| 2017 | 19803 | 3 (0.02%) | 0.03 (0.01, 0.07) | 0.77 (0.34, 1.74) | 0.53 |  |  |  |  |  |  |  |  |  |  |

Estimates were generated using Poisson regression with robust standard errors with survey round as a predictor variable. Generalized estimating equations with correlation structure selection by Quasi Information Criterion value (NNRTI: independent, NRTI: independent, PI: independent, NNRTI&NRTI: independent, NNRTI&PI: independent, NRTI&PI: exchangeable, NNRTI,NRTI,&PI: exchangeable) were used to account for repeat participants across study rounds. 95% confidence intervals indicate the Wald confidence interval around the mean value in each category. *p*-values that coefficients are different from 0 at the $\alpha=0.05$ level were calculated using the Wald method. NNRTI = non-nucleoside reverse transcriptase inhibitors. NRTI = nucleoside reverse transcriptase inhibitors. PI = integrase inhibitors.

**Supplementary Table 27: Prevalence of NNRTI, NRTI, and PI resistance among pre-treatment viremic PLHIV by survey round**

|  |  |  | **NNRTI** | | |  |  |  | **NRTI** | | |  |  |  | **PI** | | |
| --- | --- | --- | --- | --- | --- | --- | --- | --- | --- | --- | --- | --- | --- | --- | --- | --- | --- |
| **Survey round** | ***n*** | **Obs (%)** | **Prev. % (95% CI)** | **Prev. ratio (95% CI)** | ***p*-value** |  | ***n*** | **Obs (%)** | **Prev. % (95% CI)** | **Prev. ratio (95% CI)** | ***p*-value** |  | ***n*** | **Obs (%)** | **Prev. % (95% CI)** | **Prev. ratio (95% CI)** | ***p*-value** |
| 2012 | 971 | 48 (4.94%) | 4.86 (3.69, 6.42) | ref | ref |  | 997 | 19 (1.91%) | 1.9 (1.17, 3.11) | ref | ref |  | 1361 | 29 (2.13%) | 2.08 (1.43, 3) | ref | ref |
| 2014 | 422 | 28 (6.64%) | 5.46 (3.77, 7.89) | 1.12 (0.72, 1.76) | 0.62 |  | 417 | 6 (1.44%) | 1.27 (0.52, 3.1) | 0.67 (0.23, 1.92) | 0.46 |  | 499 | 8 (1.6%) | 1.58 (0.81, 3.08) | 0.76 (0.35, 1.64) | 0.48 |
| 2015 | 705 | 50 (7.09%) | 7 (5.46, 8.97) | 1.44 (1.01, 2.05) | 0.045 |  | 665 | 13 (1.95%) | 2.09 (1.26, 3.45) | 1.1 (0.56, 2.15) | 0.79 |  | 706 | 9 (1.27%) | 1.36 (0.76, 2.4) | 0.65 (0.34, 1.27) | 0.21 |
| 2017 | 439 | 43 (9.79%) | 9.61 (7.27, 12.7) | 1.98 (1.34, 2.91) | 0.00058 |  | 430 | 8 (1.86%) | 2.05 (1.11, 3.78) | 1.08 (0.49, 2.38) | 0.85 |  | 439 | 9 (2.05%) | 1.97 (1.11, 3.5) | 0.95 (0.49, 1.84) | 0.88 |
| 2019 | 119 | 10 (8.4%) | 8.24 (4.72, 14.38) | 1.69 (0.91, 3.15) | 0.096 |  | 117 | 2 (1.71%) | 1.82 (0.5, 6.6) | 0.96 (0.24, 3.78) | 0.95 |  | 122 | 1 (0.82%) | 0.82 (0.15, 4.45) | 0.39 (0.07, 2.23) | 0.29 |

Estimates were generated using Poisson regression with robust standard errors with survey round as a predictor variable. Generalized estimating equations with correlation structure selection by Quasi Information Criterion value (NNRTI: exchangeable, NRTI: exchangeable, PI: AR1) were used to account for repeat participants across study rounds. 95% confidence intervals indicate the Wald confidence interval around the mean value in each category. *p*-values that coefficients are different from 0 at the $\alpha=0.05$ level were calculated using the Wald method. NNRTI = non-nucleoside reverse transcriptase inhibitors. NRTI = nucleoside reverse transcriptase inhibitors. PI = integrase inhibitors.

**Supplementary Table 28: Association between age, community type, and sex and the prevalence of NNRTI, NRTI and PI resistance among pre-treatment viremic PLHIV adjusted by survey round**

|  |  | **NNRTI** | |  | **NRTI** | |  | **PI** | |
| --- | --- | --- | --- | --- | --- | --- | --- | --- | --- |
| **Covariate** | **Variable** | **Coeff. % (95% CI)** | ***p*-value** |  | **Coeff. % (95% CI)** | ***p*-value** |  | **Coeff. % (95% CI)** | ***p*-value** |
| Age | Intercept | 0.05 (0.03, 0.08) | ref |  | 0.01 (0.01, 0.04) | ref |  | 0.02 (0.01, 0.05) | ref |
|  | 2014 | 1.12 (0.71, 1.76) | 0.62 |  | 0.66 (0.23, 1.93) | 0.45 |  | 0.75 (0.35, 1.64) | 0.47 |
|  | 2015 | 1.44 (1.01, 2.05) | 0.045 |  | 1.11 (0.56, 2.17) | 0.77 |  | 0.66 (0.34, 1.28) | 0.22 |
|  | 2017 | 1.97 (1.34, 2.91) | 0.00063 |  | 1.09 (0.5, 2.42) | 0.82 |  | 0.95 (0.49, 1.85) | 0.89 |
|  | 2019 | 1.7 (0.91, 3.15) | 0.094 |  | 0.97 (0.24, 3.84) | 0.96 |  | 0.4 (0.07, 2.34) | 0.31 |
|  | age (24,34] | 0.94 (0.63, 1.39) | 0.76 |  | 1.73 (0.69, 4.3) | 0.24 |  | 0.92 (0.47, 1.81) | 0.81 |
|  | age (34,49] | 0.91 (0.59, 1.42) | 0.68 |  | 0.79 (0.27, 2.31) | 0.66 |  | 0.66 (0.3, 1.43) | 0.29 |
| Community type | Intercept | 0.06 (0.04, 0.08) | ref |  | 0.02 (0.01, 0.03) | ref |  | 0.02 (0.01, 0.04) | ref |
|  | 2014 | 1.12 (0.72, 1.76) | 0.61 |  | 0.7 (0.24, 2.07) | 0.52 |  | 0.76 (0.35, 1.64) | 0.49 |
|  | 2015 | 1.43 (1, 2.05) | 0.048 |  | 1.14 (0.57, 2.25) | 0.71 |  | 0.65 (0.34, 1.26) | 0.21 |
|  | 2017 | 1.97 (1.34, 2.91) | 0.00061 |  | 1.09 (0.49, 2.4) | 0.83 |  | 0.95 (0.49, 1.83) | 0.87 |
|  | 2019 | 1.67 (0.89, 3.13) | 0.11 |  | 0.9 (0.24, 3.33) | 0.88 |  | 0.39 (0.07, 2.17) | 0.28 |
|  | Fishing | 0.83 (0.58, 1.2) | 0.32 |  | 0.79 (0.37, 1.7) | 0.55 |  | 0.97 (0.47, 2.01) | 0.94 |
|  | Trading | 0.76 (0.48, 1.2) | 0.24 |  | 1.81 (0.84, 3.9) | 0.13 |  | 1.05 (0.46, 2.37) | 0.92 |
| Sequencing approach | Intercept | 0.05 (0.04, 0.07) | ref |  | 0.02 (0.01, 0.03) | ref |  |  |  |
|  | 2014 | 1.13 (0.73, 1.75) | 0.58 |  | 0.76 (0.29, 1.98) | 0.57 |  |  |  |
|  | 2015 | 1.51 (0.7, 3.28) | 0.3 |  | 4.13 (0.12, 137.63) | 0.43 |  |  |  |
|  | 2017 | 2.08 (0.95, 4.57) | 0.069 |  | 4.07 (0.12, 141.58) | 0.44 |  |  |  |
|  | 2019 | 1.78 (0.7, 4.5) | 0.22 |  | 3.64 (0.09, 152.86) | 0.5 |  |  |  |
|  | Bait capture | 0.95 (0.44, 2.04) | 0.89 |  | 0.25 (0.01, 8.49) | 0.44 |  |  |  |
| Sex | Intercept | 0.06 (0.04, 0.07) | ref |  | 0.03 (0.02, 0.04) | ref |  | 0.02 (0.01, 0.03) | ref |
|  | 2014 | 1.15 (0.73, 1.79) | 0.55 |  | 0.71 (0.24, 2.06) | 0.52 |  | 0.76 (0.35, 1.65) | 0.49 |
|  | 2015 | 1.47 (1.03, 2.1) | 0.034 |  | 1.17 (0.59, 2.31) | 0.66 |  | 0.65 (0.34, 1.27) | 0.21 |
|  | 2017 | 2.03 (1.38, 3) | 0.00034 |  | 1.17 (0.53, 2.6) | 0.7 |  | 0.95 (0.49, 1.85) | 0.89 |
|  | 2019 | 1.73 (0.93, 3.23) | 0.083 |  | 1.01 (0.25, 4.11) | 0.98 |  | 0.39 (0.07, 2.26) | 0.3 |
|  | Male | 0.73 (0.53, 1.02) | 0.063 |  | 0.42 (0.21, 0.83) | 0.013 |  | 0.98 (0.54, 1.76) | 0.93 |

**E**stimates were generated using bivariate Poisson regression with robust standard errors with survey round and an epidemiological covariate as predictor variables. PI resistance was not evaluated by sequencing approach due to a limited number of outcome events. Generalized estimating equations with correlation structure selection by Quasi Information Criterion value based on the univariate analysis (NNRTI: exchangeable, NRTI: exchangeable, PI: AR1) were used to account for repeat participants across study rounds. 95% confidence intervals indicate the Wald confidence interval around the mean value in each category. *p*-values that coefficients are different from 0 at the $\alpha=0.05$ level were calculated using the Wald method. NNRTI = non-nucleoside reverse transcriptase inhibitors. NRTI = nucleoside reverse transcriptase inhibitors. PI = integrase inhibitors.

**Supplementary Table 29: Prevalence of NRTI resistance among pre-treatment viremic PLHIV stratified by sex**

|  |  | **NRTI** | | | | |
| --- | --- | --- | --- | --- | --- | --- |
| **Covariate** | **Strata** | ***n*** | **Obs (%)** | **Prev. (95% CI)** | **Prev. ratio (95% CI)** | ***p*-value** |
| Sex | F |  |  |  |  |  |
|  | 2012 | 541 | 12 (2.22%) | 2.28 (1.22, 4.29) | ref | ref |
|  | 2014 | 213 | 6 (2.82%) | 2.61 (1.09, 6.25) | 1.14 (0.37, 3.49) | 0.82 |
|  | 2015 | 314 | 10 (3.18%) | 3.19 (1.76, 5.81) | 1.4 (0.6, 3.28) | 0.44 |
|  | 2017 | 199 | 4 (2.01%) | 2.22 (0.93, 5.29) | 0.97 (0.32, 2.92) | 0.96 |
|  | 2019 | 60 | 1 (1.67%) | 1.57 (0.23, 10.66) | 0.69 (0.09, 5.15) | 0.71 |
|  | M |  |  |  |  |  |
|  | 2012 | 456 | 7 (1.54%) | 1.4 (0.68, 2.87) | ref | ref |
|  | 2014 | 204 | 0 (0%) | 0 (0, 0) | 0 (0, 0) | <0.0001 |
|  | 2015 | 351 | 3 (0.85%) | 1.06 (0.41, 2.74) | 0.76 (0.27, 2.14) | 0.6 |
|  | 2017 | 231 | 4 (1.73%) | 1.84 (0.76, 4.48) | 1.32 (0.43, 4) | 0.63 |
|  | 2019 | 57 | 1 (1.75%) | 1.94 (0.34, 11.18) | 1.39 (0.21, 9.04) | 0.73 |

**E**stimates were generated using bivariate Poisson regression with robust standard errors with survey round as a predictor variable stratified by covariate. Estimates only generated for combination of drug and covariate that were significant in the bivariate analysis. Generalized estimating equations with correlation structure selection by Quasi Information Criterion value based on the univariate analysis (exchangeable) were used to account for repeat participants across study rounds. 95% confidence intervals indicate the Wald confidence interval around the mean value in each category. *p*-values that coefficients are different from 0 at the $\alpha=0.05$ level were calculated using the Wald method. NNRTI = non-nucleoside reverse transcriptase inhibitors. NRTI = nucleoside reverse transcriptase inhibitors. PI = integrase inhibitors.

**Supplementary Table 30: Count of amino acid mutations observed among viremic PLHIV**

|  | Viremic participant-visits | | |  |  | Viremic participant-visits | | |  |  | Viremic participant-visits | | |
| --- | --- | --- | --- | --- | --- | --- | --- | --- | --- | --- | --- | --- | --- |
| Mutation | Total (N=2323) | Pre-treatment (N=1952) | Treatment-experienced (N=371) |  | Mutation | Total (N=2323) | Pre-treatment (N=1952) | Treatment-experienced (N=371) |  | Mutation | Total (N=2323) | Pre-treatment (N=1952) | Treatment-experienced (N=371) |
| inT97A | 257 | 218 | 39 |  | prK43T | 10 | 6 | 4 |  | prL89V | 2 | 2 | 0 |
| rtK103N | 203 | 94 | 109 |  | prM46L | 9 | 9 | 0 |  | rtT215A | 2 | 2 | 0 |
| rtM184V | 161 | 25 | 136 |  | rtK219R | 9 | 6 | 3 |  | rtT69G | 2 | 2 | 0 |
| rtE138A | 98 | 83 | 15 |  | rtV106M | 9 | 5 | 4 |  | rtV179L | 2 | 2 | 0 |
| rtY181C | 77 | 15 | 62 |  | rtK238T | 9 | 3 | 6 |  | rtV75A | 2 | 2 | 0 |
| rtG190A | 74 | 22 | 52 |  | rtA62V | 9 | 2 | 7 |  | inE138K | 2 | 1 | 1 |
| rtM41L | 71 | 49 | 22 |  | rtG190S | 9 | 1 | 8 |  | inS147G | 2 | 1 | 1 |
| rtK101E | 64 | 25 | 39 |  | prK20T | 8 | 6 | 2 |  | prI47A | 2 | 1 | 1 |
| prL33F | 61 | 53 | 8 |  | inG163R | 8 | 5 | 3 |  | prI47V | 2 | 1 | 1 |
| rtK65R | 60 | 14 | 46 |  | rtV179E | 8 | 5 | 3 |  | rtK219N | 2 | 1 | 1 |
| rtV108I | 45 | 19 | 26 |  | rtK103S | 8 | 4 | 4 |  | rtL234I | 2 | 1 | 1 |
| rtN348I | 39 | 11 | 28 |  | rtL210W | 8 | 2 | 6 |  | rtV75M | 2 | 1 | 1 |
| rtK70R | 38 | 13 | 25 |  | rtG190E | 7 | 6 | 1 |  | rtY188C | 2 | 1 | 1 |
| rtK219E | 34 | 9 | 25 |  | inQ95K | 7 | 5 | 2 |  | rtK101H | 2 | 0 | 2 |
| rtD67N | 33 | 12 | 21 |  | rtY181I | 6 | 4 | 2 |  | rtK238N | 2 | 0 | 2 |
| rtH221Y | 33 | 7 | 26 |  | rtK70T | 6 | 2 | 4 |  | inH51Y | 1 | 1 | 0 |
| rtV106I | 31 | 27 | 4 |  | rtK70Q | 6 | 1 | 5 |  | inN155T | 1 | 1 | 0 |
| prV32I | 31 | 26 | 5 |  | rtM230L | 6 | 0 | 6 |  | inQ148R | 1 | 1 | 0 |
| inE157Q | 28 | 23 | 5 |  | rtK70N | 5 | 5 | 0 |  | prI50L | 1 | 1 | 0 |
| rtA98G | 28 | 12 | 16 |  | rtT215S | 5 | 5 | 0 |  | prL23I | 1 | 1 | 0 |
| rtV179D | 26 | 22 | 4 |  | rtK103T | 5 | 4 | 1 |  | prN83D | 1 | 1 | 0 |
| rtP225H | 25 | 3 | 22 |  | rtY188L | 5 | 4 | 1 |  | prV82A | 1 | 1 | 0 |
| rtM184I | 24 | 8 | 16 |  | prL24I | 5 | 3 | 2 |  | prV82L | 1 | 1 | 0 |
| rtE138G | 22 | 15 | 7 |  | rtE138Q | 5 | 3 | 2 |  | rtK65E | 1 | 1 | 0 |
| rtT215F | 21 | 3 | 18 |  | rtT215I | 5 | 3 | 2 |  | rtT215E | 1 | 1 | 0 |
| rtE138K | 20 | 19 | 1 |  | rtV106A | 5 | 1 | 4 |  | rtT215L | 1 | 1 | 0 |
| rtY115F | 19 | 4 | 15 |  | rtT215V | 5 | 0 | 5 |  | rtY181F | 1 | 1 | 0 |
| prQ58E | 18 | 17 | 1 |  | rtK101P | 4 | 4 | 0 |  | rtY188H | 1 | 1 | 0 |
| prM46I | 18 | 15 | 3 |  | inG163K | 4 | 3 | 1 |  | inR263K | 1 | 0 | 1 |
| rtK70E | 16 | 4 | 12 |  | prG48A | 4 | 2 | 2 |  | prV82M | 1 | 0 | 1 |
| rtK219Q | 15 | 4 | 11 |  | rtF227L | 4 | 1 | 3 |  | rtD67E | 1 | 0 | 1 |
| rtT215Y | 15 | 3 | 12 |  | rtY318F | 4 | 0 | 4 |  | rtD67S | 1 | 0 | 1 |
| inE92G | 13 | 13 | 0 |  | prF53L | 3 | 3 | 0 |  | rtG190Q | 1 | 0 | 1 |
| rtD67G | 13 | 7 | 6 |  | prG73T | 3 | 3 | 0 |  |  |  |  |  |
| prL10F | 12 | 12 | 0 |  | rtM230I | 3 | 3 | 0 |  |  |  |  |  |
| rtV75I | 12 | 9 | 3 |  | rtL74V | 3 | 2 | 1 |  |  |  |  |  |
| rtL74I | 12 | 1 | 11 |  | rtD67T | 3 | 1 | 2 |  |  |  |  |  |
| rtL100I | 12 | 0 | 12 |  | rtT69D | 3 | 0 | 3 |  |  |  |  |  |
| prD30N | 10 | 10 | 0 |  | prI54V | 2 | 2 | 0 |  |  |  |  |  |

**Supplementary Table 31: Prevalence of amino acid mutations among viremic pre-treatment PLHIV by survey round**

|  |  | **inT97A** | | | |  | **rtK103N** | | | |  | **rtE138A** | | | |
| --- | --- | --- | --- | --- | --- | --- | --- | --- | --- | --- | --- | --- | --- | --- | --- |
| **Survey round** | *n* | **Obs (%)** | **Prev. % (95% CI)** | **Prev. ratio (95% CI)** | ***p*-value** |  | **Obs (%)** | **Prev. % (95% CI)** | **Prev. ratio (95% CI)** | ***p*-value** |  | **Obs (%)** | **Prev. % (95% CI)** | **Prev. ratio (95% CI)** | ***p*-value** |
| 2012 | 482 | 44 (9.13%) | 9.97 (7.3, 13.61) | ref | ref |  | 11 (2.28%) | 1.67 (0.9, 3.09) | ref | ref |  | 9 (1.87%) | 2.03 (1.02, 4.02) | ref | ref |
| 2014 | 267 | 29 (10.86%) | 10.88 (7.33, 16.15) | 1.09 (0.66, 1.8) | 0.73 |  | 10 (3.75%) | 2.46 (1.26, 4.79) | 1.47 (0.59, 3.65) | 0.4 |  | 7 (2.62%) | 2.7 (1.18, 6.21) | 1.33 (0.47, 3.78) | 0.59 |
| 2015 | 656 | 62 (9.45%) | 9.57 (7.53, 12.17) | 0.96 (0.65, 1.42) | 0.84 |  | 26 (3.96%) | 3.93 (2.68, 5.76) | 2.35 (1.18, 4.72) | 0.016 |  | 25 (3.81%) | 4.03 (2.74, 5.95) | 1.99 (0.94, 4.2) | 0.072 |
| 2017 | 430 | 45 (10.47%) | 10.56 (8.01, 13.93) | 1.06 (0.7, 1.61) | 0.78 |  | 31 (7.21%) | 7.11 (5.06, 10) | 4.26 (2.11, 8.59) | <0.0001 |  | 12 (2.79%) | 2.77 (1.59, 4.85) | 1.37 (0.57, 3.3) | 0.49 |
| 2019 | 117 | 24 (20.51%) | 20 (13.89, 28.8) | 2.01 (1.24, 3.24) | 0.0044 |  | 7 (5.98%) | 6.01 (2.9, 12.46) | 3.6 (1.39, 9.33) | 0.0085 |  | 3 (2.56%) | 2.39 (0.78, 7.34) | 1.18 (0.32, 4.38) | 0.81 |
|  |  | **rtM41L** | | | |  | **prL33F** | | | |  | **rtV179D** | | | |
| **Survey round** | *n* | **Obs (%)** | **Prev. % (95% CI)** | **Prev. ratio (95% CI)** | ***p*-value** |  | **Obs (%)** | **Prev. % (95% CI)** | **Prev. ratio (95% CI)** | ***p*-value** |  | **Obs (%)** | **Prev. % (95% CI)** | **Prev. ratio (95% CI)** | ***p*-value** |
| 2012 | 482 | 4 (0.83%) | 2.15 (0.8, 5.79) | ref | ref |  | 4 (0.83%) | 1.02 (0.36, 2.86) | ref | ref |  | 2 (0.41%) | 0.74 (0.17, 3.25) | ref | ref |
| 2014 | 267 | 2 (0.75%) | 0.69 (0.17, 2.77) | 0.32 (0.06, 1.77) | 0.19 |  | 4 (1.5%) | 1.81 (0.51, 6.45) | 1.77 (0.34, 9.08) | 0.49 |  | 2 (0.75%) | 1.07 (0.24, 4.76) | 1.45 (0.18, 11.84) | 0.73 |
| 2015 | 656 | 18 (2.74%) | 2.77 (1.75, 4.39) | 1.29 (0.43, 3.83) | 0.65 |  | 6 (0.91%) | 0.8 (0.36, 1.8) | 0.78 (0.21, 2.9) | 0.71 |  | 7 (1.07%) | 0.91 (0.43, 1.92) | 1.23 (0.24, 6.47) | 0.8 |
| 2017 | 430 | 10 (2.33%) | 2.41 (1.3, 4.45) | 1.12 (0.35, 3.58) | 0.85 |  | 7 (1.63%) | 1.69 (0.81, 3.53) | 1.66 (0.47, 5.87) | 0.43 |  | 7 (1.63%) | 1.65 (0.79, 3.45) | 2.24 (0.43, 11.68) | 0.34 |
| 2019 | 117 | 7 (5.98%) | 5.66 (2.72, 11.77) | 2.63 (0.77, 9) | 0.12 |  | 2 (1.71%) | 1.53 (0.38, 6.05) | 1.49 (0.27, 8.34) | 0.65 |  | 2 (1.71%) | 2.1 (0.51, 8.65) | 2.84 (0.37, 22.01) | 0.32 |
|  |  | **rtK101E** | | | |  | **rtV108I** | | | |  | **prQ58E** | | | |
| **Survey round** | *n* | **Obs (%)** | **Prev. % (95% CI)** | **Prev. ratio (95% CI)** | ***p*-value** |  | **Obs (%)** | **Prev. % (95% CI)** | **Prev. ratio (95% CI)** | ***p*-value** |  | **Obs (%)** | **Prev. % (95% CI)** | **Prev. ratio (95% CI)** | ***p*-value** |
| 2012 | 482 | 4 (0.83%) | 0.73 (0.27, 1.94) | ref | ref |  | 5 (1.04%) | 0.84 (0.35, 2.04) | ref | ref |  | 3 (0.62%) | 0.61 (0.19, 1.94) | ref | ref |
| 2014 | 267 | 3 (1.12%) | 1.29 (0.37, 4.43) | 1.77 (0.37, 8.55) | 0.48 |  | 1 (0.37%) | 0.32 (0.05, 2.29) | 0.38 (0.04, 3.29) | 0.38 |  | 2 (0.75%) | 0.26 (0.06, 1.03) | 0.42 (0.07, 2.56) | 0.35 |
| 2015 | 656 | 7 (1.07%) | 1.15 (0.55, 2.4) | 1.58 (0.46, 5.38) | 0.47 |  | 5 (0.76%) | 0.83 (0.35, 2.01) | 0.99 (0.33, 2.96) | 0.99 |  | 2 (0.3%) | 0.35 (0.09, 1.39) | 0.57 (0.09, 3.45) | 0.54 |
| 2017 | 431 | 5 (1.16%) | 1.21 (0.51, 2.9) | 1.67 (0.45, 6.19) | 0.44 |  | 4 (0.93%) | 0.95 (0.36, 2.52) | 1.12 (0.3, 4.2) | 0.86 |  | 4 (0.93%) | 0.94 (0.35, 2.5) | 1.54 (0.34, 6.99) | 0.58 |
| 2019 | 117 | 3 (2.56%) | 2.27 (0.74, 7.04) | 3.13 (0.7, 13.93) | 0.13 |  | 0 (0%) | 0 (0, 0) | 0 (0, 0) | <0.0001 |  | 0 (0%) | 0 (0, 0) | 0 (0, 0) | <0.0001 |
|  |  | **rtG190A** | | | |  |  |  |  |  |  |  |  |  |  |
| **Survey round** | *n* | **Obs (%)** | **Prev. % (95% CI)** | **Prev. ratio (95% CI)** | ***p*-value** |  |  |  |  |  |  |  |  |  |  |
| 2012 | 482 | 4 (0.83%) | 0.84 (0.29, 2.39) | ref | ref |  |  |  |  |  |  |  |  |  |  |
| 2014 | 267 | 3 (1.12%) | 1.07 (0.32, 3.5) | 1.28 (0.26, 6.25) | 0.76 |  |  |  |  |  |  |  |  |  |  |
| 2015 | 656 | 5 (0.76%) | 0.68 (0.28, 1.63) | 0.81 (0.2, 3.19) | 0.76 |  |  |  |  |  |  |  |  |  |  |
| 2017 | 431 | 4 (0.93%) | 0.94 (0.35, 2.5) | 1.12 (0.27, 4.73) | 0.87 |  |  |  |  |  |  |  |  |  |  |
| 2019 | 117 | 2 (1.71%) | 1.51 (0.38, 6) | 1.81 (0.32, 10.25) | 0.5 |  |  |  |  |  |  |  |  |  |  |

Estimates were generated using Poisson regression with robust standard errors with survey round as a predictor variable. Generalized estimating equations with independent correlation structure to aid convergence for rare mutations were used. 95% confidence intervals indicate the Wald confidence interval around the mean value in each category. *p*-values that coefficients are different from 0 at the $\alpha=0.05$ level were calculated using the Wald method. NNRTI = non-nucleoside reverse transcriptase inhibitors. NRTI = nucleoside reverse transcriptase inhibitors. PI = integrase inhibitors.

**Supplementary Table 32: Prevalence of NNRTI, NRTI, and PI resistance among treatment-experienced viremic PLHIV by survey round**

|  | **NNRTI** | | | | |  | **NRTI** | | | | |  | **PI** | | | | |
| --- | --- | --- | --- | --- | --- | --- | --- | --- | --- | --- | --- | --- | --- | --- | --- | --- | --- |
| **Survey round** | ***n*** | **Obs (%)** | **Prev. (95% CI)** | **Prev. ratio (95% CI)** | ***p*-value** |  | ***n*** | **Obs (%)** | **Prev. (95% CI)** | **Prev. ratio (95% CI)** | ***p*-value** |  | ***n*** | **Obs (%)** | **Prev. (95% CI)** | **Prev. ratio (95% CI)** | ***p*-value** |
| 2015 | 160 | 87 (54.37%) | 51.77 (46.47, 57.67) | ref | ref |  | 152 | 74 (48.68%) | 50.04 (42.59, 58.79) | ref | ref |  | 157 | 5 (3.18%) | 3.28 (1.38, 7.82) | ref | ref |
| 2017 | 184 | 87 (47.28%) | 51.49 (46.24, 57.34) | 0.99 (0.97, 1.02) | 0.72 |  | 182 | 66 (36.26%) | 36.46 (30.06, 44.22) | 0.73 (0.58, 0.92) | 0.0084 |  | 185 | 4 (2.16%) | 2.13 (0.81, 5.63) | 0.65 (0.18, 2.36) | 0.51 |

Estimates were generated using Poisson regression with robust standard errors with survey round as a predictor variable. Generalized estimating equations with correlation structure selection by Quasi Information Criterion value (NNRTI: exchangeable, NRTI: independent, PI: exchangeable) were used to account for repeat participants across study rounds. 95% confidence intervals indicate the Wald confidence interval around the mean value in each category. *p*-values that coefficients are different from 0 at the $\alpha=0.05$ level were calculated using the Wald method. NNRTI = non-nucleoside reverse transcriptase inhibitors. NRTI = nucleoside reverse transcriptase inhibitors. PI = integrase inhibitors.

**Supplementary Table 33: Association between age, community type, and sex and the prevalence of NNRTI, NRTI and PI resistance among treatment-experienced viremic PLHIV adjusted by survey round**

|  |  | **NNRTI** | |  | **NRTI** | |  | **PI** | |
| --- | --- | --- | --- | --- | --- | --- | --- | --- | --- |
| **Covariate** | **Variable** | **Coeff. % (95% CI)** | ***p*-value** |  | **Coeff. % (95% CI)** | ***p*-value** |  | **Coeff. % (95% CI)** | ***p*-value** |
| Age | Intercept | 0.48 (0.4, 0.58) | ref |  | 0.34 (0.22, 0.51) | ref |  | 0.04 (0.01, 0.13) | ref |
|  | 2015 | 0.97 (0.93, 1.01) | 0.094 |  | 0.7 (0.56, 0.89) | 0.0036 |  | 0.66 (0.16, 2.74) | 0.57 |
|  | age (24,34] | 1.1 (0.93, 1.31) | 0.27 |  | 1.62 (1.03, 2.56) | 0.037 |  | 0.82 (0.13, 5.31) | 0.84 |
|  | age (34,49] | 1.09 (0.89, 1.35) | 0.41 |  | 1.56 (0.98, 2.47) | 0.061 |  | 0.8 (0.12, 5.12) | 0.81 |
| Community type | Intercept | 0.52 (0.43, 0.64) | ref |  | 0.54 (0.43, 0.69) | ref |  | 0.03 (0.01, 0.14) | ref |
|  | 2015 | 1.04 (0.95, 1.14) | 0.42 |  | 0.74 (0.58, 0.94) | 0.013 |  | 0.65 (0.18, 2.3) | 0.5 |
|  | Fishing | 0.89 (0.69, 1.15) | 0.39 |  | 0.86 (0.63, 1.16) | 0.32 |  | 1.17 (0.24, 5.68) | 0.84 |
|  | Trading | 1.08 (0.81, 1.44) | 0.59 |  | 0.94 (0.67, 1.33) | 0.73 |  | 1.29 (0.18, 9.01) | 0.8 |
| Sex | Intercept | 0.54 (0.46, 0.64) | ref |  | 0.54 (0.45, 0.64) | ref |  | 0.04 (0.01, 0.09) | ref |
|  | 2015 | 1.13 (0.89, 1.43) | 0.32 |  | 0.74 (0.58, 0.94) | 0.012 |  | 0.67 (0.18, 2.49) | 0.55 |
|  | Male | 0.74 (0.58, 0.93) | 0.011 |  | 0.82 (0.62, 1.09) | 0.17 |  | 0.74 (0.18, 3.04) | 0.68 |

**E**stimates were generated using bivariate Poisson regression with robust standard errors with survey round and an epidemiological covariate as predictor variables. PI resistance was not evaluated by sequencing approach due to a limited number of outcome events. Generalized estimating equations with correlation structure selection by Quasi Information Criterion value based on the univariate analysis (NNRTI: exchangeable, NRTI: independent, PI: exchangeable) were used to account for repeat participants across study rounds. 95% confidence intervals indicate the Wald confidence interval around the mean value in each category. *p*-values that coefficients are different from 0 at the $\alpha=0.05$ level were calculated using the Wald method. NNRTI = non-nucleoside reverse transcriptase inhibitors. NRTI = nucleoside reverse transcriptase inhibitors. PI = integrase inhibitors.

**Supplementary Table 34: Prevalence of NNRTI, NRTI, and PI resistance among treatment-experienced viremic PLHIV by survey round and age, community type, and sex**

|  |  | **NNRTI** | | | | |  | **NRTI** | | | | |
| --- | --- | --- | --- | --- | --- | --- | --- | --- | --- | --- | --- | --- |
| **Covariate** | **Strata** | ***n*** | **Obs (%)** | **Prev. (95% CI)** | **Prev. ratio (95% CI)** | ***p*-value** |  | ***n*** | **Obs (%)** | **Prev. (95% CI)** | **Prev. ratio (95% CI)** | ***p*-value** |
| Age category | age (14,24] |  |  |  |  |  |  |  |  |  |  |  |
|  | 2015 | .. | .. | .. | .. | .. |  | 31 | 8 (25.81%) | 26.43 (14.37, 48.59) | ref | ref |
|  | 2017 | .. | .. | .. | .. | .. |  | 23 | 8 (34.78%) | 35.24 (19.94, 62.28) | 1.33 (0.59, 3.01) | 0.49 |
|  | age (24,34] |  |  |  |  |  |  |  |  |  |  |  |
|  | 2015 | .. | .. | .. | .. | .. |  | 62 | 33 (53.23%) | 54.43 (43.16, 68.64) | ref | ref |
|  | 2017 | .. | .. | .. | .. | .. |  | 97 | 38 (39.18%) | 39.25 (30.57, 50.39) | 0.72 (0.52, 0.99) | 0.044 |
|  | age (34,49] |  |  |  |  |  |  |  |  |  |  |  |
|  | 2015 | .. | .. | .. | .. | .. |  | 59 | 33 (55.93%) | 57.49 (46.01, 71.84) | ref | ref |
|  | 2017 | .. | .. | .. | .. | .. |  | 62 | 20 (32.26%) | 32.68 (22.74, 46.95) | 0.57 (0.38, 0.85) | 0.0063 |
| Sex | F |  |  |  |  |  |  |  |  |  |  |  |
|  | 2015 | 98 | 55 (56.12%) | 57.03 (43.02, 75.6) | ref | ref |  | .. | .. | .. | .. | .. |
|  | 2017 | 100 | 57 (57%) | 58.17 (44.13, 76.68) | 1.02 (0.62, 1.68) | 0.94 |  | .. | .. | .. | .. | .. |
|  | M |  |  |  |  |  |  | .. | .. | .. | .. | .. |
|  | 2015 | 62 | 32 (51.61%) | 42.27 (34.43, 51.9) | ref | ref |  | .. | .. | .. | .. | .. |
|  | 2017 | 84 | 30 (35.71%) | 43.16 (35.33, 52.71) | 1.02 (0.94, 1.11) | 0.62 |  | .. | .. | .. | .. | .. |

**E**stimates were generated using bivariate Poisson regression with robust standard errors with survey round as a predictor variable stratified by covariate. Estimates only generated for combination of drug and covariate that were significant in the bivariate analysis. Generalized estimating equations with correlation structure selection by Quasi Information Criterion value based on the univariate analysis (NNRTI: exchangeable, NRTI: independent, PI: exchangeable) were used to account for repeat participants across study rounds. 95% confidence intervals indicate the Wald confidence interval around the mean value in each category. *p*-values that coefficients are different from 0 at the $\alpha=0.05$ level were calculated using the Wald method. NNRTI = non-nucleoside reverse transcriptase inhibitors. NRTI = nucleoside reverse transcriptase inhibitors. PI = integrase inhibitors.

**Supplementary Table 35: Prevalence of amino acid mutations among treatment-experienced viremic PLHIV**

|  |  | **rtM184V** | | | |  | **rtK103N** | | | |  | **rtY181C** | | | |
| --- | --- | --- | --- | --- | --- | --- | --- | --- | --- | --- | --- | --- | --- | --- | --- |
| **Survey round** | *n* | **Obs (%)** | **Prev. % (95% CI)** | **Prev. ratio (95% CI)** | ***p*-value** |  | **Obs (%)** | **Prev. % (95% CI)** | **Prev. ratio (95% CI)** | ***p*-value** |  | **Obs (%)** | **Prev. % (95% CI)** | **Prev. ratio (95% CI)** | ***p*-value** |
| 2015 | 143 | 57 (39.86%) | 40.76 (33.32, 49.86) | ref | ref |  | 43 (30.07%) | 31.8 (24.83, 40.73) | ref | ref |  | 31 (21.68%) | 22.35 (16.34, 30.56) | ref | ref |
| 2017 | 181 | 61 (33.7%) | 33.89 (27.61, 41.59) | 0.83 (0.63, 1.09) | 0.19 |  | 47 (25.97%) | 25.68 (20.03, 32.92) | 0.81 (0.58, 1.13) | 0.21 |  | 26 (14.36%) | 14.67 (10.27, 20.95) | 0.66 (0.43, 1.01) | 0.056 |
|  |  | **rtG190A** | | | |  | **rtK65R** | | | |  | **inT97A** | | | |
| **Survey round** | *n* | **Obs (%)** | **Prev. % (95% CI)** | **Prev. ratio (95% CI)** | ***p*-value** |  | **Obs (%)** | **Prev. % (95% CI)** | **Prev. ratio (95% CI)** | ***p*-value** |  | **Obs (%)** | **Prev. % (95% CI)** | **Prev. ratio (95% CI)** | ***p*-value** |
| 2015 | 143 | 19 (13.29%) | 13.21 (8.64, 20.21) | ref | ref |  | 20 (13.99%) | 13.41 (8.85, 20.31) | ref | ref |  | 13 (9.09%) | 8.29 (4.89, 14.05) | ref | ref |
| 2017 | 181 | 22 (12.15%) | 12.34 (8.33, 18.27) | 0.93 (0.55, 1.59) | 0.8 |  | 22 (12.15%) | 12.07 (8.14, 17.9) | 0.9 (0.53, 1.53) | 0.7 |  | 18 (9.94%) | 9.96 (6.41, 15.48) | 1.2 (0.61, 2.38) | 0.6 |
|  |  | **rtK101E** | | | |  | **rtK219E** | | | |  | **rtH221Y** | | | |
| **Survey round** | *n* | **Obs (%)** | **Prev. % (95% CI)** | **Prev. ratio (95% CI)** | ***p*-value** |  | **Obs (%)** | **Prev. % (95% CI)** | **Prev. ratio (95% CI)** | ***p*-value** |  | **Obs (%)** | **Prev. % (95% CI)** | **Prev. ratio (95% CI)** | ***p*-value** |
| 2015 | 143 | 15 (10.49%) | 10.1 (6.21, 16.44) | ref | ref |  | 10 (6.99%) | 6.93 (3.78, 12.7) | ref | ref |  | 11 (7.69%) | 7.41 (4.17, 13.19) | ref | ref |
| 2017 | 181 | 17 (9.39%) | 9.51 (6.04, 14.97) | 0.94 (0.51, 1.75) | 0.85 |  | 14 (7.73%) | 7.64 (4.6, 12.67) | 1.1 (0.51, 2.36) | 0.8 |  | 13 (7.18%) | 7.35 (4.34, 12.43) | 0.99 (0.45, 2.16) | 0.98 |
|  |  | **rtV108I** | | | |  |  |  |  |  |  |  |  |  |  |
| **Survey round** | *n* | **Obs (%)** | **Prev. % (95% CI)** | **Prev. ratio (95% CI)** | ***p*-value** |  |  |  |  |  |  |  |  |  |  |
| 2015 | 143 | 8 (5.59%) | 6.33 (3.23, 12.42) | ref | ref |  |  |  |  |  |  |  |  |  |  |
| 2017 | 181 | 13 (7.18%) | 7.08 (4.18, 11.99) | 1.12 (0.5, 2.52) | 0.79 |  |  |  |  |  |  |  |  |  |  |

Estimates were generated using Poisson regression with robust standard errors with survey round as a predictor variable. Generalized estimating equations with independent correlation structure to aid convergence for rare mutations were used. 95% confidence intervals indicate the Wald confidence interval around the mean value in each category. *p*-values that coefficients are different from 0 at the $\alpha=0.05$ level were calculated using the Wald method. NNRTI = non-nucleoside reverse transcriptase inhibitors. NRTI = nucleoside reverse transcriptase inhibitors. PI = integrase inhibitors.
